# Real-world data: A systematic literature review on the barriers, challenges, and opportunities associated with their inclusion in the health technology assessment process

**DOI:** 10.1101/2023.10.18.23297151

**Authors:** Konstantinos Zisis, Elpida Pavi, Mary Geitona, Kostas Athanasakis

**Affiliations:** Laboratory for Health Technology Assessment (LabHTA), Department of Public Health Policy, School of Public Health, University of West Attica, Athens, Greece; Institute for Health Economics, Athens, Greece; Department of Social & Educational Policy, Faculty of Social and Political Sciences, University of Peloponnese, Corinth, Greece

**Author notes:** Corresponding author: Konstantinos Zisis. These authors contributed equally to this work (Konstantinos Zisis, Elpida Pavi, Kostas Athanasakis). These authors also contributed equally to this work.

**Keywords:** Key-words: Real-world data, real-world evidence, health technology assessment, acceptance, barriers, challenges, opportunities, systematic review

## Abstract

**Objective:** This systematic review aimed to assess the current use and acceptance of real-world data (RWD) and real-world evidence (RWE) in health technology assessment (HTA) process. It additionally aimed to discern stakeholders’ viewpoints concerning RWD and RWE in HTA and illuminate the obstacles, difficulties, prospects, and consequences associated with the incorporation of RWD and RWE into the realm of HTA.

**Methods:** A comprehensive PRISMA-based systematic review was performed in July 2022 in PubMed/Medline, Scopus, IDEAS-RePEc, International HTA database, and Centre for Reviews and Dissemination with ad-hoc supplementary search in Google Scholar and international organization websites. The review included pre-determined inclusion criteria while the selection of eligible studies, the data extraction process and quality assessment were carried out using standardized and transparent methods.

**Results:** Twenty-nine (n=29) studies were included in the review out of 2.115 studies identified by the search strategy. In various global contexts, disparities in RWD utilization were evident, with randomized controlled trials (RCTs) serving as the primary evidence source. RWD and RWE played pivotal roles, surpassing relative effectiveness assessments (REAs) and significantly influencing decision-making and cost-effectiveness analyses. Identified challenges impeding RWD integration into HTA encompassed limited local data access, complexities in non-randomized trial design, data quality, privacy, and fragmentation. Addressing these is imperative for optimal RWD utilization. Incorporating RWD/RWE in HTA yields multifaceted advantages, enhancing understanding of treatment efficacy, resource utilization, and cost analysis, particularly via patient registries. RWE complements assessments of advanced therapy medicinal products (ATMPs) and rare diseases. Local data utilization strengthens HTA, bridging gaps when RCT data is lacking. RWD aids medical device decision-making, cancer drug reassessment, and indirect treatment comparisons. Challenges include data availability, stakeholder acceptance, expertise, and privacy. However, standardization, training, collaboration, and guidance can surmount these barriers, fostering enhanced RWD utilization in HTA.

**Conclusions:** RWD and RWE are recognized as valuable when RCTs are lacking. Acceptance and use of RWD/RWE vary, with challenges including limited availability, methodological issues, expertise gaps, fragmentation, and data quality concerns. Addressing these barriers is crucial for effective utilization in HTA.

## 1. Introduction

RWE and RWD are increasingly used for evaluating health technologies to inform decision-making in the healthcare sector. RWD refers to data related to patient health status and/or the delivery of healthcare that are routinely collected from various sources outside of traditional clinical trial settings. RWE refers to data generated from RWD and it’s actually the clinical evidence about the usage, benefits, and risks of medical products, which is derived from the analysis of RWD. The evidence derives from sources such as electronic health records, claims data, product or disease registries, pragmatic trials, and data generated by patients (patient-reported outcomes) as well as digital health technologies, among others [1, 2]. RWE can provide a more comprehensive and representative picture of how treatments and interventions work in real-world conditions, beyond the controlled environment of clinical trials. The role of RWE is undergoing continuous development and broadening while has gained prominence in healthcare decision-making, particularly during the COVID-19 pandemic [3]. While RCTs are still considered the benchmark for assessing the effectiveness of treatments including new cancer treatments, there is a growing consensus that relying solely on RCTs may not provide comprehensive solutions to all pertinent clinical or research inquiries and RWE can contribute in advancing decisions by providing complementary evidence [4].

In a general context, the advantages of using RWD in patient care are to:

- Evaluate the effectiveness and safety of treatments and interventions in real-world populations and environments provides a more holistic view of patient health and care outcomes, as data is derived from routine clinical care rather than controlled settings
- generate data on subpopulations that may be underrepresented in clinical trials by capturing a wider range of patient populations and health conditions, including underrepresented groups, and identify rare or long-term adverse events that may not be captured in clinical trials
- monitor the safety and efficacy of new treatments or interventions in real-world settings, beyond the limited scope of clinical trials [5].

The utilization of RWD and the generation of RWE hold immense promise for transforming healthcare decision-making. However, there are also challenges associated with the use of RWD, including issues related to inconsistent data quality, comparability and bias (subject to bias and measurement errors, both random and non-random) [6], as well as the need for appropriate statistical methods and analytical frameworks. Such challenges among others, are the following:

▪ Data Quality and Consistency: RWD originates from various sources in the real-world healthcare ecosystem, including electronic health records, claims databases, and patient registries. Consequently, data quality can be inconsistent due to differences in data collection methodologies and standards across healthcare institutions. Incomplete, inaccurate, or missing data can lead to flawed analyses and unreliable conclusions. Furthermore, the diverse nature of RWD sources means that data may vary in terms of completeness, timeliness, and relevance.
▪ Bias and Measurement Errors: RWD is inherently subject to bias and measurement errors, which can emanate from several sources. Selection bias can occur when certain patient populations are overrepresented or underrepresented in the data due to factors such as healthcare seeking behavior or data collection practices. Information bias may arise from discrepancies in the way data is recorded or measured, leading to inaccuracies. Additionally, non-random error can be introduced through factors like data entry mistakes, misclassification of variables, or systematic differences in data collection across institutions. These biases and errors can skew RWE findings, potentially leading to misleading conclusions about the safety and effectiveness of medical interventions [7].

Considering the formidable challenges inherent in the field, it is noteworthy that the prominence of RWE in shaping healthcare decision-making continues to ascend and the importance of RWE in healthcare decision-making is growing. Regulatory agencies such as the U.S. Food and Drug Administration (FDA) recognize its potential and have issued guidance on its use in regulatory decision-making. These guidelines provide a structured framework for how RWE can be employed to support various stages of drug development and post-market surveillance. For example, the FDA has issued guidance on the use of RWE in regulatory decision-making [8], while the Institute for Clinical and Economic Review (ICER) has developed a framework [9] for integrating RWE into coverage decisions and acknowledges the value of RWE in evaluating the real-world effectiveness and cost-effectiveness of medical interventions, particularly in comparison to traditional clinical trial evidence. While RWD is progressively attaining prominence in influencing healthcare decision-making, it remains a subject of discernible complexity and resistance within the healthcare milieu.

Based on the above, the objective of the study is to investigate the integration of real-world data and real-world evidence in health technology assessment process around the world. In particular, the aim of this systematic review was to capture, through a comprehensive systematic review: a) the current use and acceptance of RWD and RWE in the health technology assessment process; b) the prioritization of barriers, challenges, opportunities, and potential implications arising from the integration of evidence generated from RWD/RWE within the HTA process; c) the identification of stakeholders’ perspectives concerning to RWD and RWE in the HTA process.

## 2. Materials and Methods

Considering the above objective, the research questions defined for this review were the following:

⮚ Is the utilization and acceptance of RWD and RWE prevalent in the HTA process?
⮚ What are the barriers, challenges, potential benefits and feasibilities, as well as opportunities presented by the integration of RWD into the HTA process?
⮚ What are the viewpoints and declarations of stakeholders concerning to RWD and RWE in the HTA process?

No formal protocol was established or registered for this systematic review.

### 2.1 Study design, inclusion and exclusion criteria

A PRISMA-based systematic review [10, 11] was conducted to identify articles assessed by the researchers, employing inclusion criteria to ascertain study eligibility aligned with the review’s objectives. The search strategy, as detailed in section 2.2, was utilized to encompass these criteria. Inclusion criteria were as follows:

⮚ Population: There were no restrictions on populations, and studies were included from populations and sub-populations from all countries around the world and without any unique chracteristics.
⮚ Intervention: RWD and evidence that arise from the use and analysis of RWD.
⮚ Comparator: No comparator.
⮚ Outcomes: Data regarding the current use of RWD in HTA, the barriers, challenges, weaknesses of their integration in the process, opportunities, as well as the perspectives of stakeholders regarding the use of RWD in the HTA process were included. Additionally, data related to the views of stakeholders regarding RWD in the HTA process were also included.
⮚ Types of studies: All types of studies were included, such as reviews, policy texts, primary research, RCTs, qualitative research studies.
⮚ Language: Only studies written in English were included.
⮚ Timeline: No time restrictions were specified for the publication of studies and policy reports.

The exclusion criteria for studies in this analysis were as follows:

⮚ Study Types: Abstracts (oral and posters) that did not include at least one of the above outcome criteria were excluded.
⮚ Language: Any other written language apart from English was not included in this review.

### 2.2 Search strategy

The detailed search strategy, which was performed on July 2022, is provided in S1 Appendix.

Search strategy was implemented to the following databases: PubMed/Medline, Scopus, IDEAS-RePEc, International HTA database, Centre for Reviews and Dissemination. In addition, supplementary ad-hoc searches for relevant information were performed on Google Scholar, as well as various international organizations such as the World Health Organization (WHO) and Organisation for Economic Co-operation and Development (OECD), and specific health technology assessment organizations such as National Institute for Health and Care Excellence (NICE), Haute Autorité de santé (HAS), and Institute for Clinical & Economic Review (ICER) to identify relevant texts and references related to the study objectives.

### 2.3 Study selection methods

The literature discovered through the search was archived in a bibliographic database (EndNote), with duplicate entries subsequently removed. A pilot training check process was conducted initially to ensure consistency in selection and identify areas for modifications in the inclusion criteria to provide a more comprehensive and explicit list of study types that would be considered eligible for this review. Two researchers independently checked a random sample of approximately fifty (50) titles and abstracts for eligibility, and a high level of agreement was achieved which indicates that the two researchers largely agreed on whether each of these documents met the inclusion criteria established for the study. After this, a single researcher checked the remaining titles and abstracts for eligibility. Later, the studies resulting from the removal of duplicate entries were uploaded into Abstrackr [12], a specialized software developed by Brown University and the Center for Evidence Synthesis in Health. All abstracts were examined, and full-text documents were retrieved for the files that were flagged for inclusion. The retrieved articles were then analyzed in detail based on the full text.

### 2.4 Data extraction and synthesis methods

The study data was meticulously extracted and organized into four tables, a process undertaken to streamline and enhance the subsequent analysis and synthesis of the information. The design of these tables was thoughtfully structured to systematically capture pertinent information derived from the selected studies. The first table contained details relevant to the characteristicts including author, year of study, country, study type, objectives, health technology studied, population and therapeutic category, and subcategory of real-world data. The second table was dedicated to encompassing data concerning the contemporary utilization and reception of RWD and RWE. In contrast, the third table comprehensively addressed the hurdles, challenges, and complexities encountered when integrating RWD-RWE into HTA. Meanwhile, the fourth table was designed to encompass the potential advantages, opportunities, and viability associated with the adoption of RWD-RWE within the realm of HTA. To ensure consistency and pinpoint any potential adjustments required for the data extraction model, two researchers initially conducted an independent pilot test on a random sample of ten (10) studies. During this process, an appropriate level of agreement was observed, denoting that there was a satisfactory degree of consensus or concurrence among the researchers involved in the extraction of data from the selected studies. The extraction of the remaining studies was conducted by a primary researcher, supported by a secondary researcher who remained readily available to offer assistance in clarifying information or in situations where the primary researcher encountered challenges or uncertainties during the extraction process.

### 2.5 Appraisal of methodological quality

The methodological quality of the studies included in this review was assessed using several critical appraisal tools, namely the Critical Appraisal Skills Programme (CASP) tool for qualitative research [13], the Joanna Briggs Institute (JBI) checklist for systematic reviews and evidence syntheses [14], and Joanna Briggs Institute (JBI) checklist for text and opinions [15]. Each tool evaluated different aspects of study quality by one reviewer, including the study design, data collection methods, data analysis, and reporting of results. For the assessment of each study done using the CASP tool, the reviewer assessed the quality of the study design, data collection methods, data analysis, and interpretation of findings. The Critical Appraisal Skills Programme (CASP) tool is the most used tool for quality appraisal in health-related qualitative evidence syntheses [16]. Meanwhile, the JBI checklist was used to evaluate the relevance of the studies to the review question, study design, sample size, data collection methods, data analysis, and reporting of results.

Quality appraisal, in detail, of eligible studies can be found in the S2 Appendix. In the overarching context, it is pertinent to elucidate that the quality of the incorporated studies exhibits a discernible spectrum, wherein, a number of studies may be aptly delineated as demonstrating a standard of moderate quality, while the preponderance of the corpus can be distinguished as manifesting a commendable standard of good quality.

## 3. Results

During the search process, a total of 2.115 studies were identified based on the pre-specified selection criteria after removing duplicates (n=50). Among these, 137 studies were selected for inclusion after title and abstract review. Full-text versions of all studies were obtained, with the exception of eleven studies whose authors did not respond to the request of their manuscript since were also not available in the literature. Following a thorough examination of the complete texts, 108 studies were excluded due inadequate data (n = 80), oral/poster presentations without much data (n = 14), non-availability of full-text (n = 11) and non-English manuscripts (n = 3). Eventually, 29 studies out of the 137 met the inclusion criteria and were considered eligible for analysis.

Figure 1 illustrates the study selection process in accordance with the PRISMA flow diagram.

**Figure 1.**
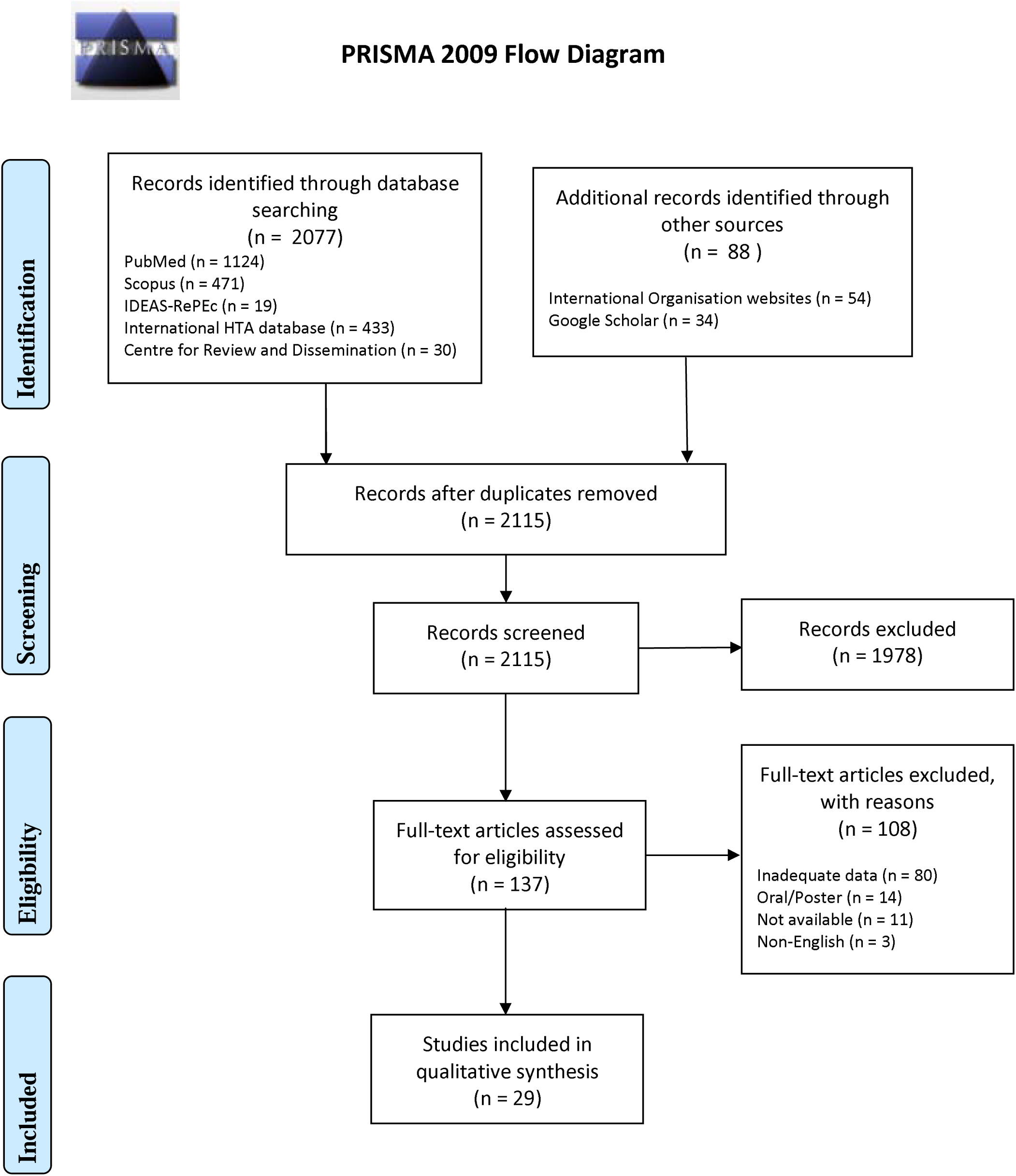
PRISMA flow chart of the search strategy.

### 3.1 Description of study characteristics

Overall, 29 studies were included in this review, among which, most of them were refering to European countries. Most of these 29 studies were referring to multiple countries within their analyses while few of them assessed information related to RWD/RWE for HTA in continents. In particular, England (n=6), Germany (n=5), UK (n=4), Sweden (n=4), Netherlands (n=3), Scotland (n=3), France (n=3), Norway (n=2), Italy (n=2), Spain (n=1), Austria (n=1), Denmark (n=1), and Belgium (n=1). In addition, Europe was referred in three studies, while one study included European Union countries and another one study included Central and Eastern Europe. On the other hand, several included studies referred to North America, and particularly United States (n=2) and Canada (n=2) and one study referred to South America countries and particularly to Argentina, Brazil, Colombia, and Chile. The review also included one study referring to Asian countries (Bhutan, China, India, Indonesia, Japan, Malaysia, Philippines, Singapore, South Korea, Taiwan, Thailand). Last, but not least, Middle East and North Africa (n=1) as well as Saudi Arabia (n=1) were part of the final studies while international scope was referred in five studies.

**Table 1.**
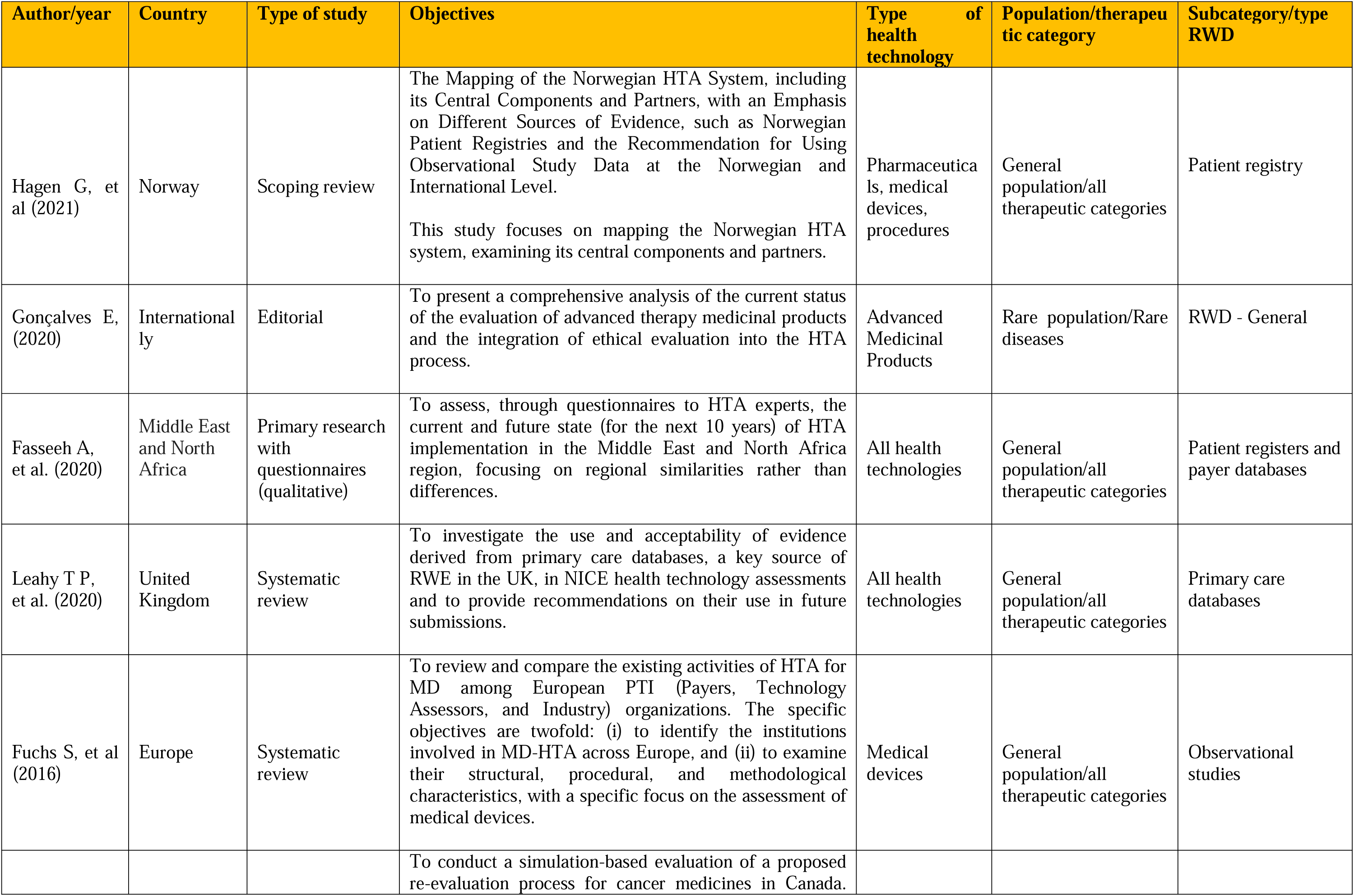
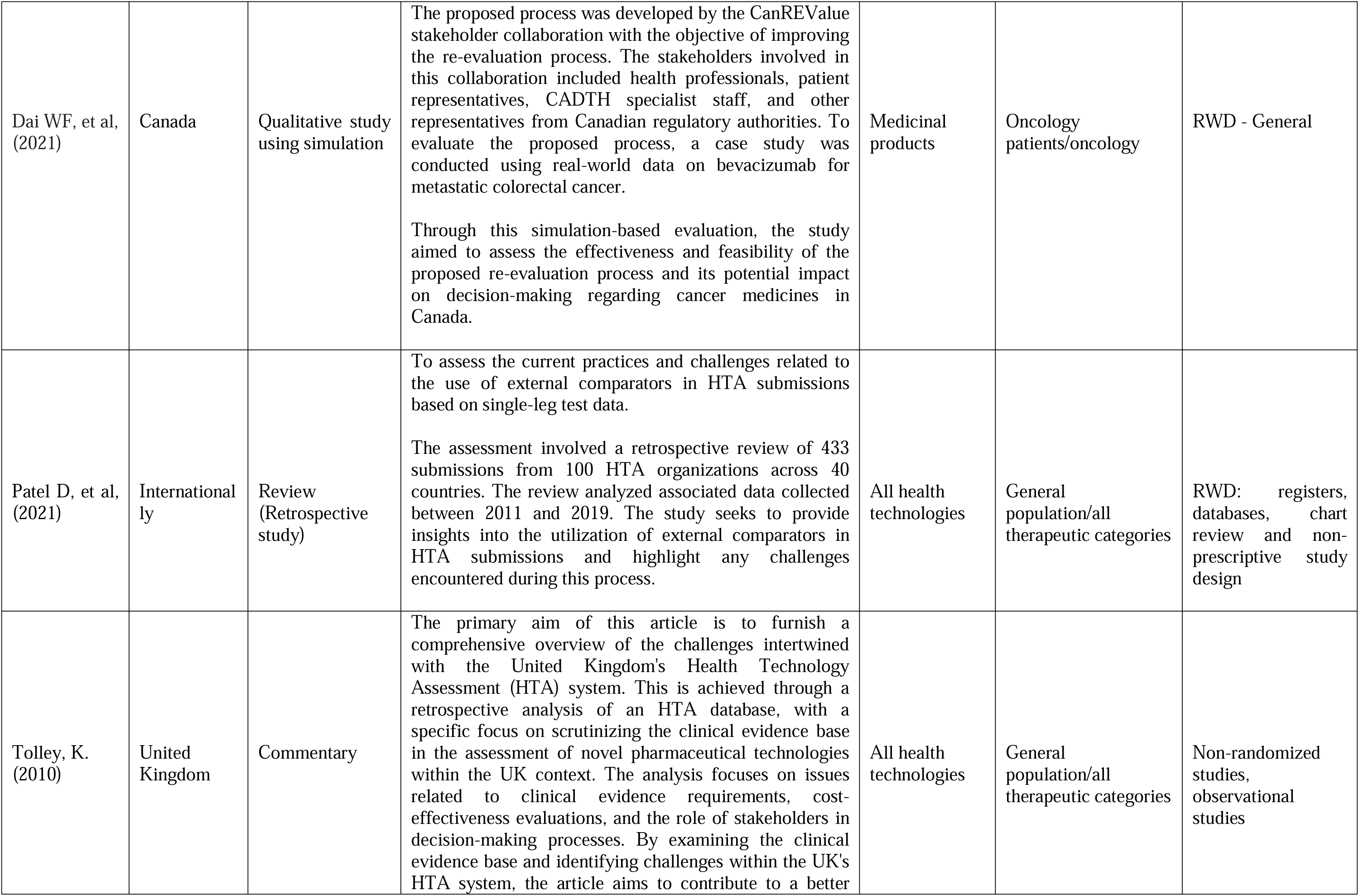
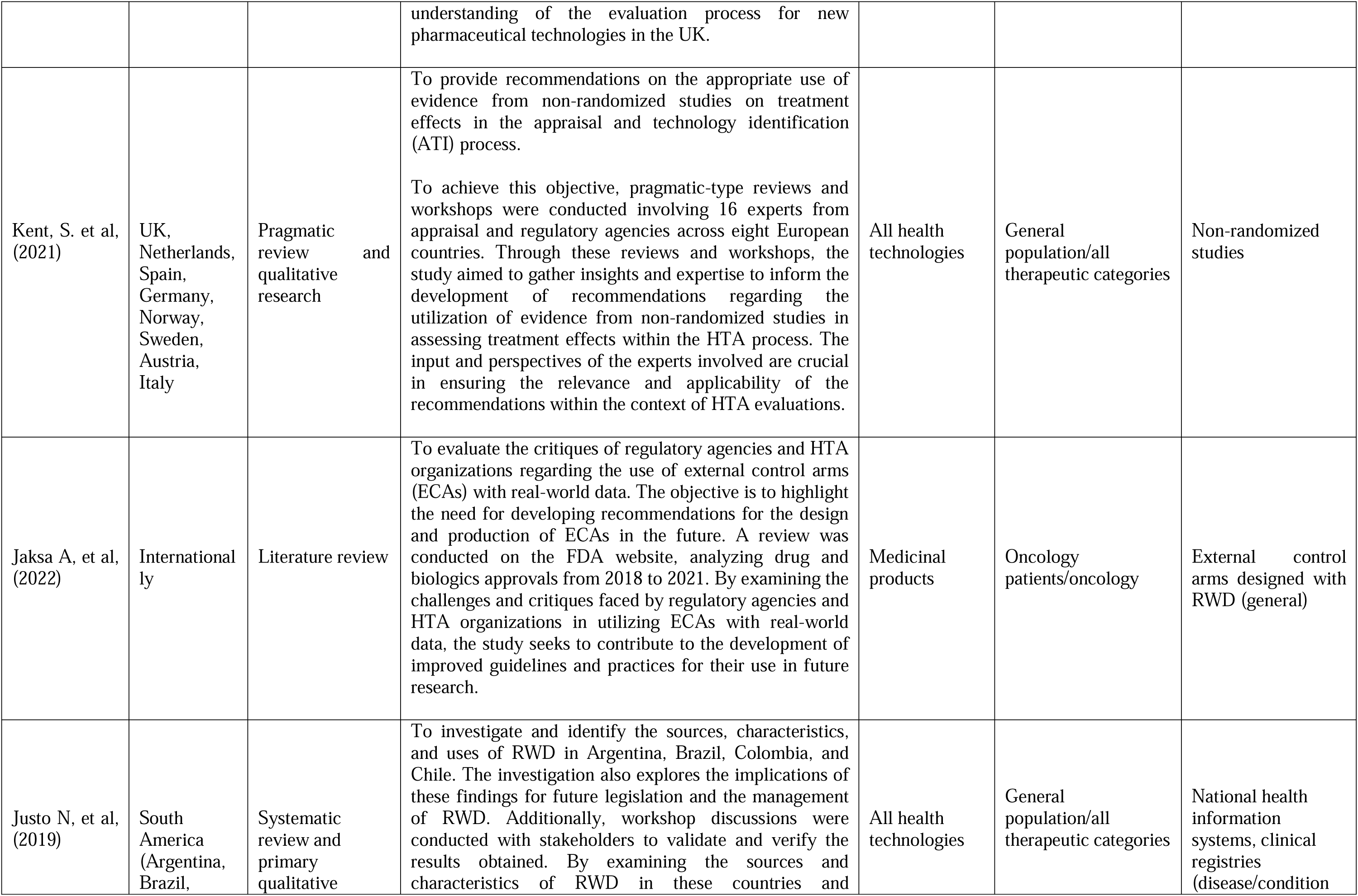
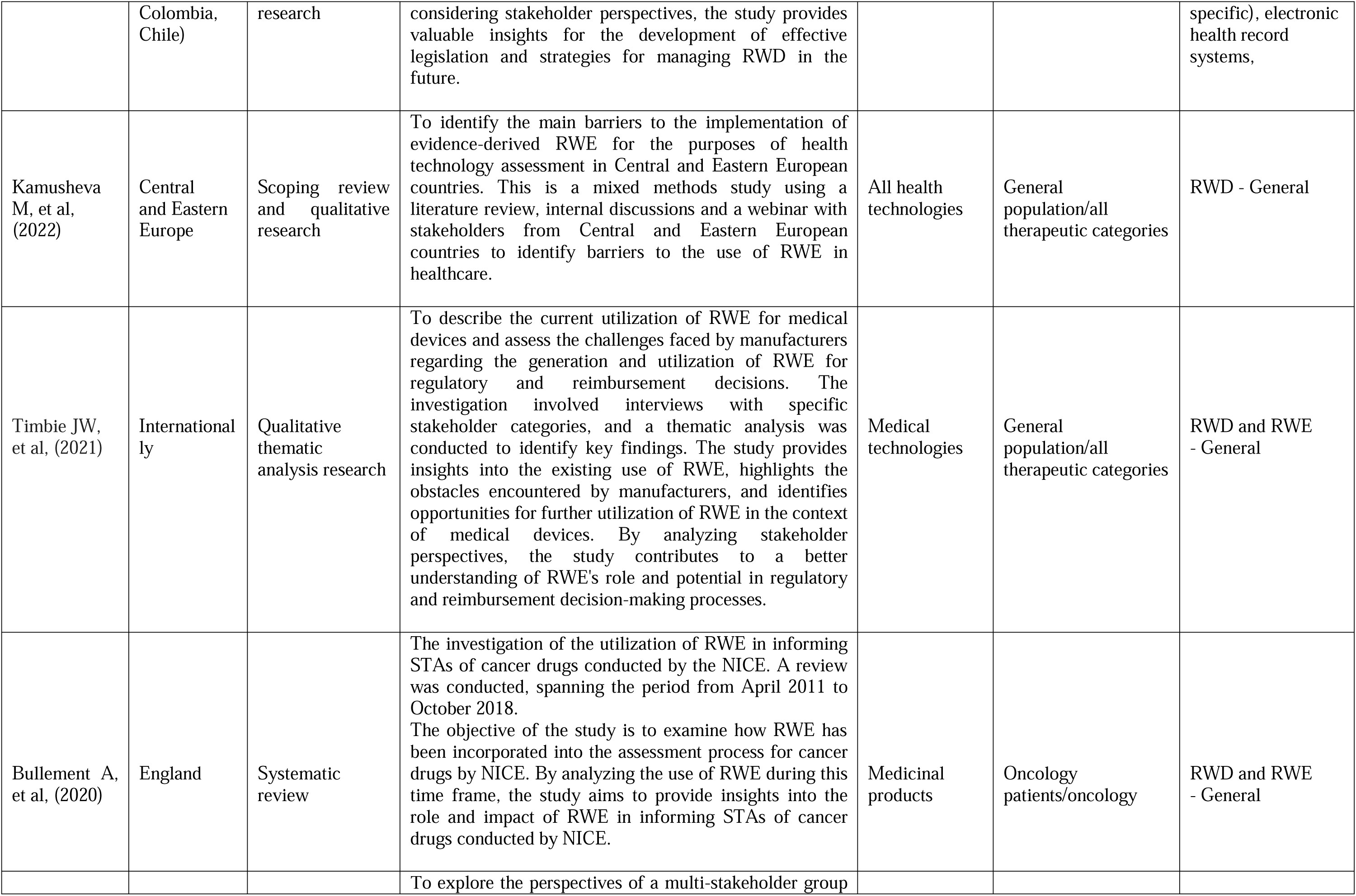
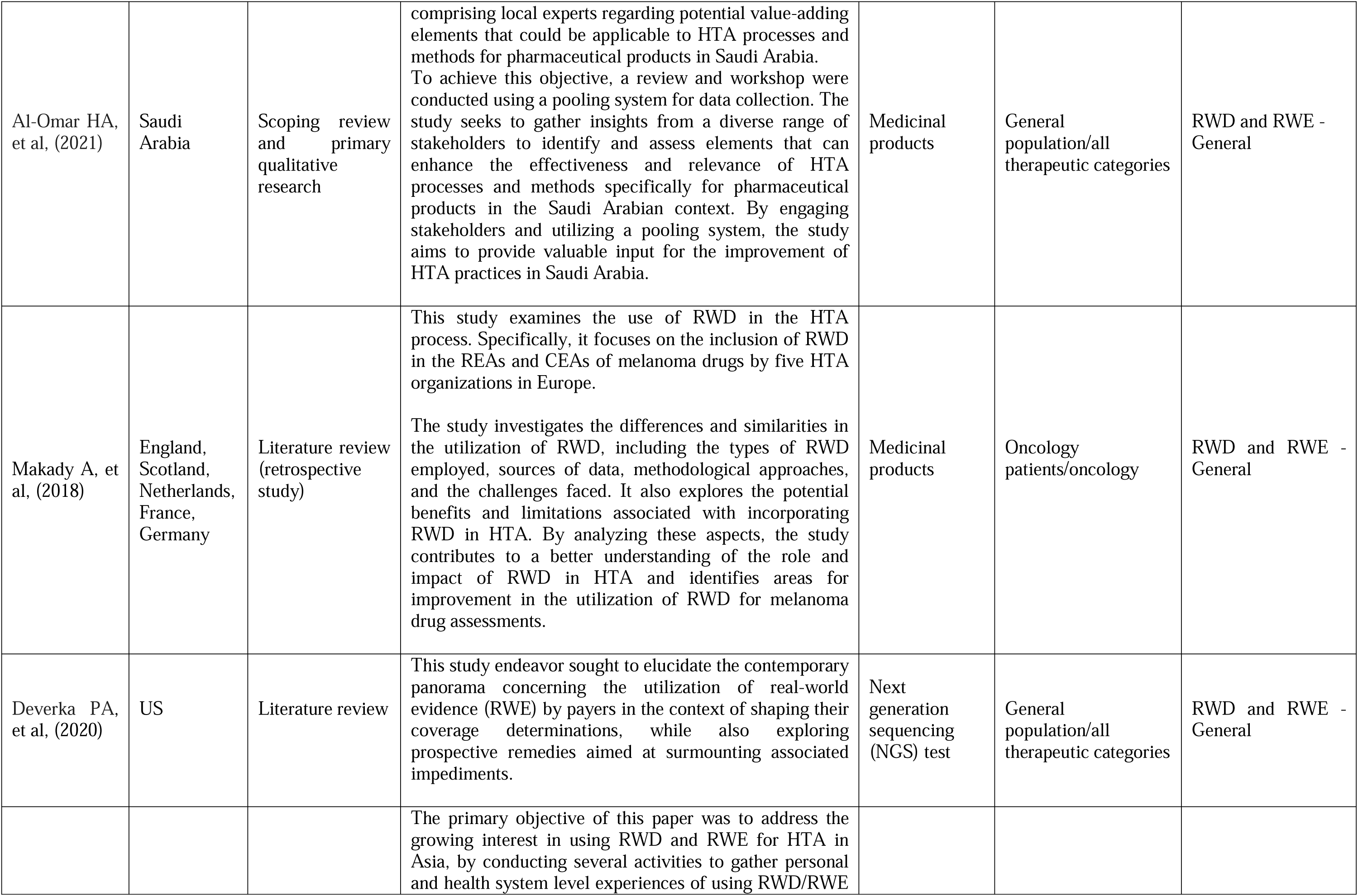
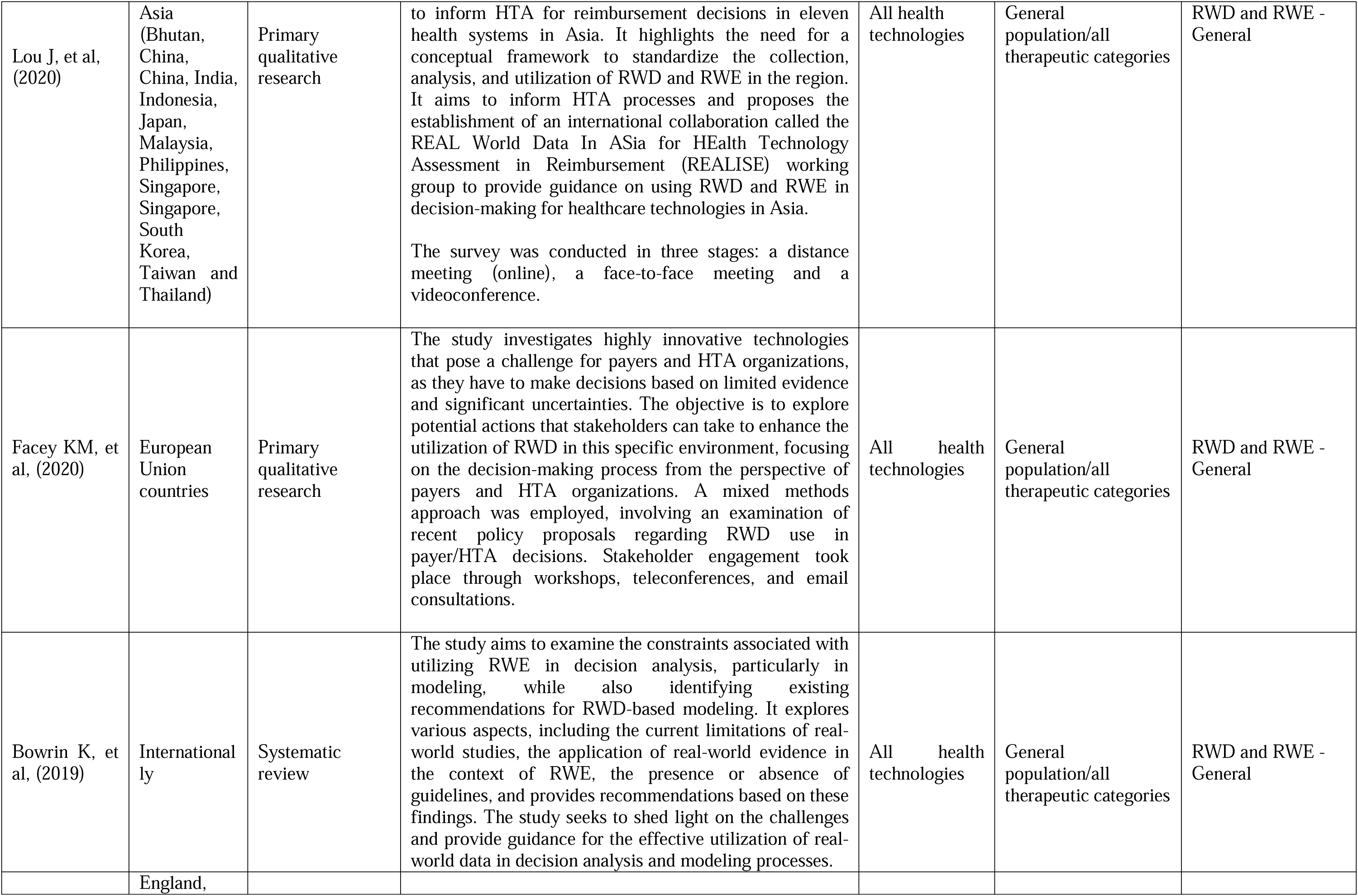
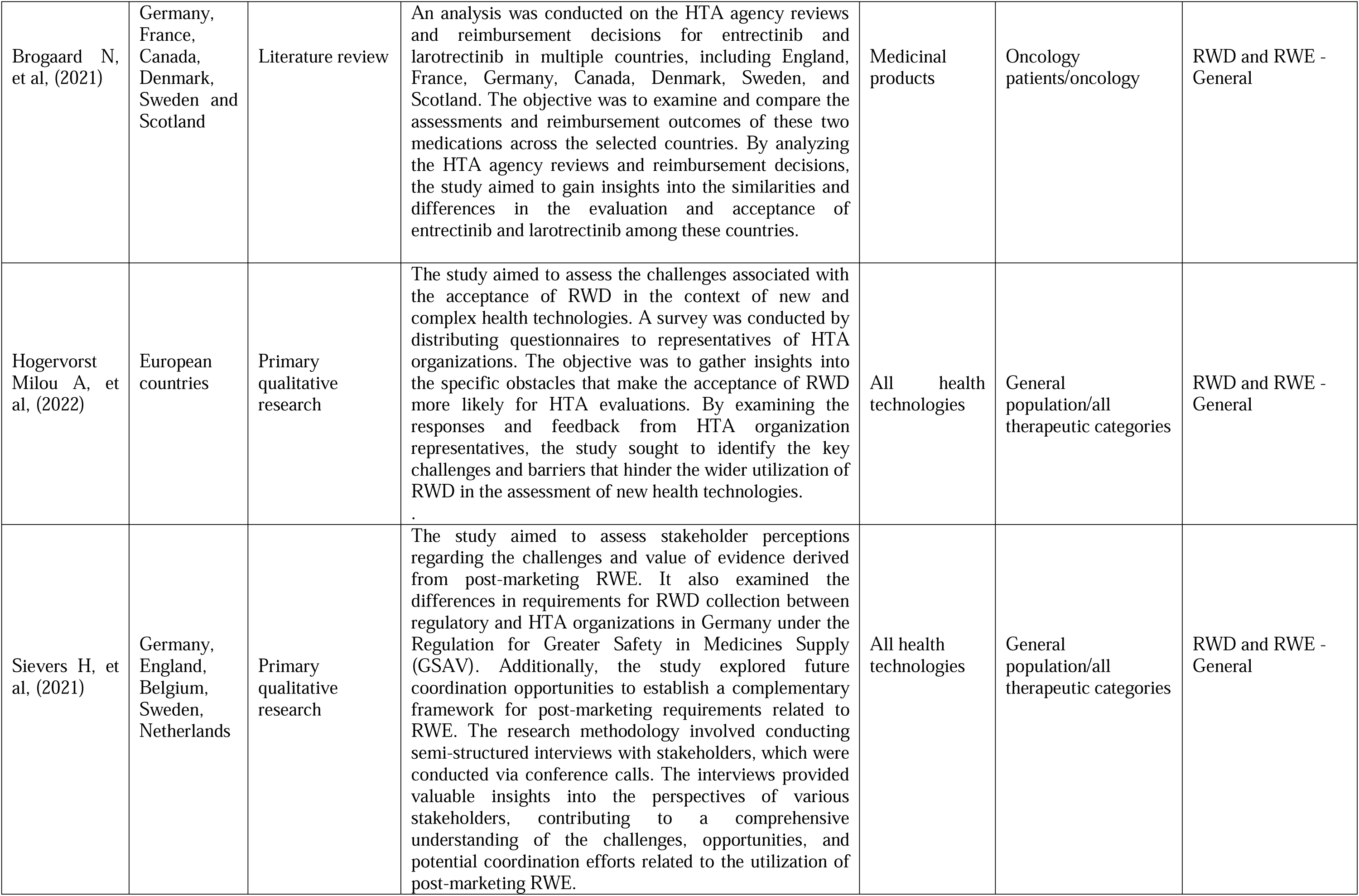
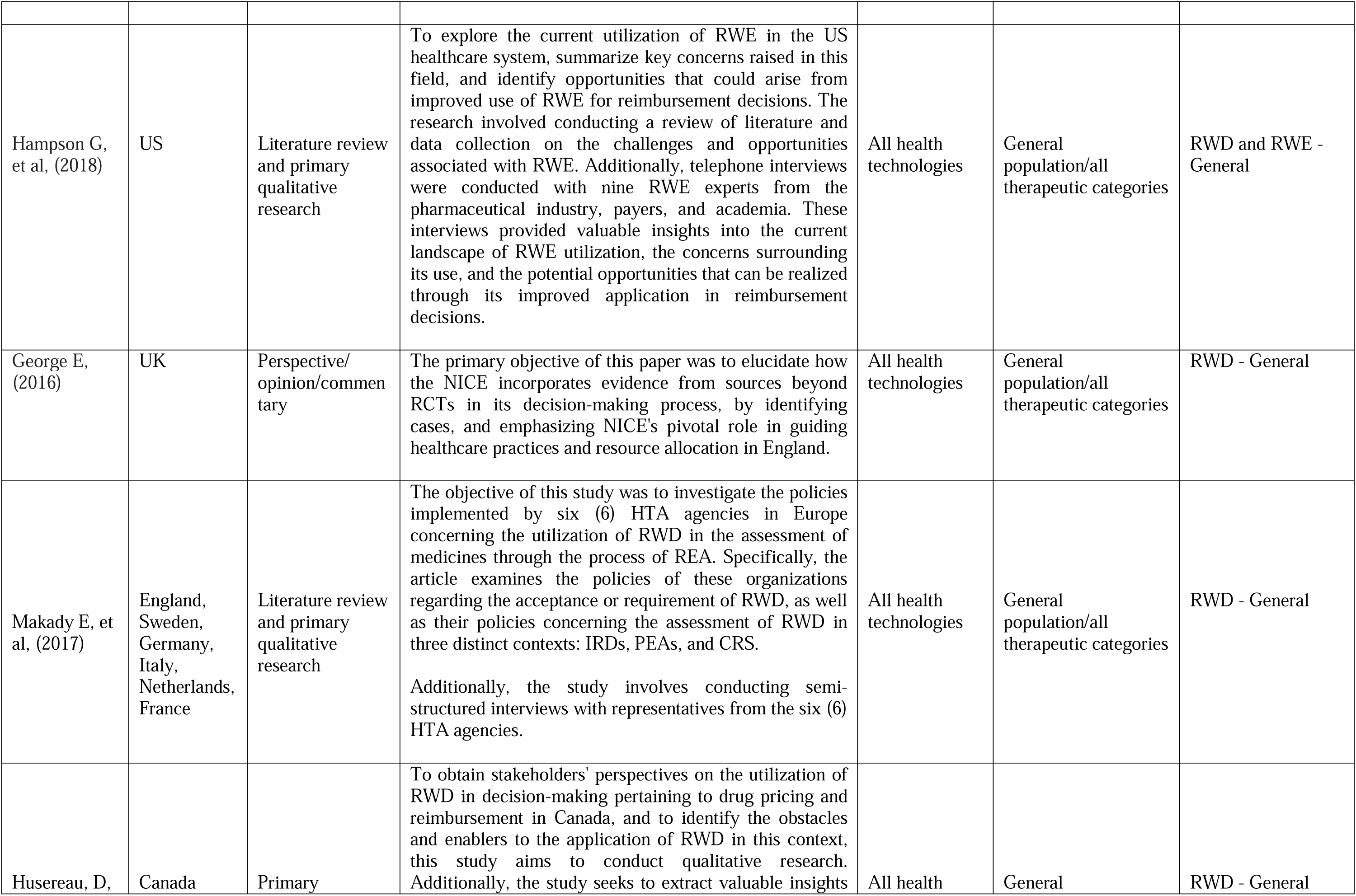
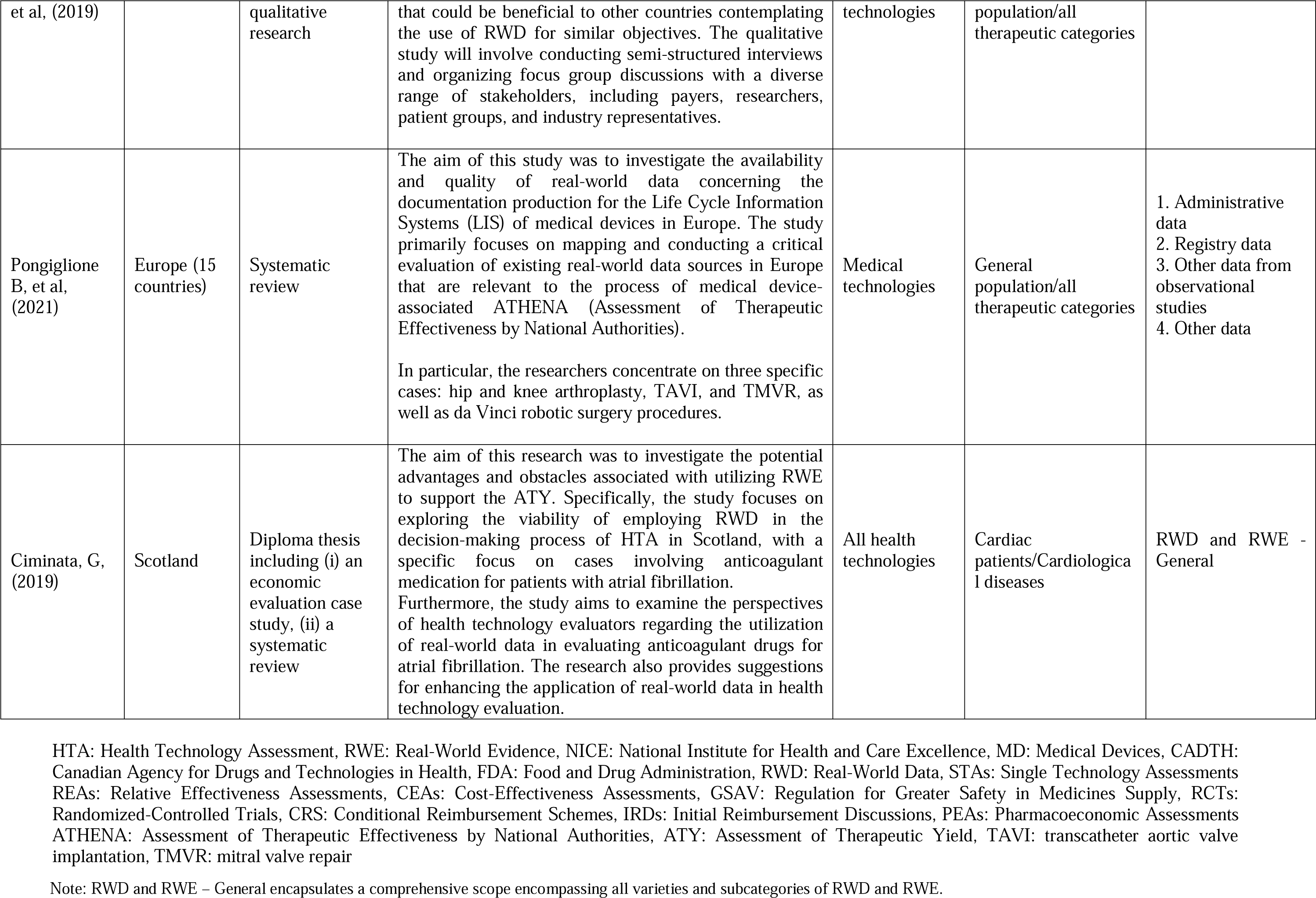
Characteristics of included studies.

### 3.2 Current use and acceptance of real-world data/real-world evidence in HTA

**Table 2.**
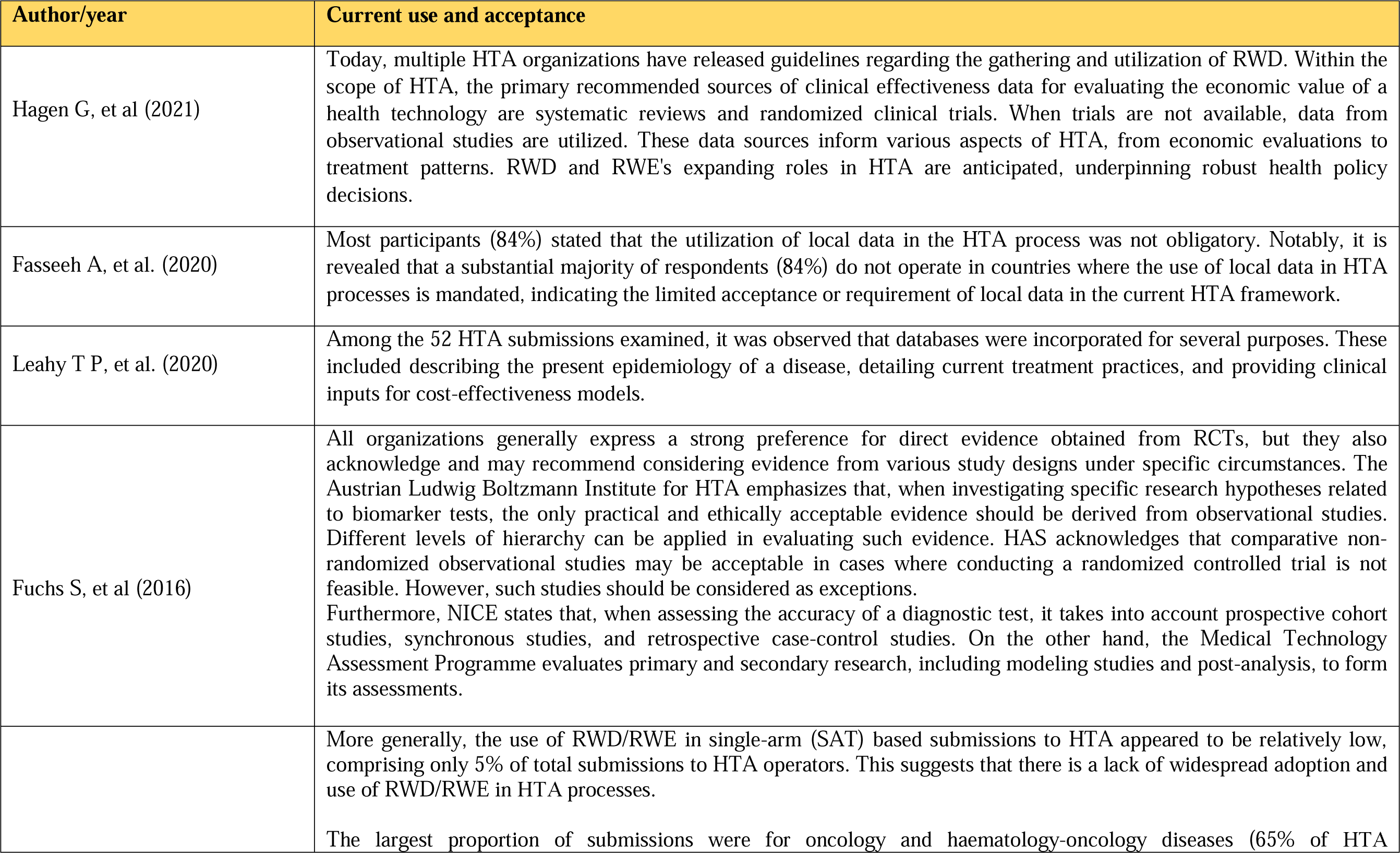
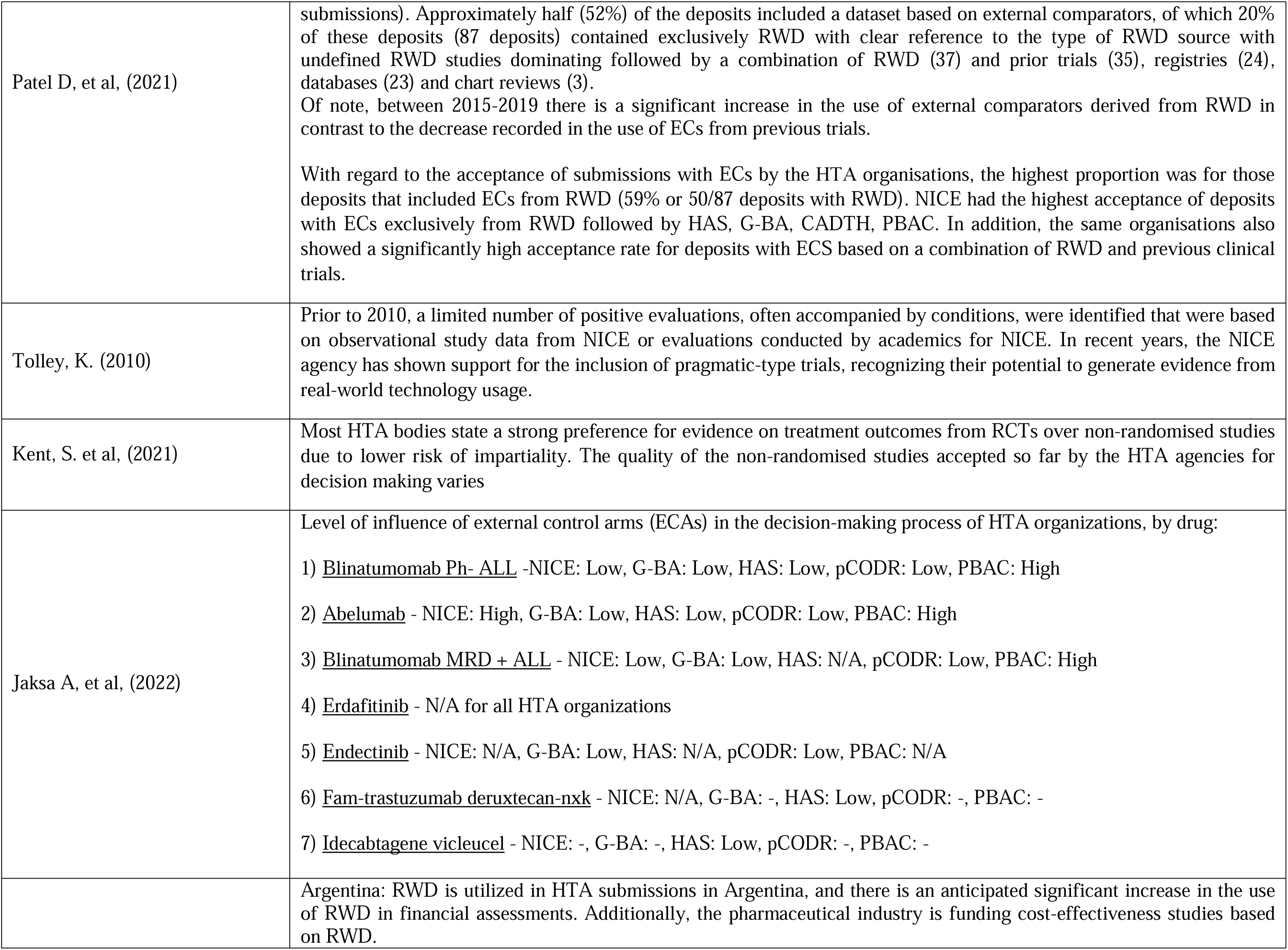
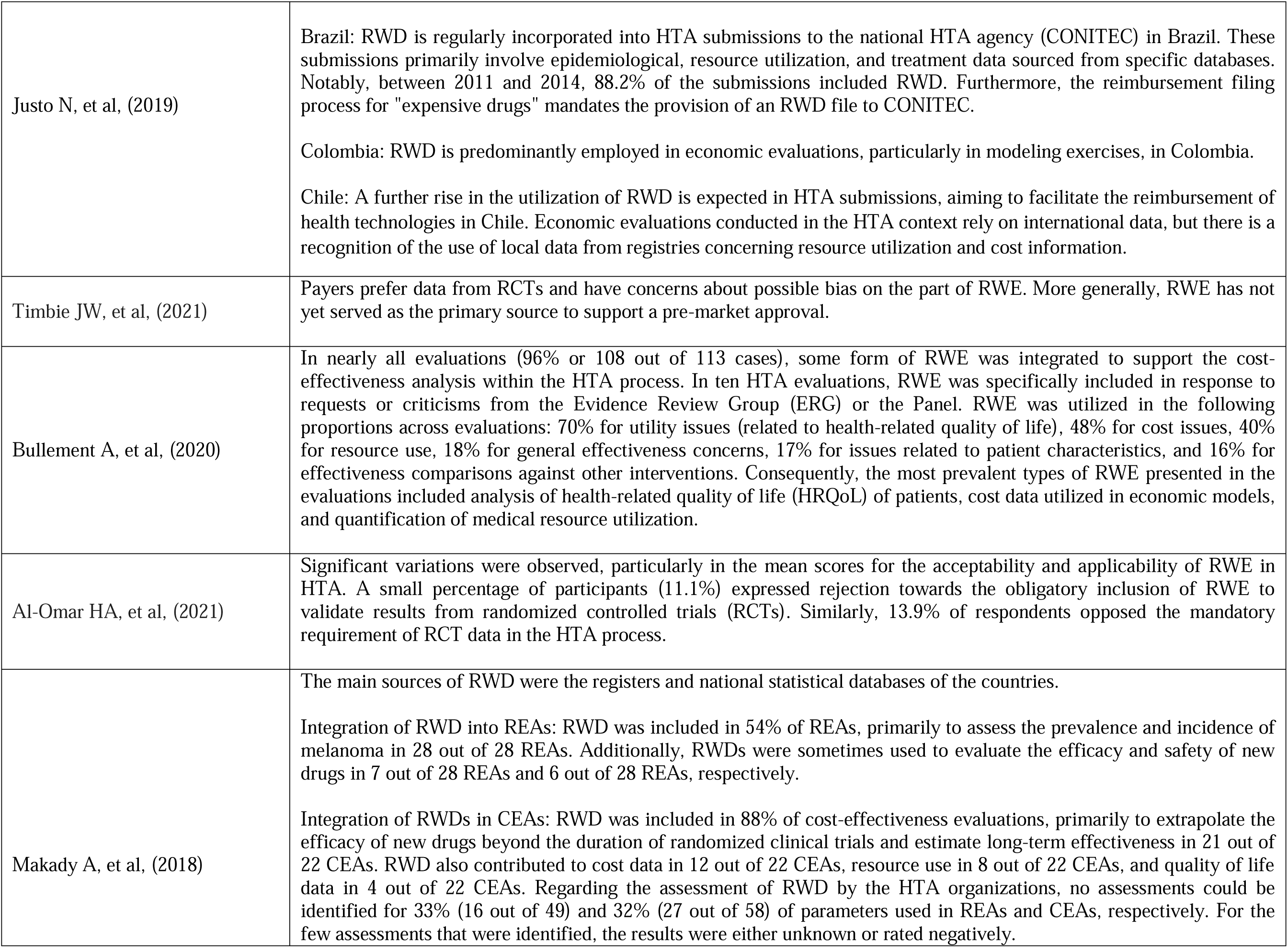
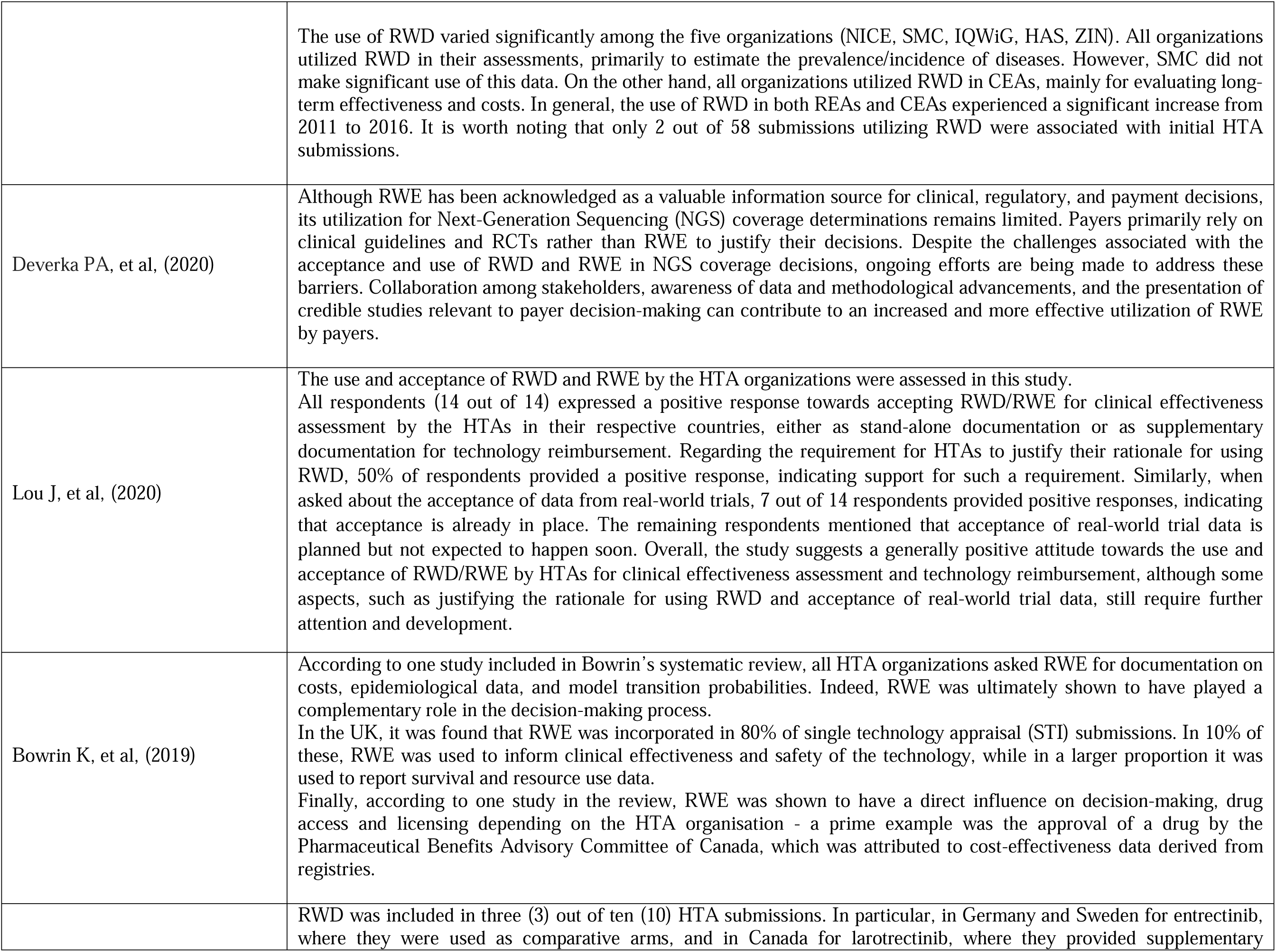
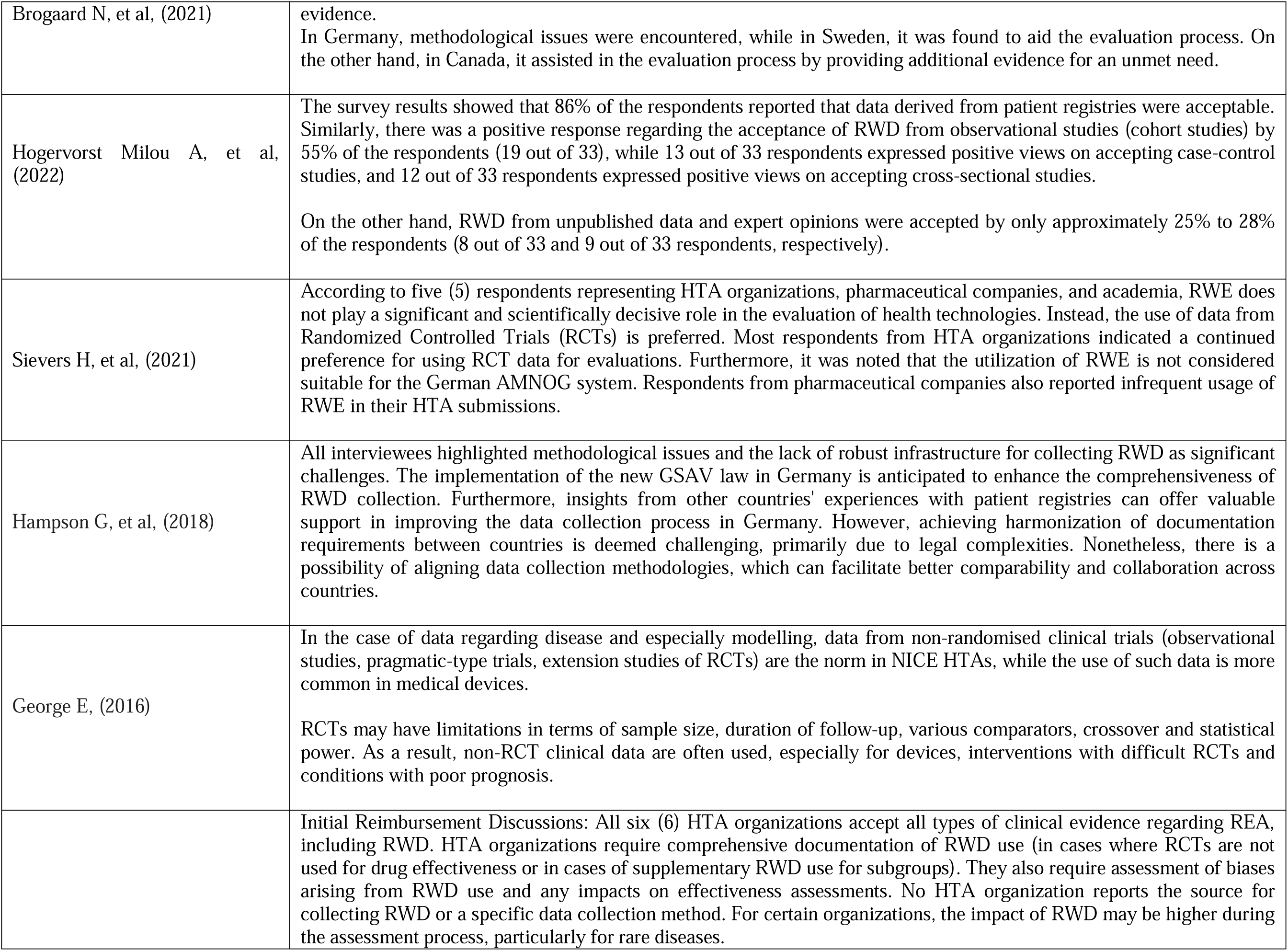
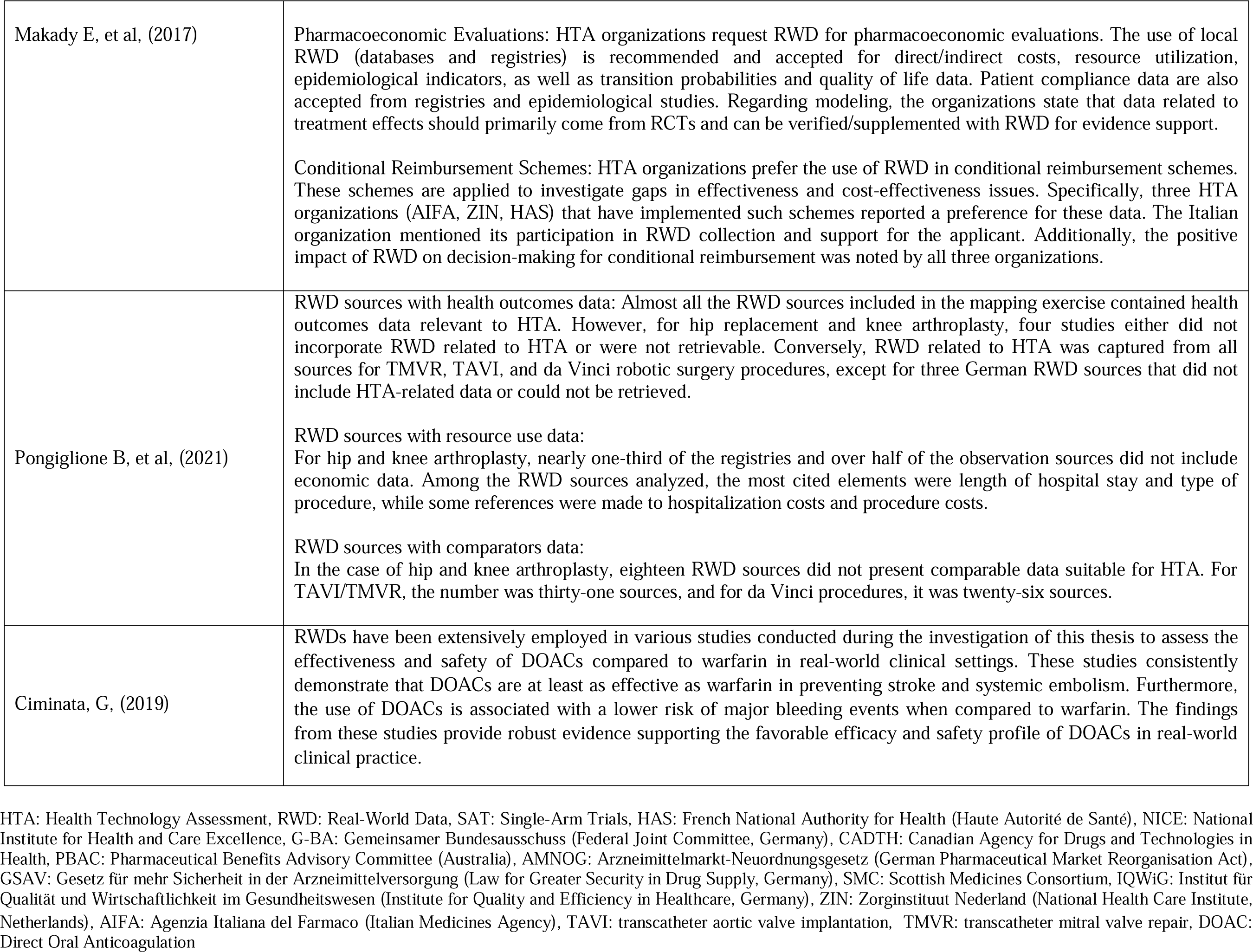
Current use and acceptance of RWD and RWE in HTA.

Out of the twenty-nine studies (n=29) reviewed, twenty-four studies (n=24) highlighted the current use or acceptance of RWD and/or RWE in the process of HTA. Hagen et al. (2021) [17] outlined the Norwegian HTA process and highlighted the use of RWD from observational studies when RCTs are lacking. Guidelines in Norway govern the collection and utilization of RWD, including data from Norwegian patient registries. Fasseeh et al. (2020) [18] found that the use of local data in HTA processes is not obligatory in the Middle East and North Africa region, suggesting that reliance on local data may not be a prerequisite in the region’s HTA processes. The utilization and acceptance of evidence from primary care databases in HTA conducted by the NICE was investigated by Leahy et al. (2020) [19]. These databases were used to depict disease epidemiology, describe treatments, and provide clinical inputs for cost-effectiveness models. Fuchs et al. (2016) [20] conducted a systematic review on the utilization of real-world data/evidence (RWD/RWE) in HTA for medical devices in European HTA organizations. The review found that while RCTs are preferred, alternative study designs such as observational studies are acknowledged in certain circumstances. On the other hand, Patel et al. (2021) [21] found that the use of RWD/RWE in single-arm based HTA submissions was relatively low, accounting for only 5% of total submissions. Oncological and hematological-oncological diseases were the most common areas of study. The use of external comparators derived from real-world data increased over time, with varying acceptance rates among HTA organizations. Furthermore, the clinical evidence used in the evaluation of new pharmaceutical technologies in the UK’s HTA system was analyzed by Tolley (2010) [22]. In particular, the study highlighted challenges related to evidence requirements, cost-effectiveness assessments, and stakeholder involvement. NICE emphasized the importance of including pragmatic trials for real-world documentation. Kent et al. (2021) [23] provided recommendations on the use of evidence from non-randomized studies on treatment effects in the HTA process. Most HTA bodies prefer RCT data due to lower bias risk, but there are variations in the acceptance of non-randomized trials among organizations. Furtheremore, the consideration and acceptance of external control arms (ECAs) using real-world data in regulatory and HTA evaluations for specific drugs in oncology was reviewed by Jaksa et al. (2022) [24]. Varying levels of acceptance were observed among different HTA organizations, including NICE, G-BA, HAS, pCODR, and PBAC. Additional included studies showed the significance of RWE in decision-making processes, variations in its utilization across different countries, and the prevailing preference for RCTs as a primary source of evidence in HTA. Justo et al.’s (2019) [25] systematic review and qualitative research across Argentina, Brazil, Colombia, and Chile explored RWD sources, attributes, and applications for health technologies. Findings indicate RWD’s growing role in Health Technology Assessments (HTA) and reimbursement, driven by anticipated increases and pharmaceutical industry investment. Colombia emphasizes RWD in economic evaluations, while Chile anticipates wider use for HTA and cost data. These insights inform future legislation and RWD management strategies. Timbie et al.’s (2021) [26] qualitative thematic analysis, conducted internationally, explores the contemporary application of RWE in the realm of medical devices. The research advances comprehension of RWE’s role in regulatory and reimbursement decision-making, despite prevailing payer preferences for RCTs and concerns about potential bias in RWE. Notably, RWE has yet to assume the principal role in pre-market approval processes. Bullement et al. (2020) [27] conducted a systematic review showing extensive incorporation of RWE in the cost-effectiveness analysis of cancer drugs evaluated by NICE in England, emphasizing its significant role in decision-making. Al-Omar et al. (2021) [28] identified differing stakeholder perspectives in Saudi Arabia regarding the mandatory integration of RWE and RCT data in HTA, indicating the need for further discussions in developing HTA processes for pharmaceuticals. The utilization of RWD in HTA was also observed by Makady et al. (2018) [29] and partocularly in melanoma medicine assessments across five European countries, noting variations in the types and extent of RWD incorporation, with an increasing trend over time. The importance of RWE in reimbursement decision-making process of specific technologies related to genomics was investigated by Deverka et al. (2020) [30]. They found limited utilization of RWE in coverage decision-making for next-generation sequencing (NGS) tests by US payers, with a primary reliance on clinical guidelines and RCTs. Lou et al. (2020) [31] reported positive acceptance of RWD and RWE for clinical efficacy assessment by HTA organizations in multiple Asian countries, but with varying opinions on the requirement to justify the rationale for using RWD. The frequent use of RWE in decision analysis and modeling for HTA was highlighted by Bowrin et al. (2019) [32], with RWE influencing various aspects of decision-making, drug access, and licensing. In addition, Brogaard et al. (2021) [33] observed the inclusion and utilization of RWD in HTA assessments and reimbursement decisions for entrectinib and larotrectinib, demonstrating its varying impact and benefits in different countries. Hogervorst et al.’s (2022) [34] primary qualitative research in European countries examined the acceptance challenges surrounding RWD for complex health technologies. Survey results indicated that patient registry data received high acceptance (86%), while unpublished data and expert opinions had lower acceptance rates (approximately 25% to 28%). Sievers et al. (2021) [35] conducted qualitative research on stakeholder perceptions of post-marketing RWE and RWD collection requirements in several European countries. The study highlighted a prevailing preference for RCT data and limited utilization of RWE in the evaluation of health technologies, particularly in the German context.

On the other hand, Hampson et al. (2018) [36] conducted research in the United States, finding that initial assessments of health technologies relied on data from RCTs by organizations such as the United States ICER and payers. The use of RWE for comparative clinical effectiveness posed challenges, but RWE was considered more relevant for re-evaluations by payers. Difficulties in collecting similar data hindered the widespread adoption of RWE. George (2016) [37] emphasized the common use of non-randomized clinical trial data in health technology assessments by the NICE in the United Kingdom. Non-randomized clinical trial data was valuable for assessing medical devices due to limitations in RCTs. NICE recognized the importance of including non-randomized clinical trial data to comprehensively evaluate health technologies. Makady et al. (2017) [38] examined the policies of six HTA organizations in Europe and found that they accepted various types of clinical evidence, including RWD, for initial reimbursement discussions. RWD was particularly requested for pharmacoeconomic assessments, and local RWD sources such as databases and registries were recommended. The use of RWD had a positive impact on the decision-making process. Pongiglione et al. (2021) [39] investigated RWD sources for HTA documentation on medical devices in Europe, focusing on procedures such as hip and knee arthroplasty, percutaneous aortic valve implantation (TAVI), mitral valve repair (TMVR), and da Vinci robotic surgery. Four categories of RWD sources were identified: administrative data, registry data, other observational studies, and other RWD sources with health outcomes data. Data availability varied across cases, and some sources lacked comparable data for HTA. Ciminata (2019) [40] explored the use of RWD to support HTA in Scotland, specifically for anticoagulant drugs in atrial fibrillation. RWD studies consistently showed that direct oral anticoagulants (DOACs) were at least as effective as warfarin in preventing stroke and systemic embolism, with a lower risk of major bleeding.

### 3.3 Barriers, challenges, and difficulties encountered in incorporating RWD/RWE within HTA

**Table 3.**
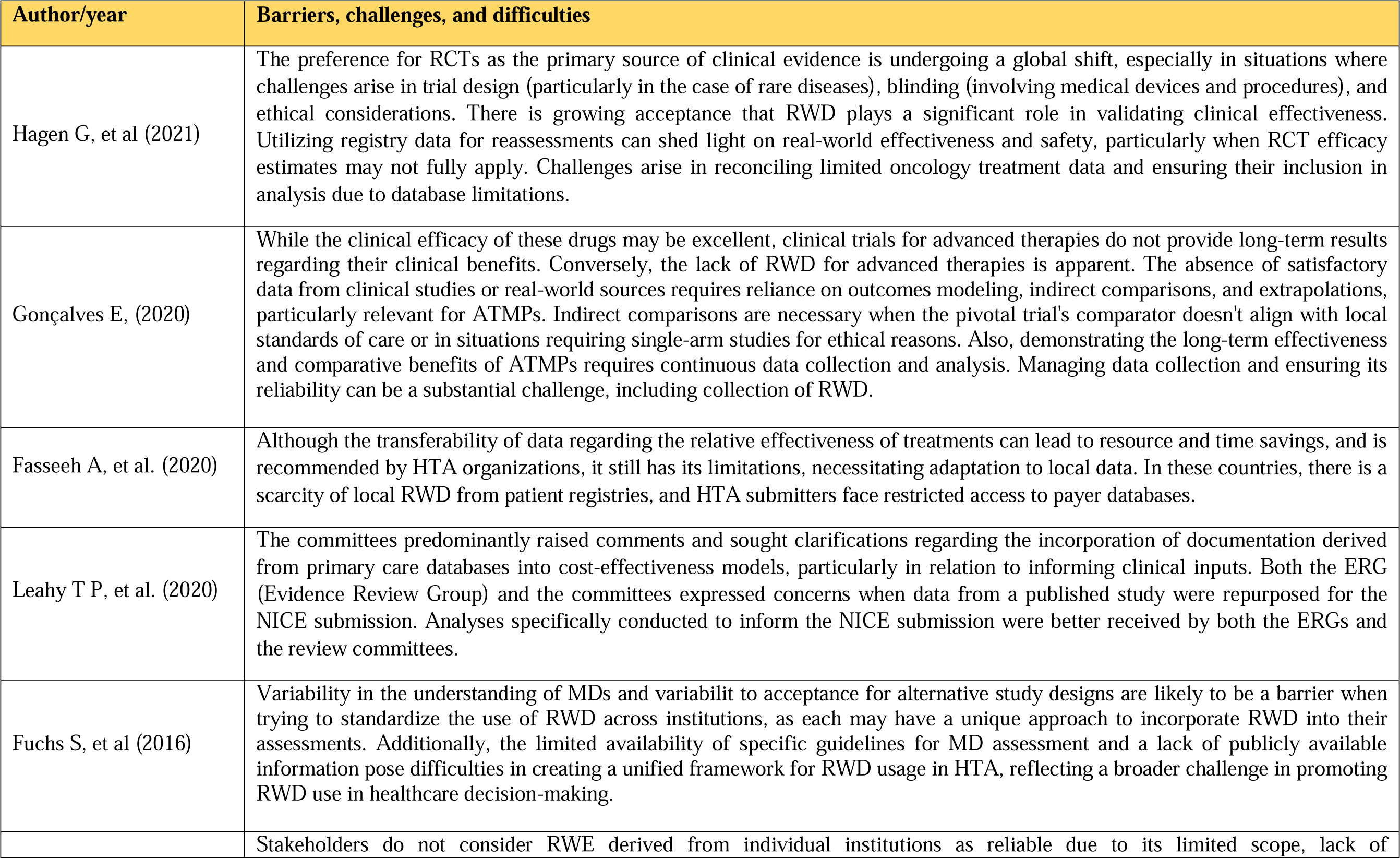
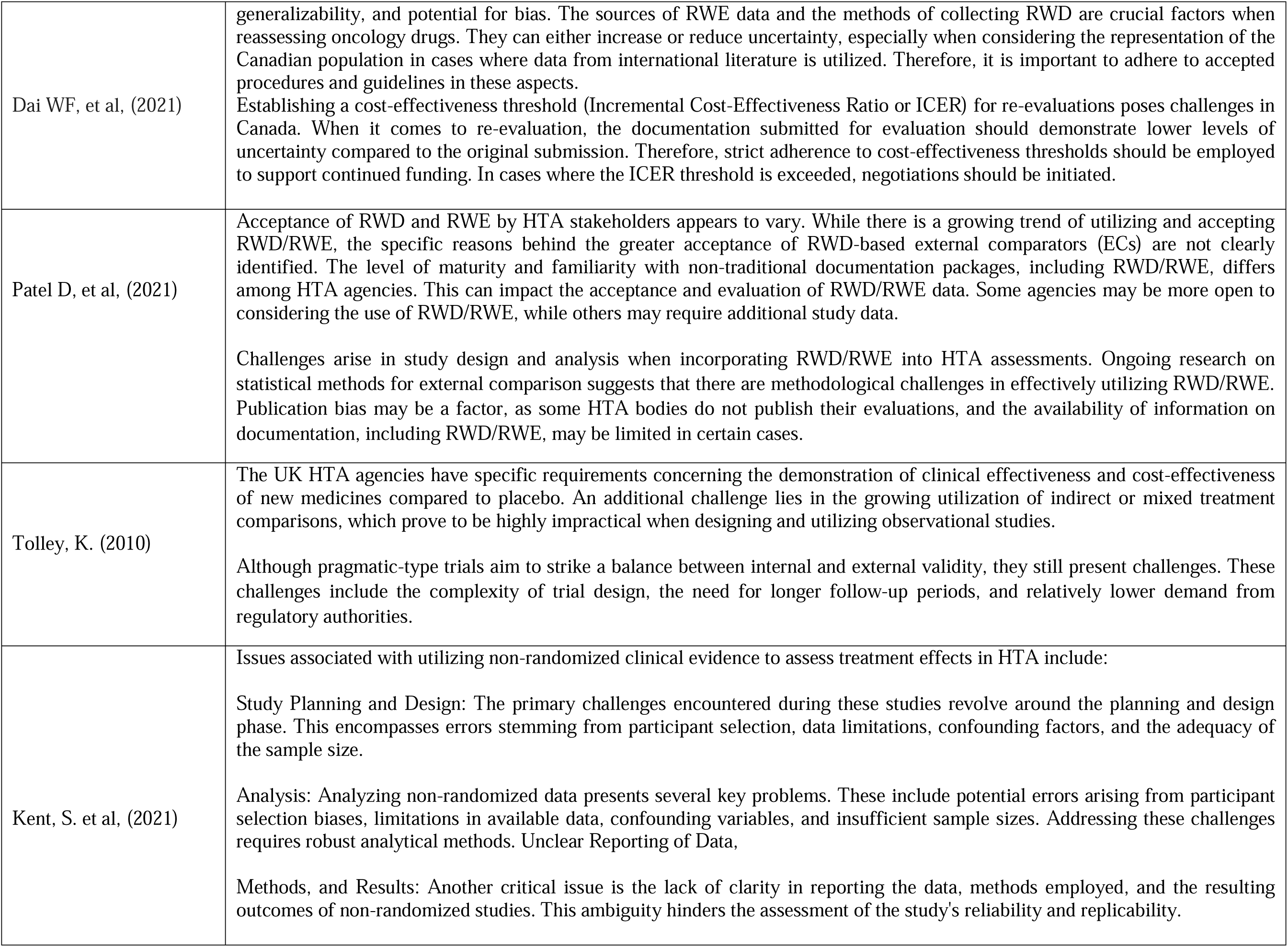
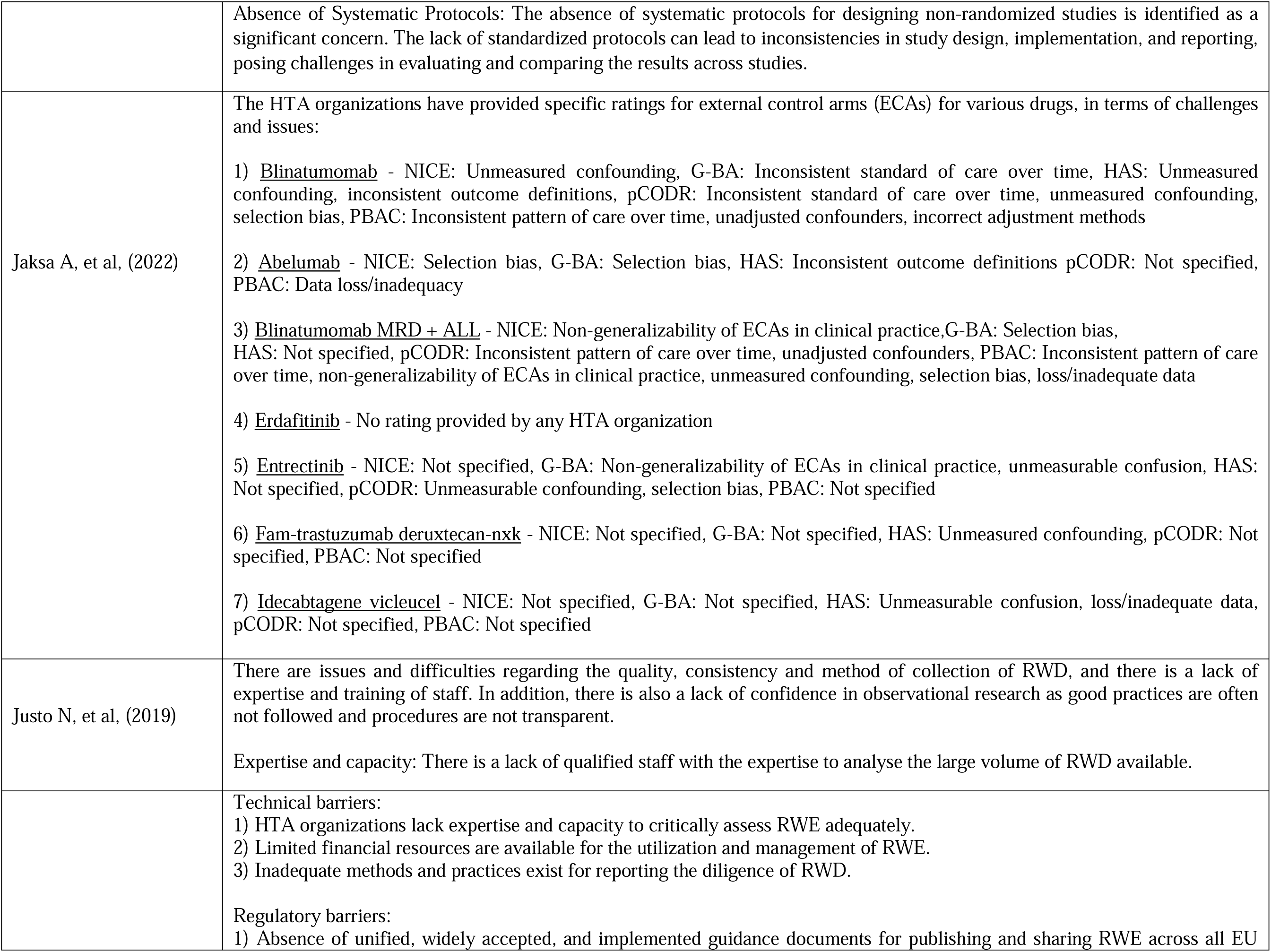
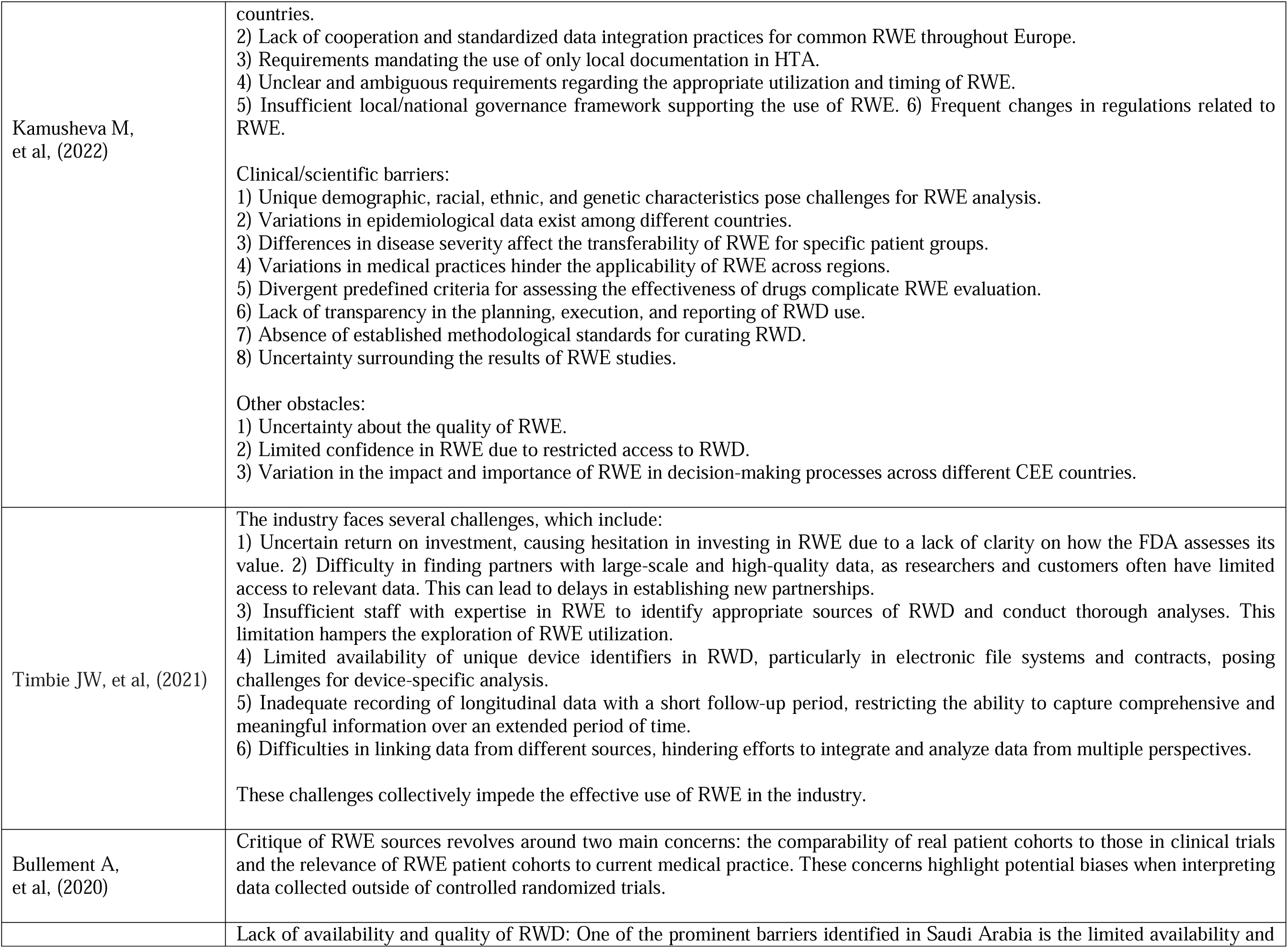
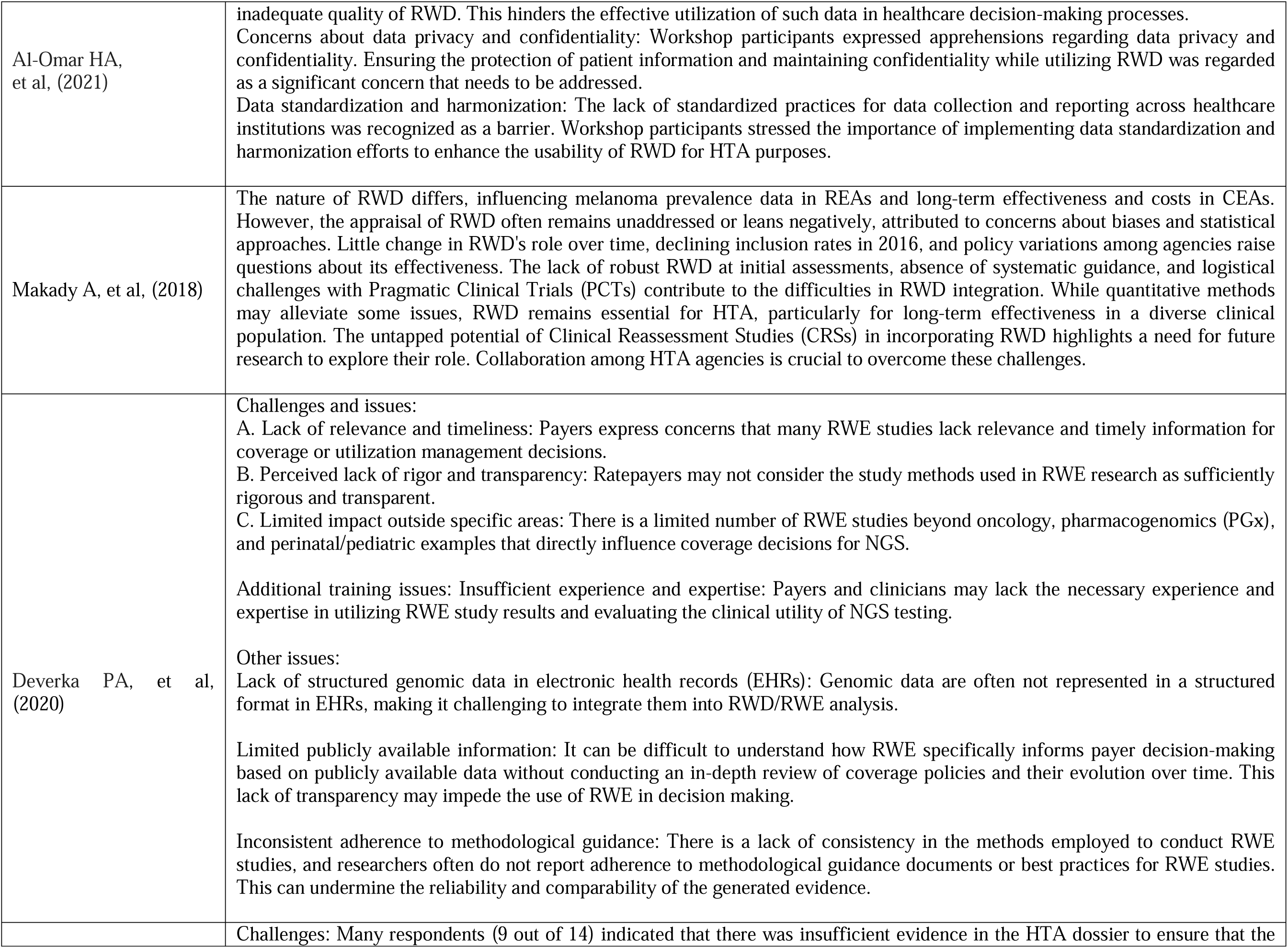
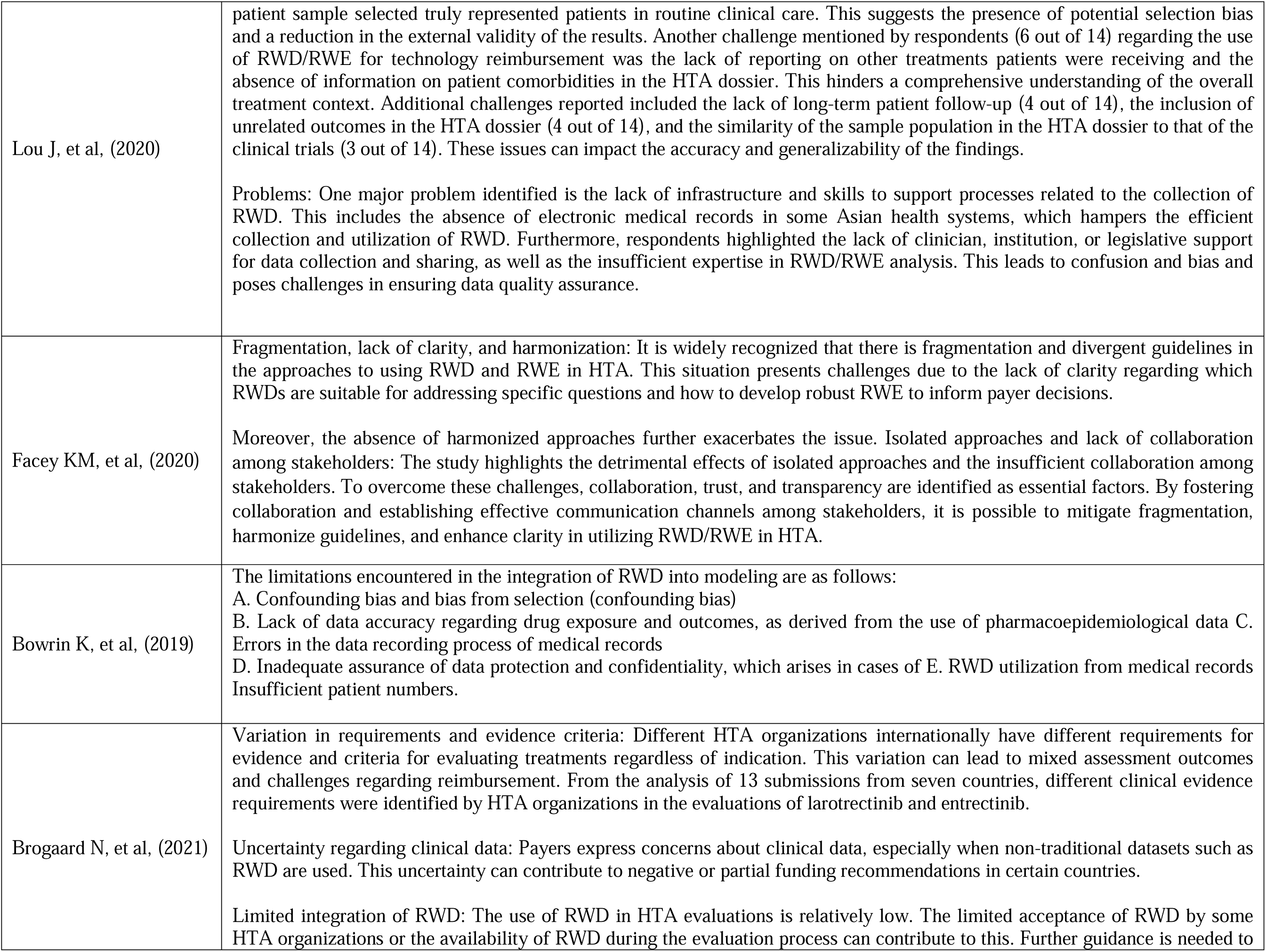
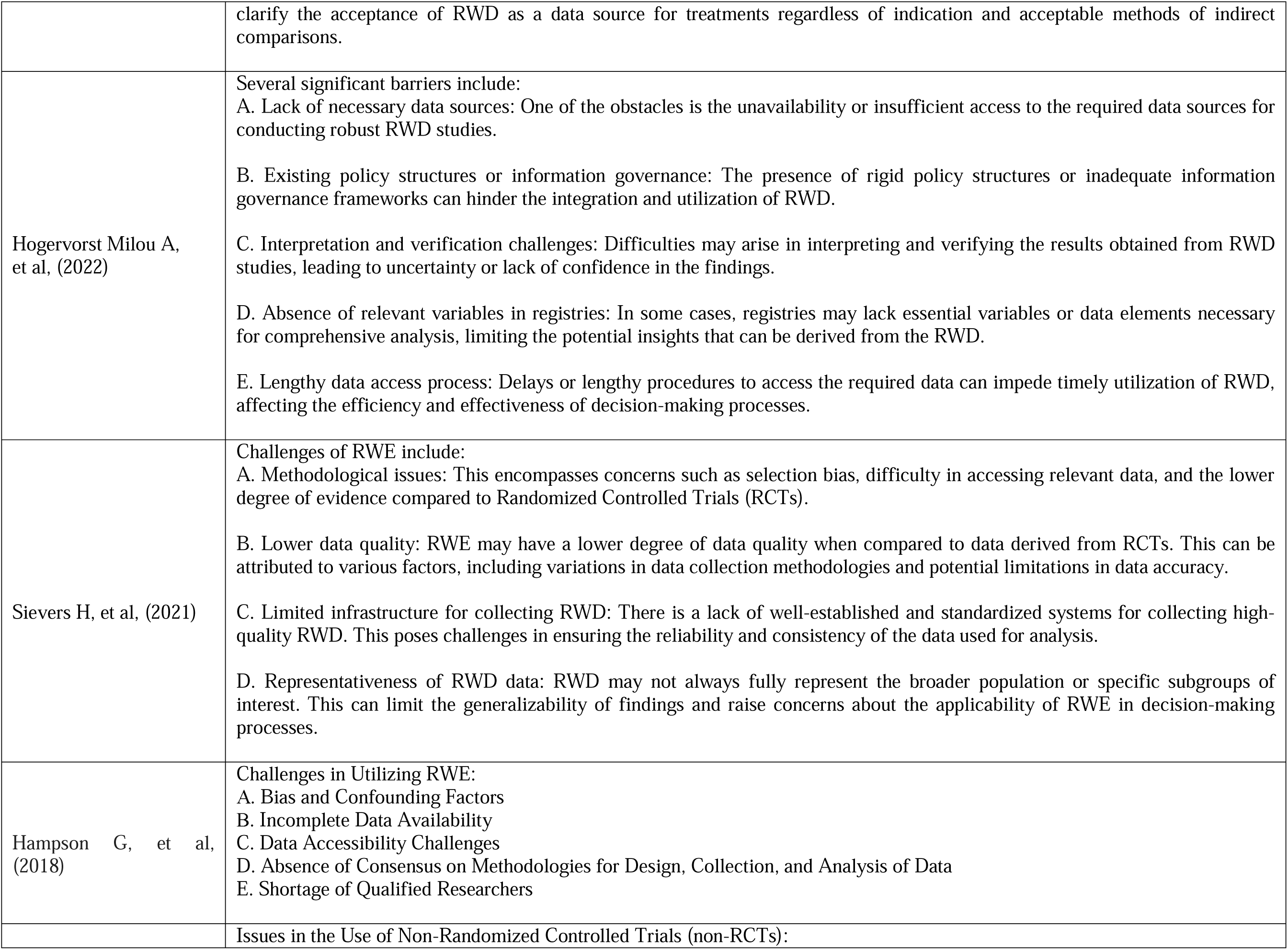
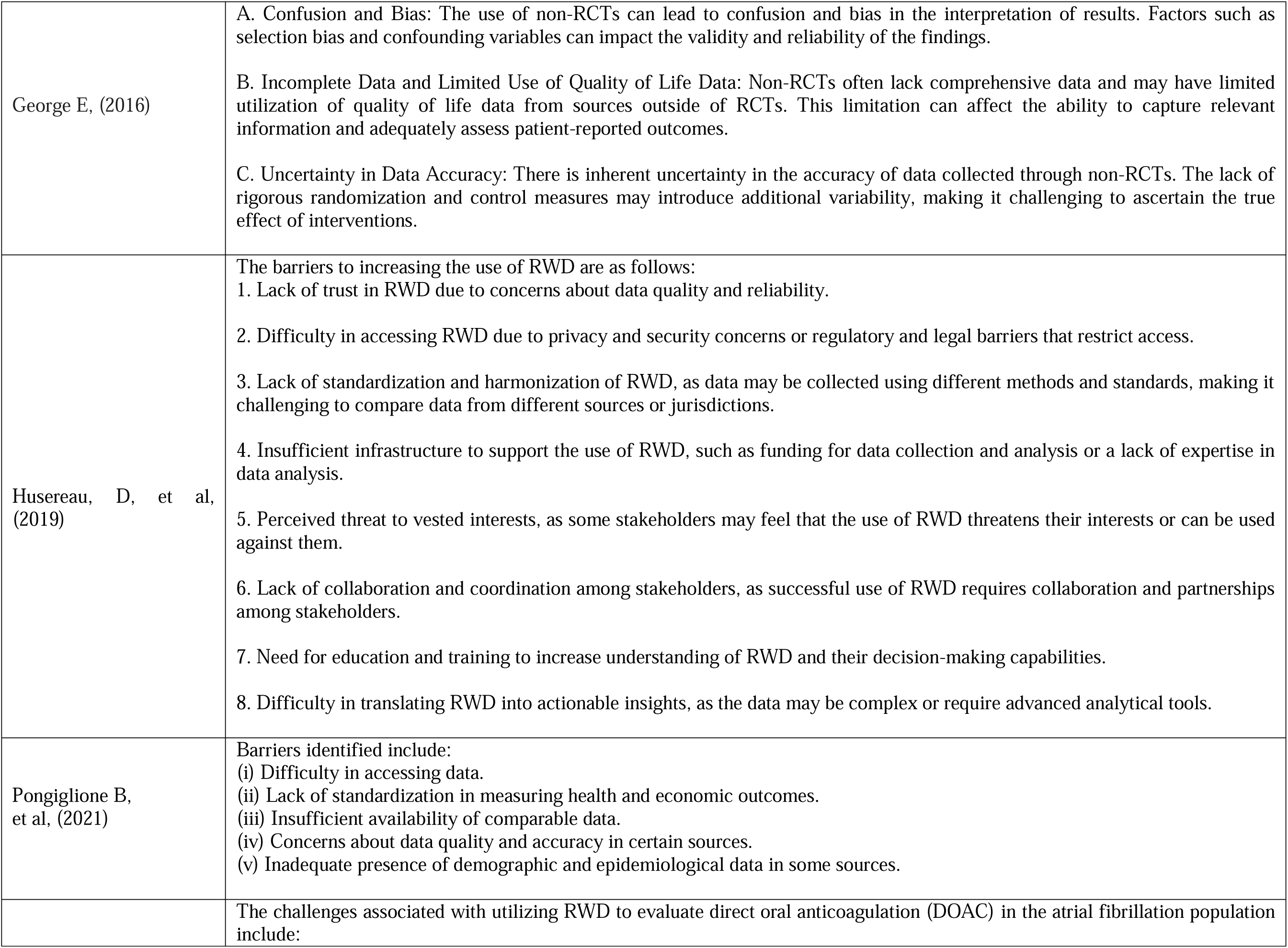
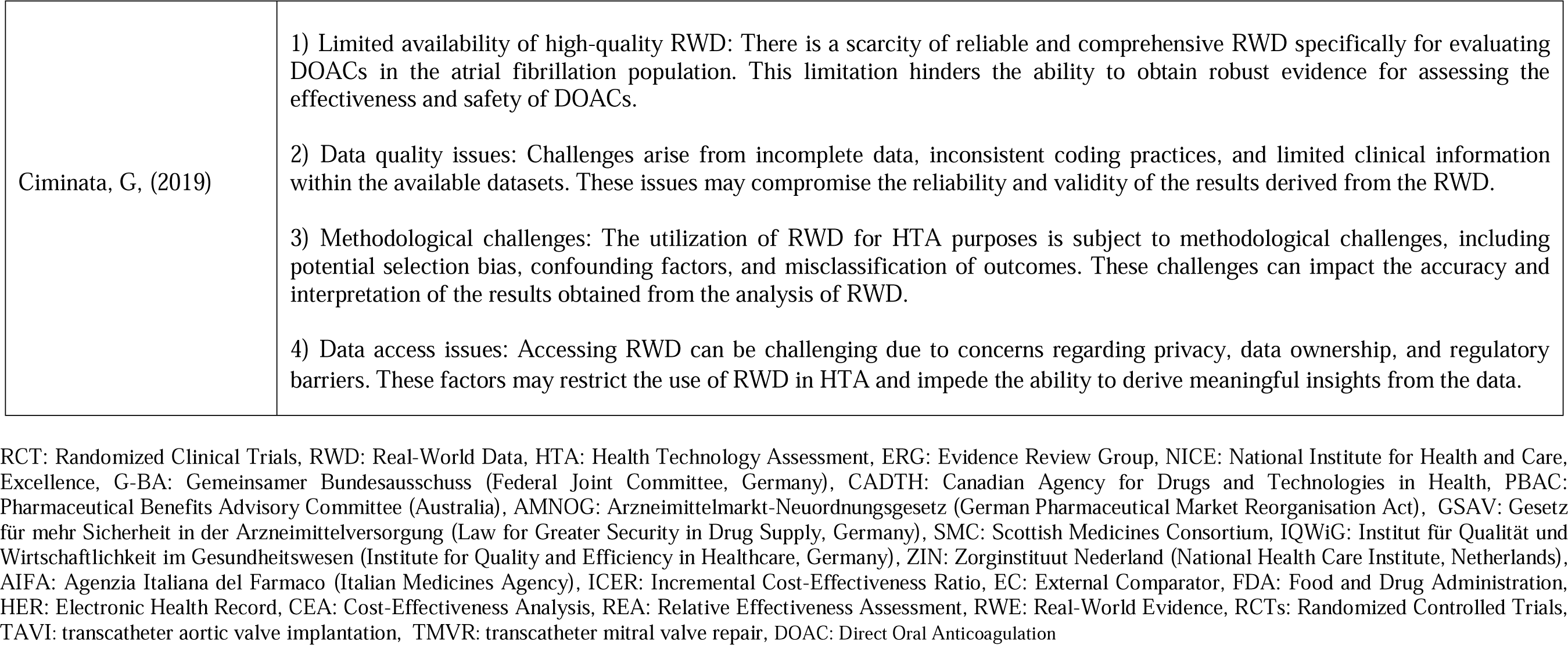
Barriers, challenges, and difficulties encountered in incorporating RWD/RWE within HTA.

The scoping review by Hagen et al. (2021) [17] emphasized the growing recognition of RWD from observational studies in Norwegian HTA. Real-world data contributes significantly to verifying clinical efficacy across different therapeutic categories within the general population. The editorial by Gonçalves (2020) [41] highlighted the importance of integrating ethical assessment into the HTA process for advanced therapy medicinal products (ATMPs). The inclusion of real-world data in HTA processes for ATMPs is crucial to provide a more comprehensive understanding of their real-world effectiveness but there is absence of satisfactory data from both clinical studies or real-world sources. The study by Fasseeh et al. (2020) [18] focused on HTA implementation in the Middle East and North Africa (MENA) region. The study revealed limitations in the availability and transferability of local real-world data, emphasizing the challenges faced in accessing and utilizing comprehensive local data sources for HTA processes in the region. The systematic review by Leahy et al. (2020) [19] investigated the use of documentation derived from primary care databases in health technology assessments conducted by NICE in the UK. Tailored analyses using data from primary care databases were better received by evaluation committees, highlighting the importance of generating specific analyses for NICE submissions. Another systematic review conducted by Fuchs et al. (2016) [20] focused on HTA activities for medical devices among European HTA organizations. The review highlighted the differences in the interpretation of MDs and variations in the willingness to embrace alternative study designs. This from another perspective could pose challenges in the pursuit of standardizing the integration of RWD across institutions. The qualitative study by Dai et al. (2021) [42] evaluated a proposed re-evaluation process for cancer drugs in Canada. The study emphasized the importance of reliable RWE, challenges in establishing an incremental cost-effectiveness ratio (ICER) for reassessments, and the importance of rigorous evaluation criteria in the re-evaluation process for cancer drugs. The retrospective review by Patel et al. (2021) [21] assessed the use of external comparators (ECs) based on single-arm test data in HTA submissions. Variability in the acceptance of RWD and RWE among HTA stakeholders was found, with challenges identified in study design, analysis, publication bias, and limited information. Tolley’s (2010) [22] commentary examined challenges within the UK’s HTA system, including clinical evidence requirements, cost-effectiveness evaluations, and stakeholder involvement. The commentary emphasized the specific requirements of UK HTA agencies for demonstrating clinical effectiveness and cost-effectiveness compared to placebo. Kent et al. (2021) [23] provided recommendations on the use of evidence from non-randomized trials in HTA. The study identified challenges in study planning, design, analysis, reporting, and systematic protocols in using non-randomized clinical evidence in HTA. Jaksa et al. (2022) [24] reviewed regulatory and HTA agencies’ critiques of external control arms (ECAs) with real-world data. The study emphasized the need for future recommendations in ECA design and production based on different critiques raised by HTA agencies. Justo et al. (2019) [25] explored RWD sources, characteristics, and uses in South America. The study identified problems with RWD quality, consistency, collection methods, lack of expertise, and confidence in observational research. Kamusheva et al. (2022) [43] conducted a scoping review and qualitative research in Central and Eastern European countries to identify barriers to implementing RWE in health technology assessment. The study found technical, regulatory, clinical, and scientific barriers, as well as other barriers, that hinder the adoption and integration of RWE into healthcare decision-making processes in the region. Timbie et al. (2021) [26] conducted a survey and interviews to examine the challenges faced by manufacturers in utilizing RWE for regulatory and reimbursement decisions in the medical device industry. The study identified barriers such as uncertain return on investment, difficulty in accessing high-quality data, and lack of RWE expertise, which hinder the widespread use of RWE. In addition, the systematic review conducted by Bullement et al. (2020) [27] on the use of RWE in single technology assessments (STAs) of cancer medicines by the NICE in England, found key criticisms regarding the sources of RWE, such as the comparability of real patient cohorts with clinical trial patients and the relevance of RWE patient cohorts to current clinical practice. Al-Omar et al. (2021) [28] conducted a scoping review and primary qualitative research in Saudi Arabia to explore the perspectives of local experts on potential evidence relevant to HTA processes and methods for pharmaceutical products. The study identified barriers such as limited availability and quality of real-world data, concerns about data privacy and confidentiality, and lack of standardization in data collection and reporting practices. What’s more, Makady et al. (2018) [29] conducted a retrospective study on the use of RWD in the HTA process for melanoma drugs in Europe. The study found that RWD inclusion was more common in CEAs compared to REAs, but there was a lack of consistent evaluation of RWD and variability across different HTA organizations. Deverka et al. (2020) [30] conducted a systematic review identifying barriers and issues related to the use of RWE in payer decision-making. The study found concerns about the relevance and timeliness of RWE studies, perceived inadequacy of RWE methods, limited influence of genomic studies on coverage decisions, and lack of expertise among payers and clinicians in using RWE. Lou et al. (2020) [31] conducted a qualitative study in multiple Asian countries on the use of RWD and RWE in all health technologies. The study identified challenges such as lack of representative patient samples, incomplete patient information, inadequate infrastructure and skills for RWD analysis, and lack of support for data sharing. Facey et al. (2020) [44] conducted a qualitative study in European Union countries to explore the use of RWD in decision-making processes for highly innovative technologies. The study identified challenges such as fragmentation, lack of clarity and harmonization, isolated approaches, and a lack of collaboration among payers and HTA organizations. An additional systematic review by Bowrin et al. (2019) [32] with the aim to explore the limitations of using RWE in decision analysis and modeling, identified limitations such as confounding bias, lack of accuracy in drug exposure and outcome data, challenges in data protection and confidentiality, and insufficient patient numbers. Brogaard et al.’s (2021) [33] analysis encompassed England, Germany, France, Canada, Denmark, Sweden, and Scotland, focusing on HTA agency reviews and reimbursement decisions for entrectinib and larotrectinib. The study aimed to compare assessments and reimbursement outcomes for these medications across countries, shedding light on similarities and differences in their evaluation and acceptance. Key findings included variation in evidence criteria, payer concerns about clinical data, especially non-traditional datasets like RWD, and limited integration of RWD in HTA evaluations, suggesting the need for guidance on RWD acceptance and indirect comparison methods. Hogervorst et al. (2022) [34] conducted a qualitative research study in European countries to assess challenges related to the acceptance of RWD in the context of complex health technologies. The study identified obstacles such as a lack of necessary data sources, inadequate policy structures, difficulties in interpretation and verification, absence of relevant variables, and time constraints in accessing data. Sievers, et al. (2021) [35] conducted a qualitative research study in multiple European countries to explore stakeholder perceptions of post-marketing RWE. The study identified challenges including methodological issues, low data quality, limited infrastructure for RWD collection, and ensuring data representativeness. Stakeholders emphasized the value of RWE but stressed the need to address these challenges for optimal use in decision-making processes. Hampson et al. (2018) [36] investigated RWE utilization by conducting a literature review and interviews with RWE experts revealed challenges: bias, incomplete data, accessibility issues, methodological disparities, and a shortage of qualified researchers. Enhancing RWE’s role in reimbursement decisions requires addressing these concerns and fostering consensus on methodologies. George E (2016) [37] provided a UK perspective on the use of non-RCT data within the NICE. The commentary highlighted challenges associated with non-RCT data, such as confusion, biases, incomplete data, limited use of quality-of-life data, and uncertainty regarding data accuracy. The discussion shed light on considerations for utilizing real-world data in evaluating health technologies across various therapeutic categories. Husereau et al. (2019) [45] conducted a qualitative research study in Canada on the use of RWD for drug pricing and reimbursement decisions. The study identified barriers to increasing RWD utilization, including lack of trust, difficulty in accessing data, lack of standardization, insufficient infrastructure, perceived threats to stakeholders’ interests, lack of cooperation, and challenges in translating data into action. Pongiglione et al.’s (2021) [39] systematic review encompassed 15 European countries, examining the availability and quality of real-world data pertinent to Life Cycle Information Systems (LIS) for medical devices. The study critically evaluated real-world data sources, emphasizing their relevance to medical device-associated ATHENA (Assessment of Therapeutic Effectiveness by National Authorities). Focused on hip and knee arthroplasty, TAVI, TMVR, and da Vinci robotic surgery, the research identified barriers, including data accessibility challenges, lack of outcome standardization, limited comparable data, data quality concerns, and demographic and epidemiological data gaps. The latest study by Ciminata (2019) [40] focusing on the use of RWE for assessing anticoagulant medications in patients with atrial fibrillation, explored the opportunities and challenges of using RWD in the decision-making process. It highlighted challenges such as limited availability of high-quality data, data quality issues, methodological challenges, and data access barriers. The thesis stressed the need for standardization in RWD collection, management, and analysis to improve the quality and comparability of results.

### 3.4 Potential Benefits, Opportunities, and Feasibility of Utilizing RWD/RWE in the HTA Process

**Table 4.** Potential Benefits, Opportunities, and Feasibility of Utilizing RWD/RWE in the HTA Process.

| Author/year | Potential Benefits, Opportunities, and Feasibility |
| --- | --- |
| Hagen G, et al (2021) | Efficiently structured registries can offer valuable data on health utilities, resource utilization, and costs related to the event/adverse action procedure of HTA. Patient registries play a crucial role in providing insightful information about current treatment patterns and adherence to treatment in real-world scenarios.<br>Patient registries offer the most comprehensive documentation of the natural progression of a disease based on local data and are recommended for consideration in economic evaluation modeling. Patient registries can serve as additional evidence to validate the real-world clinical effectiveness derived from clinical trials. This is especially relevant in the case of oncology medications that receive approval based on limited data, as registries can provide substantial contributions to the evidence base. There is a proposal to enhance collaboration between epidemiologists and HTA researchers, aiming to integrate data from population studies and registries more effectively. The data obtained from registries can significantly |
|  | contribute to health technology reassessments within the HTA process, and we can anticipate significant advancements in their utilization in the years to come. |
| Gonçalves E, (2020) | Collecting and analyzing RWD should complement the assessment of the long-term efficacy and comparative effectiveness of advanced therapies.<br>Highlighting outcome modeling can provide an important opportunity to address indirect comparisons between different treatments and extrapolate data, potentially bridging the gap caused by the absence of appropriate short-term clinical trial and RWE data. |
| Fasseeh A, et al. (2020) | A significant majority of respondents (82%) expressed their willingness to invest in patient registries and make payer databases accessible to HTA professionals. The endeavor to collect and utilize local data can serve as a valuable learning experience for HTA providers, particularly in countries that have made substantial investments in information technology infrastructure for healthcare financing and delivery. This can contribute to enhancing the quality of HTA assessments. In policy decision-making, there is a need to reinforce the significance of local documentation and local RWD, which can be achieved by extensively utilizing local patient registries and payer databases. |
| Leahy T P, et al. (2020) | Primary care databases are playing an increasingly important role in informing NICE submissions and showcasing the utilization of RWE in healthcare decision-making. The inclusion of primary care databases to inform clinical inputs in cost-effectiveness (CE) models is well received by both the Evidence Review Group (ERG) and the review committees, particularly when a dedicated study is conducted to extract these inputs. The absence of extensive commentary observed when databases were utilized to inform current treatment practices may indicate a general acceptance that databases serve as reliable sources of such data. The potential of UK primary care databases in NICE submissions is being increasingly recognized and utilized, particularly in informing the parameters of economic models. |
| Fuchs S, et al (2016) | In response to these challenges, several initiatives have been undertaken to tackle them. These initiatives include the implementation of standardized data elements and outcome measures, the advancement of analytical methods and tools for analyzing RWE, and the establishment of data exchange platforms and networks. RWD and evidence from non-randomized controlled trials (non-RCTs) are progressively gaining recognition for HTA of medical devices in Europe. |
| Dai WF, et al, (2021) | The participating stakeholders acknowledged that the availability of evidence generated through the utilization of RWE on clinical benefit and cost-effectiveness from various provinces in the country instills confidence in the results. They particularly emphasized the usefulness of this evidence in the context of the re-evaluation process, which aims to support policy decisions. When considering the use of RWE from other jurisdictions or districts in decision support, it should only be considered as an option if the data collection adheres to accepted standards and potential biases are effectively managed by experts in survey methodology. However, participants expressed that the use of RWE from other jurisdictions is more acceptable in cases involving rare diseases, as it can be challenging to adequately represent the population solely based on Canadian data.<br>Another noteworthy aspect highlighted was the inclusion of patient experience in the reporting of documentation. In other words, patients should be actively involved in the reassessment processes by providing data on their experience through questionnaires, for instance. Additionally, patient-generated outcomes (PROs) can make a valuable contribution to the |
|  | desired evidence. It was also suggested to engage the same patients who participated in the initial evaluation when conducting the re-evaluation of oncology drugs. |
| Patel D, et al, (2021) | The evaluation of evidence-based submissions for HTA based on single-arm trials lacks formal guidance and established best practices, making it challenging to construct comparison cohorts. Therefore, it is necessary to develop a well-designed strategy when relying on single-arm trial data, incorporating robust methods for data collection and a well-executed analytical approach for indirect treatment comparison.<br>Monitoring and guideline development: The increasing utilization of RWD in HTA submissions, albeit still relatively low (5% of all submissions), indicates a growing trend. It is important to closely monitor this category of submissions, and the study can assist HTA providers and payers in developing guidelines and policies for effectively handling such submissions.<br>Oncology, rare diseases, and orphan drugs: Submissions for orphan drugs have a higher acceptance rate compared to non-orphan drugs. HTA agencies have a higher acceptance threshold for external comparators (ECs) than for RWD in the case of non-orphan drugs, underscoring the importance of reliable EC data. RWD-based ECs have shown more positive results for non-orphan drug submissions compared to other types of EC data. Furthermore, a significant proportion of single-arm trial-based submissions focus on oncology and hemato-oncology indications, reflecting the industry's emphasis on novel targeted therapies, cellular and gene therapies, and rare cancers with biomarker-targeted subpopulations. This presents an opportunity to leverage RWD for studying these specific populations and therapeutic interventions.<br>Between 2015 and 2019, there was an observed increase in the use and acceptance of external comparators (ECs) based on RWD, while the acceptance of submissions without ECs decreased. Notably, NICE demonstrated a significantly higher acceptance of non-randomized clinical trial data compared to other HTA agencies. |
| Tolley, K. (2010) | The use of observational studies, including registries, is anticipated to gain greater importance in the post-marketing phase, as initiatives are increasingly being implemented to evaluate the effectiveness and cost-effectiveness of medicines in real-life settings.<br>Certain countries, like the Netherlands, have implemented conditional drug reimbursement schemes, where reimbursement is provided temporarily for expensive and innovative medicines, with subsequent provision of evidence-based RWE. A similar case was observed in the UK regarding multiple sclerosis. As a result, drug access strategies are being formulated, which will eventually lead to increased demand for data beyond traditional clinical trials. In light of these developments, companies should consider adapting their research and development strategies accordingly.<br>Traditional randomized clinical trials alone may not be adequate for the comprehensive evaluation of health technologies, unlike in the regulatory process where they currently remain the primary approach. To address the requirements of drug access and reimbursement, a broad range of studies is necessary, considering both internal and external validity. Pragmatic trials and observational studies tend to offer a higher level of external validity, which is crucial when assessing cost-effectiveness relationships. |
|  | When data from RCTs are insufficient for making informed decisions, it is crucial to implement rigorous and |
| Kent, S. et al, (2021) | comprehensive procedures to ensure the quality of data derived from non-randomized studies. Recommendations for addressing the issues encountered in conducting studies:<br>A) Provide justification for the need to conduct the study and highlight the use of non-randomized study data.<br>B) Design prospective studies and engage in early scientific advisory processes.<br>C) Understand potential systematic errors and provide clear references to mitigation and management strategies.<br>D) Perform sensitivity analyses for hypothetical scenarios.<br>E) Pre-specify protocols with clear references to statistical methods.<br>Z) Provide comprehensive reporting of methods and results, with the additional use of checklists.<br>H) Offer quantitative evidence when identifying uncertainty.<br>Θ) Describe impartiality and utilize well-validated tools for assessing overall risk.<br>Recommendations for addressing issues in HTA:<br>A) Enhance and standardize the provision of scientific advisory processes to determine the value of specific evidence. B)<br>Strengthen conditional reimbursement processes to ensure the production of further evidence after initial reimbursement decisions.<br>C) Invest in and develop skills of HTA agency personnel in the design, analysis, and interpretation of non-randomized studies.<br>D) Issue guidelines on best practices.<br>E) Support further research on non-randomized studies and international initiatives for high-quality data. |
| Jaksa A, et al, (2022) | While the organizations did not reach a consensus on the quality and impact of external control arms (ECAs), two main criticisms were identified: selection bias and confusion. These concerns can be addressed through careful data selection and study design, aiming to minimize bias and improve clarity. The incorporation of RWD and RWE in HTA offers an opportunity to complement single-arm oncology trials with contextual information on patient experience. This broader inclusion of data can provide a more comprehensive understanding of treatment outcomes and patient perspectives. Exploring the evaluation of ECAs beyond oncology and in different conditions is important to gain insights into their influence on regulatory and HTA decision-making processes. This ongoing exploration can contribute to enhancing the understanding and utilization of ECAs in a wider range of therapeutic areas.<br>Selection bias and confounding emerged as common criticisms of ECAs, emphasizing the importance of utilizing high-quality RWD and implementing rigorous study designs. There is a clear need for additional guidance on study design and implementation when utilizing ECAs, as well as specific guidance on RWD-based ECAs from regulatory bodies such as the FDA. |
| Justo N, et al, (2019) | The HTA organizations must strike a balance between promoting local RWE without excessively delaying assessments. Assessments can be expedited by considering international RWE after addressing transferability concerns. The expansion of healthcare coverage and the inclusion of a growing number of diseases and interventions are exerting pressure on healthcare budgets. RWE can be invaluable for monitoring the outcomes and cost-effectiveness of high-cost interventions, making careful monitoring essential. Enhancing data recording: Initiatives aimed at improving data recording, such as the implementation of electronic medical records (EMRs), standardizing coding systems, reducing free |
|  | text, and providing coding systems training to staff, are viewed as opportunities to enhance the usability of RWD.<br>Establishment of RWD units and evidence-based policy: The increasing number of HTA units and the adoption of pharmacoeconomic guidelines and evidence-based policy planning are encouraging trends for the utilization of RWE. These advancements offer a promising outlook for the expanded use of RWE in South America. |
| Kamusheva M, et al, (2022) | Adequate management of collected RWE from other countries is crucial, and the electronic collection of RWE is necessary. However, some Central and Eastern European countries face challenges due to their limited operational information technology infrastructure. Furthermore, the lack of government stability and frequent policy changes in the healthcare sector result in regularly amended and unstable legislation. The dynamic environment leads to constant changes in local legislative requirements for HTA and significant differences in approaches to managing the process of RWE utilization. Therefore, achieving stability is essential in this regard.<br>Moreover, collaborative initiatives and infrastructure development are expected to increase to facilitate the transferability of RWE to CEE countries. The focus will shift towards determining the appropriate implementation of RWE. Lastly, there is a need for improvement in factors such as pharmacotherapy guidelines and the availability of health technologies in certain CEE countries. Additionally, the development of suitable statistical analysis methods for the transmission of RWE is urgently required. |
| Timbie JW, et al, (2021) | Recommendations for addressing the issues related to the inclusion of RWE:<br>1) Provide more specific and up-to-date guidance on the utilization of RWE, including successful examples of its application. Industry stakeholders emphasize the need for clear guidance from payers, which would assist in designing studies using RWD.<br>2) Enhance the robustness of RWE by improving study design and analytical methods. This would ensure that the research conducted using RWD meets rigorous standards and produces reliable and valid results.<br>3) Engage in RWE-related activities sponsored by organizations like NESTcc, CDRH, and industries, which bring together stakeholders to develop a framework for enhancing RWE generation and detection methods.<br>4) Take part in the FDA's Payor Communication Task Force and share insights gained from working group discussions. This collaboration allows the industry to gather important information for designing studies using RWD, while payers gain a deeper understanding of reimbursement decisions in studies utilizing RWD.<br>5) Develop alternative reimbursement strategies supported by RWE when sufficient experimental data is not available to support full coverage. Some payers are exploring alternative payment arrangements that are tied to the performance of medical devices in real-world settings, providing an avenue for reimbursement based on RWE outcomes. |
| Bullement A, et al, (2020) | To enhance the utilization of RWE in submissions to NICE (National Institute for Health and Care Excellence), it is advisable for companies to take proactive steps in addressing common criticisms associated with RWE. This includes justifying the methodologies employed for RWE analysis and demonstrating the relevance and reliability of the RWE sources used. Additionally, the development of best practice guidelines for reporting RWD and adhering to these standards when presenting RWE can significantly enhance the effectiveness of documentation submitted to NICE.<br>RWE has demonstrated its significance in decision-making processes by filling data gaps in cost-effectiveness analyses |
|  | submitted by companies. This data is ultimately utilized to inform decisions, particularly through its impact on the ICER (incremental cost-effectiveness ratio) estimate. This study indicates that the utilization of RWE in NICE submissions for cancer drugs is widespread and generally proves to be a valuable source of information, effectively supporting the decision-making process. |
| Al-Omar HA, et al, (2021) | In relation to the challenges associated with RWD in the assessment, treatment, and evaluation (ATY) process, the following observations were made:<br>Availability and accuracy of data: The availability and accuracy of data, including registries, RWE, quality of life (QoL), and costing data, were deemed important by 86.4% of participants.<br>Scattered healthcare system in Saudi Arabia: The fragmented nature of the healthcare system in Saudi Arabia was recognized as a challenge, which could impact the collection, integration, and accessibility of reliable RWD.<br>Lack of acceptance by stakeholders: There was a lack of acceptance and recognition of the value of RWD by various stakeholders involved in the HTA process, which may hinder its effective utilization.<br>Lack of expertise and human resources: Insufficient expertise and limited human resources in the field of RWD were identified as challenges, potentially impeding the effective analysis and interpretation of available data.<br>Lack of reliable data on diseases: The absence of reliable and comprehensive data on specific diseases was highlighted as a challenge, which may limit the applicability and generalizability of RWD in the HTA process. |
| Makady A, et al, (2018) | Alignment with policies: The findings of this study are in line with a previous review of policies regarding the use of RWD by HTA organizations. The review discovered that the current utilization of RWD in practice is consistent with the policies, which may vary slightly between agencies and depending on the specific context being analyzed.<br>Pragmatic type trials (PCTs) as a potential source of RWD: PCTs were suggested as a potential means of gathering RWD that can bridge the gap between randomized controlled trials (RCTs) and real-world settings.<br>Quantitative methods and sensitivity analyses: The study discusses quantitative modeling and sensitivity analysis methods, such as bootstrapping and probabilistic sensitivity analyses (PSA), as potential alternatives to RWD for addressing the efficacy-effectiveness gap. However, it should be noted that these methods are still based on hypotheses and RCT data, and the importance of RWD remains crucial for evaluating long-term efficacy in a diverse clinical population.<br>Potential role in re-assessments: RWDs could play a significant role in the reassessment of drugs by confirming previous efficacy estimates, cost-effectiveness ratios (ICERs), and budget implications. Comprehensive re-evaluation reports conducted under conditional reimbursement systems (CRS) could greatly benefit from the utilization of RWDs. |
| Deverka PA, et al, (2020) | Opportunities for training and development incentives in RWE and NGS: A. Offer training programs on observational study methods and pragmatic type trials (PCTs) to stakeholders, including payers, to enhance their understanding and utilization of RWE.<br>B. Encourage funding initiatives to support the conduct and publication of RWE studies that demonstrate the benefits and risks associated with NGS testing. This would provide valuable evidence to inform coverage decisions.<br>Opportunities for improving methods of generating RWE:<br>A. Existing RWE assessment tools developed specifically for payers should be adapted to the context of NGS through the collaboration of multi-stakeholder groups. This would ensure best practices and evaluation tools are tailored to the unique aspects of NGS.<br>B. Develop transparent engagement processes that involve payers to better understand their information needs and incorporate them into the design and execution of RWE studies.<br>C. Additionally, the use of artificial intelligence (AI)-based methods, such as natural language processing, machine learning, and deep learning, can be leveraged to process and analyze unstructured data from electronic health records (EHRs) and patient-generated data. This would alleviate the burden of manual curation and enhance the efficiency of data analysis in RWE studies.<br>Despite the potential benefits, payers still heavily rely on clinical guidelines and randomized controlled trials (RCTs) as the primary sources of clinical evidence for coverage decisions, rather than RWE. Several factors contribute to this reliance, including challenges surrounding the quality of RWE data, standardization of genomic data representation, limited involvement of payers in study development, and a preference for RCTs over observational data. Efforts are underway to address these barriers, such as the development of data infrastructure, increased payer participation in study design, adherence to methodological best practices, and consensus on NGS guidelines.<br>Outcomes-based contracts (OBCs) between industry and payers can benefit from the utilization of RWE. However, challenges related to obtaining accurate data, defining appropriate outcome measures, ensuring patient data privacy, and managing costs often impede the widespread implementation of OBCs. |
| Lou J, et al, (2020) | The development of a guidance document specifically tailored to the context of Asia can be instrumental in addressing the challenges identified in this study. This guidance document can provide recommendations, best practices, and standardized approaches for the utilization of RWD and RWE in the region, thereby supporting stakeholders in navigating the complexities and mitigating the identified challenges.<br>Collaboration among various stakeholders, including healthcare providers, researchers, policymakers, and industry representatives, is crucial in overcoming the challenges associated with RWD/RWE utilization. By fostering collaboration and promoting the adoption of good practices, stakeholders can share knowledge, experiences, and expertise, leading to improved methodologies, data quality, and overall effectiveness in utilizing RWD/RWE.<br>All participants reached a consensus that RWE should serve as supplementary documentation and is unlikely to replace data derived from clinical trials for reimbursement decisions. The findings suggest a recognition of the potential of RWD/RWE in HTA, along with a need for further understanding, capacity-building, and improved collaboration among |
|  | stakeholders to fully harness the benefits of utilizing RWD/RWE in the HTA process. |
| Facey KM, et al, (2020) | Opportunities and applications:<br>A. Collaborating with the academic community to develop methods for utilizing RWD in the decision-making process, as well as promoting cooperation between countries, especially in the context of rare diseases.<br>B. Involving the industry in multilateral dialogues regarding RWD, through initiatives such as public-private partnerships and international programs like the IMI Big Data for Better Outcomes in Europe and the US FDA's Sentinel program.<br>C. Governance and transparency: Ensuring transparency in the production of RWE is crucial. Efforts are being made to address this issue through initiatives like the ISPOR RWE Transparency Task Force, which aims to promote transparency in the generation and reporting of RWE.<br>D. EU Multilateral Learning Network for RWE: Establishing a learning network that involves relevant stakeholders is essential for harnessing the potential of RWD and RWE in healthcare. This network should focus on data standardization, improving data quality, facilitating data sharing, and implementing robust data processing and analysis methods.<br>E. Regulatory support: Regulatory agencies, such as the US FDA, are actively engaged in RWE projects aimed at providing guidance on the relevance and reliability of RWD. These initiatives involve leadership engagement and target shared knowledge and consistency in regulatory approaches. They are widely recognized as necessary measures to address the challenges associated with RWD/RWE and ensure their effective utilization in regulatory decision-making processes. |
| Bowrin K, et al, (2019) | According to a study included in the review, the limitations encountered in modeling data through registries can be addressed by using a practical guide for utilizing registry data to inform evidence-based decisions regarding the cost-effectiveness relationship of new drugs. Furthermore, the evaluation of cost-effectiveness models using observational data can be achieved using a specialized checklist that includes five questions for assessing statistical issues.<br>Opportunities:<br>According to ten (10) guidelines from organizations, it has been determined that RWE is useful (but not mandatory) in providing evidence for clinical practice regarding treatment pathways, comparative interventions, resource utilization and costs, long-term natural history of the disease, as well as the actual effectiveness and safety of interventions. Sixteen (16) pharmacoeconomic guidelines recommend or consider the integration of RWE into pharmacoeconomic models and HTA submissions, with Canada, Belgium, and Poland recommending the incorporation of RWD into HTA submissions. |
| Brogaard N, et al, (2021) | Monitoring future assessments and potential reassessments would provide a better acceptance of RWD as a data source for tumor-agnostic therapies, and further guidance is needed to clarify what is acceptable regarding indirect comparisons.<br>Conditional reimbursement: Some countries, such as England and France, have established conditional approval processes. This allows treatments with data uncertainty to receive funding recommendations while requiring the provision of additional data at a later date. This provides an opportunity for the acceptance of currently available evidence, including RWD/RWE, with the potential for further data collection and evaluation. |
|  | Lack of guidance for HTA evaluations: There is a need for more guidance on how HTA evaluations and economic models should be structured for future oncology-agnostic therapies. Some HTA systems are developing internal guidelines to ensure consistent assessment of oncology-agnostic therapies, but further guidance is still needed to address uncertainties in clinical data and the integration of RWD. |
| Hogervorst Milou A, et al, (2022) | RWD could be considered acceptable in the following situations:<br>A. In cases of high disease burden or when the indications of the evaluated treatment are severe or even life-threatening.<br>B. If the findings of randomized controlled trials (RCTs) are outdated or conflicting with the available literature of RCTs.<br>C. When there is uncertainty regarding resource utilization in clinical practice.<br>D. In the case of highly innovative health technologies or when alternative access is not feasible.<br>E. When the trials used for licensing are compared with treatments not used in the country's practice.<br>F. In situations of lack of reliable evidence and when there is uncertainty in the clinical domain as well as a high degree of uncertainty in cost-effectiveness analyses.<br>G. In cases of well-established use, according to European legislation. |
| Sievers H, et al, (2021) | Capabilities of RWE include:<br>A. Filling gaps and reducing uncertainty: RWE can provide supplementary documentation that supports the data obtained from Randomized Controlled Trials (RCTs), helping to address any gaps or uncertainties in the evidence base.<br>B. Demonstrating real-world value: RWE allows for the evaluation of a medicine's effectiveness and value in everyday clinical practice, providing insights into its performance outside the controlled environment of clinical trials.<br>C. Ethical considerations: In situations where conducting an RCT may be ethically challenging or not feasible, RWE offers an opportunity to gather evidence and generate insights.<br>Opportunities associated with RWE include: Harmonization and greater acceptance:<br>A. RWE presents an opportunity for harmonizing documentation requirements and increasing the acceptance of RWE as a valuable source of evidence in healthcare decision-making processes.<br>B. Stakeholder dialogue and consensus: Through pHTAorms like the Joint Scientific Advice, stakeholders can engage in discussions and reach consensus on various issues, bridging the gaps between regulatory requirements and the expectations of HTA agencies.<br>C. Early joint dialogues: Particularly during the stages when the European Medicines Agency (EMA) is considering |
|  | technology licensing, early joint dialogues involving multiple stakeholders can help facilitate a more comprehensive understanding of the evidence landscape.<br>D. Alignment of HTA documentation: The EUnetHTA pHTAorm provides an opportunity for aligning documentation requirements between different HTAs, promoting consistency and efficiency in the evaluation of healthcare technologies. |
| Hampson G, et al, (2018) | Opportunities for Improving the Use of RWE:<br>A) Enhancing the quality and reliability of RWE studies through the establishment of a national mandatory registry for observational studies, national data repositories, investments in the quality and consistency of electronic medical records, and the development of strict protocols and consensus on best practice guidelines.<br>B) Implementing effective governance regulations that clarify the scope of RWE data that can be shared.<br>C) Greater commitment to the development of pragmatic trial designs, which can bridge the evidence gap between RWE and Randomized Controlled Trials (RCTs).<br>D) Fast-track approval processes, such as accelerated approval pathways, for various innovations where sufficient evidence is lacking. In these processes, RWE can play a decisive role. |
| George E, (2016) | Opportunities for Improving Non-Randomized Controlled Trials (non-RCTs):<br>A. International Collaboration: The participation of NICE in cross-border projects, such as the IMI Get Real project and the ADAPT SMART project, can contribute to the development of frameworks and methodologies for integrating RWD into HTA processes.<br>B. Areas for Improvement: There are opportunities to enhance the usability and reliability of non-RCT data. This includes improvements in case identification and data source linkage, the routine capture of health-related quality of life (HRQL) and patient preferences, the inclusion of important patient variables (sociodemographic, disease severity, comorbidities), and explicit variable definitions to minimize data gaps.<br>C. Complementary Use of RCTs and RWD: RCTs alone cannot address all the needs of HTA and decision-makers. The utilization of data from non-RCT sources provides an opportunity to strengthen evidence standards and generate evidence throughout the technology lifecycle. RCTs and RWD can complement each other to provide a more comprehensive understanding of effectiveness and safety in healthcare interventions. |
| Makady E, et al, (2017) | The lack of alignment and guidance regarding the practical aspects of RWD collection and analysis may discourage marketing authorization holders from investing in RWD generation for HTA purposes. Clarity and harmonization of policies could provide incentives for stakeholders to collect and utilize RWD more effectively.<br>The approach of policy alignment regarding the use of RWD by HTA organizations in Europe is useful, and the presentation of guidelines for the collection and analysis of such data is recommended. Policy harmonization can appropriately support the industry in producing additional or alternative data for drugs where RCTs cannot provide strong evidence. EUnetHTA can support harmonization through a discussion pHTAorm. The use of RWD can be particularly valuable for rare diseases or orphan drugs where conducting randomized controlled trials (RCTs) may be challenging. |
|  | RWD can provide data on treatment outcomes in situations where conventional clinical trials are not feasible. |
| Husereau, D, et al, (2019) | Opportunities:<br>The use of RWD is expected to increase in the future, supported by technological advancements, the growing availability of data, and the increasing demand for evidence to support decision-making. To fully harness the potential of RWD, there is a need for continuous investment in data infrastructure, including standardization and harmonization of data, development of analytical tools, and personnel training.<br>There is a need for greater collaboration and partnerships among stakeholders, including researchers, regulatory authorities, payers, and industry, to ensure the quality and reliability of RWD and promote their use in decision-making. These opportunities highlight the potential for leveraging RWD to generate RWE that can inform healthcare policies, improve patient outcomes, and enhance the efficiency and effectiveness of healthcare interventions. By addressing the challenges and investing in the necessary infrastructure and collaborations, RWD can be a valuable resource in shaping the future of healthcare. |
| Pongiglione B, et al, (2021) | Ensuring standardization is essential, encompassing consistent and appropriate selection, measurement, utilization, and reporting of results in clinical research and practice. This approach will aid in mitigating biases and enhancing the validity and reliability of findings. Enhanced coordination at the European Union (EU) level would be particularly advantageous in facilitating the generation of comparable RWD across countries. To further advance the utilization of RWD, a collaborative approach involving stakeholders is necessary for the initiation, design, and analysis of RWD, with industry cooperation. If discussions at the EU level regarding early HTA and the requirement for registers or observation studies become more prevalent, the quality and nature of RWD collected will assume increasing importance in informing policy decisions. |
| Ciminata, G, (2019) | There is a need to standardize the collection, management, and analysis of RWD in order to enhance the quality and comparability of the results. By incorporating RWD, cost-effectiveness analyses can inform HTA decisions regarding the utilization of DOACs versus warfarin in individuals with atrial fibrillation. |

HTA: Health Technology Assessment, RWD: Real-World Data, RWE: Real-World Evidence, NICE: National Institute for Health and Care Excellence, ECs: External Comparators, FDA: U.S. Food and Drug Administration, EMRs: Electronic Medical Records, CEE: Central and Eastern European, NESTcc: National Evaluation System for health Technology Coordinating Center, CDRH: Center for Devices and Radiological Health, ICER: Incremental Cost-Effectiveness Ratio, EMA: European Medicines Agency, IMI: Innovative Medicines Initiative, RCTs: Randomized Controlled Trials, HRQL: Health-Related Quality of Life, EU: European Union, DOACs: Direct Oral Anticoagulants

The first study conducted by Hagen et al. (2021) [17] focused on the Norwegian HTA system and highlighted the importance of patient registries as valuable sources of evidence. The study emphasized that well-organized registries have the potential to provide crucial data on health utilities, resource use, and costs, enhancing the assessment of health technologies and informing decision-making processes. Patient registries were found to offer insights into treatment patterns, real-world treatment adherence, and the natural history of diseases, contributing to accurate economic evaluations and modeling. In the editorial by Gonçalves (2020) [41], the integration of ethical evaluation into the HTA process for advanced therapy medicinal products (ATMPs) is discussed, with a specific focus on the use of RWD. The editorial emphasizes the importance of collecting and analyzing evidence from RWD to complement the assessment of ATMPs, particularly in the context of rare diseases. It highlights the need for long-term efficacy and comparative effectiveness data, supplemented by real-world evidence, to ensure comprehensive evaluations of these advanced therapies. Fasseeh et al. (2020) [18] conducted a Middle East and North Africa Primary Survey to assess the implementation of HTA in the region. The study revealed a willingness among respondents to invest in patient registries and make payer databases accessible to the HTA sector. It underscored the importance of collecting and utilizing local data to enhance the quality of HTA practices. Furthermore, Leahy et al. (2020) [19] conducted a UK Systematic Review that explored the utilization of evidence from primary care databases in NICE HTA assessments. The study identified primary care databases as a key source of RWD for informing NICE submissions and aiding healthcare decision-making. The acceptance and positive feedback from review committees and the Evidence Review Group (ERG) indicate the reliability and accuracy of primary care databases as a data source. Another systematic review by Fuchs et al. (2016) [20] with the aim to review and compare the activities of HTA for medical devices among European organizations involved in Mutual Recognition Agreements (MRAs), highlighted various initiatives to address the challenges in medical device assessment, such as standardized data elements, outcome measures, and data exchange platforms. These initiatives aim to enhance the consistency and quality of MD assessment processes across European HTA organizations. Dai et al. (2021) [42] evaluated the proposed re-evaluation process for cancer medicines in Canada, focusing on the use of RWD and indicated that stakeholders expressed confidence in the results, citing the availability of RWE on clinical benefit and cost-effectiveness from various Canadian provinces. The study recommended involving patients in the reassessment process and leveraging RWE for studying rare cancers and targeted therapies. Patel et al. (2021) [21] conducted an international retrospective study reviewing HTA submissions from various countries. The study highlighted the need for a well-designed strategy and robust data collection methods for indirect treatment comparison. It emphasized the increasing use of RWD in HTA submissions and the importance of monitoring and developing guidelines for RWD submissions. Tolley (2010) [22] offered a commentary on the evaluation of new pharmaceutical technologies within the UK’s HTA system. The commentary highlighted the importance of observational studies, including registers, for assessing the real-life efficacy and cost-effectiveness of medicines during the post-marketing phase. It emphasized the need for conditional drug reimbursement schemes and the adaptation of research and development strategies to meet the demand for evidence from unproven clinical trials. On the other hand, the author discusses challenges in evaluating new pharmaceutical technologies within the UK’s HTA system and highlights the importance of real-world data during the post-marketing phase. Kent et al. (2021) [23] provides recommendations for using evidence from non-randomized trials in HTA processes, emphasizing study design, reporting potential errors, and strengthening scientific advice processes. Jaksa et al. (2022) [24] review critiques of ECAs utilizing real-world data, highlighting selection bias and confusion as key issues. The expansion of healthcare coverage and increasing number of diseases and interventions covered put pressure on healthcare budgets as mentioned in the systematic review and qualitative research through workshops with stakeholders conducted by Justo et al. (2019) [25]. RWE can help monitor outcomes and cost-effectiveness, but careful data recording and evidence-based policy planning are crucial. Kamusheva et al. (2022) [43] conducted a scoping review and qualitative research by having internal discussions and a webinar with stakeholders from Central and Eastern European countries to identify barriers to the use of RWE in healthcare. The study focused on Central and Eastern European countries. Those countries face challenges in collecting RWE due to limited IT infrastructure and frequently changing legislation. Collaborative initiatives and infrastructure development are needed. Timbie et al. (2021) [26] explored the use of RWE for medical devices, recommending clearer guidance, rigorous study design, engagement in RWE-related activities, and alternative reimbursement strategies. Bullement et al. (2020) [27] conducted a systematic review with the aim of how RWE has been used to inform Single Technology Assessments (STAs) of cancer drugs conducted by the NICE. Developing best practice guidelines and addressing common criticisms can enhance the use of RWE in submissions to NICE. Al-Omar et al. (2021) [28] identify challenges in utilizing RWD in HTA processes in Saudi Arabia, including data availability, acceptance by stakeholders, and expertise and resources. In Makady’s, et al. (2018) [29] literature review focusing on England, Scotland, Netherlands, France, and Germany, it was examined whether RWDs are included in the REAs and CEAs of melanoma drugs, and the evaluation of RWDs for their intended purposes by five HTA organisations in Europe. Alignment with policies, quantitative methods, and the role of RWD in re-evaluations are discussed. RWDs could play a critical role in drug reassessment by confirming previous efficacy estimates, cost-effectiveness ratios (ICERs) and budget implications. Comprehensive re-evaluation reports under conditional reimbursement systems (CRS) could benefit from the use of RWDs. Deverka et al. (2020) [30] examined the use of RWE in coverage decision-making for NGS-based tests. Opportunities for training and development incentives in RWE and NGS were identified, including providing training on observational study methods and pragmatic trials for stakeholders, particularly payers. Additionally, encouraging funding for RWE studies that demonstrate the benefits and harms of NGS testing was recommended. Improving the methods of studies that generate RWE was also highlighted. Several groups have developed RWE assessment tools specifically for payers, and it was suggested that multi-stakeholder groups should adapt these best practices and evaluation tools to NGS. Furthermore, transparent engagement processes should be established to incorporate payer information needs. Lastly, the study emphasized the potential of artificial intelligence (AI)-based methods, such as natural language processing and machine learning, in processing and analyzing unstructured data from electronic health records and patient-generated data, reducing the need for manual curation.

The first study conducted by Hagen et al. (2021) [17] focused on the Norwegian HTA system and Lou et al. (2020) [31] conducted a study across multiple Asian countries, examining the use of real-world data and evidence in the assessment of health technologies in the general population. The study identified challenges and proposed the development of a guidance document specifically tailored for Asia to address these challenges. Collaboration and the adoption of good practices among stakeholders were emphasized as crucial strategies to overcome these challenges and effectively utilize real-world data and evidence in health technology assessment processes across Asia. Facey et al. (2020) [44] investigated the use of RWD in decision-making by payers and HTA organizations in European Union countries. The study employed a mixed-methods approach and identified several opportunities and applications for leveraging RWD, including collaboration with academia, engaging industry in multilateral dialogues, promoting governance and transparency in RWD production, establishing an EU multilateral learning network for RWD, and seeking regulatory support and guidance. These actions have the potential to improve the use of RWD in decision-making processes, enhance the reliability and relevance of RWD, and foster knowledge exchange in the health sector across European Union countries. Bowrin et al. (2019) [32] conducted a systematic review to investigate the limitations of using RWE in decision analysis, particularly in modeling, and to identify existing recommendations on RWD-based modeling. The review highlighted the need for a practical guide on utilizing registry data for cost-effectiveness decisions and proposed a checklist to assess statistical issues when evaluating cost-effectiveness models with observational data. It also revealed opportunities for incorporating RWE in decision-making, as several agency guidelines recognized its usefulness in providing evidence on clinical practice, treatment pathways, resource utilization, disease natural history, and intervention efficacy and safety. Brogaard et al. (2021) [33] analyzed HTA agency assessments and reimbursement decisions for tumor-agnostic therapies in multiple countries. The study focused on the role of RWD in these assessments and highlighted the importance of monitoring future evaluations and potential re-evaluations to enhance the acceptability of RWD as a data source. Conditional reimbursement procedures and the need for clear guidance on indirect comparisons and acceptable use of RWD were identified as opportunities. The study also emphasized the lack of guidance on structuring HTA assessments and developing economic models for future tumor-agnostic therapies, calling for further guidance and integration of RWD into the assessment process. Hogervorst et al. (2022) [34] conducted qualitative research in European countries to assess the challenges associated with the acceptance of RWD in the evaluation of new health technologies. The study identified circumstances where RWD could be deemed acceptable, such as high disease burden, severe indications, outdated or inconsistent RCT findings, uncertainty regarding resource utilization, innovative technologies, limited treatment options, and clinical practice comparisons. The study also highlighted the importance of reliable documentation, clinical uncertainty, high uncertainty in cost-effectiveness analyses, and well-established use according to European legislation as factors that could warrant the use of RWD. Sievers et al. (2021) [35] conducted qualitative research in several European countries to explore stakeholder perceptions of post-marketing RWE and its value in healthcare decision-making. The study identified key challenges and opportunities related to RWE utilization, emphasizing its potential to complement RCT data, address gaps, and reduce uncertainty. Opportunities included harmonizing documentation requirements, fostering greater acceptance, and facilitating dialogue among stakeholders through platforms like the Joint Scientific Advice and EUnetHTA. The study highlighted the importance of coordinating post-marketing RWE requirements between regulatory agencies and HTA organizations. Hampson et al. (2018) [36] explored the use of RWE in the healthcare system in the USA. They identified opportunities for improvement, including the establishment of a national mandatory register for observational studies, national data repositories, and better quality electronic medical records. The study recommended rigorous protocols and consensus on good practice guidelines for generating high-quality RWE. Effective governance arrangements for data sharing and pragmatic trials that bridge the gap between RWE and TC were also suggested. The study proposed fast-track licensing processes to utilize RWE for decision-making on innovative therapies with limited documentation. George (2016) [37] focused on using RWD in health technology assessments by the NICE in the UK. The article emphasized the need for improvements in non-RCT data, including better case identification, data source linkage, and standardized recording of health-related quality of life and patient preferences. Transnational participation in projects such as IMI Get Real and ADAPT SMART was highlighted to develop frameworks and methodologies for integrating RWD into NICE assessments. The article suggested that using both RCTs and RWD can provide a more comprehensive understanding of healthcare interventions’ effectiveness and safety. Makady et al. (2017) [38] reviewed policies of six health technology assessment organizations in Europe regarding the use of RWD in assessments of medicines. The study found a lack of alignment and guidance on practical aspects of RWD collection and analysis, which may discourage investment in RWD production. The researchers recommended aligning policies and providing guidance on RWD collection and analysis to incentivize stakeholders. Harmonization of policies can support the generation of additional or alternative data for medicines where RCTs are not feasible, especially for rare diseases or orphan drugs. Husereau et al. (2019) [45] conducted qualitative research in Canada to explore stakeholders’ views on using RWD for drug pricing and reimbursement decisions. The study highlighted the need for investments in data infrastructure, standardization, analytical tools, and staff training to fully leverage the potential of RWD. Collaboration among researchers, regulators, payers, and industry stakeholders was emphasized to ensure the quality and reliability of RWD. Data standardization and harmonization, along with cooperation and partnerships, were identified as important for effective utilization of RWD in decision-making processes. Pongiglione et al. (2021) [39] conducted a systematic review on the use of RWD in HTA of medical devices in Europe. The study emphasized the need for standardization in the selection, measurement, use, and reporting of RWD to address biases and ensure consistency in clinical research and practice. Stronger coordination at the EU level and a coordinated approach involving stakeholders and industry were recommended to enhance the use of RWD in HTA processes and inform policy decisions. Ciminata (2019) [40] explored the opportunities and challenges of using RWE to support the Authorization for Temporary Use (ATF) decision-making process in Scotland, specifically focusing on anticoagulant medication in patients with atrial fibrillation. The study emphasized the need for standardization in RWD collection, management, and analysis to improve the quality and comparability of results.

## 4. Discussion

In this study, we conducted a systematic review to comprehensively evaluate the available evidence on RWD and RWE for HTA process. Multiple included studies emphasize the significance of RWD and RWE as alternative sources of evidence when RCTs are lacking or not feasible. While HTA organizations typically favor systematic reviews and RCTs as primary sources of clinical effectiveness data for economic evaluations of health technologies, they also recognize the value of observational studies, patient registries, and primary care databases as valuable sources of real-world evidence. However, the acceptance and utilization of RWD/RWE vary among different HTA organizations and countries. Over time, there has been a noticeable trend towards increased acceptance and use of RWD/RWE in HTA. Nonetheless, certain challenges and limitations associated with its acceptance and use in HTA have been highlighted in studies, including concerns about bias, methodological issues, and the need for further discussions and considerations. Although stakeholders, including HTA agencies, payers, and pharmaceutical companies, generally acknowledge the value of RWD/RWE, there is still a prevailing preference for data from RCTs. RWD/RWE is frequently employed in HTA for several purposes, including describing disease epidemiology, illustrating treatment landscapes, providing clinical inputs for cost-effectiveness models, assessing long-term effectiveness and costs, and evaluating health-related quality of life. However, there is regional variation in the implementation of HTA and the use of RWD/RWE, with differences observed between countries in Europe, South America, the Middle East, and Asia.

On the other hand, this review concluded important findings in regards to the barriers and issues arize with the use and acceptance of RWD and RWE for HTA. These findings highlight the challenges faced in leveraging real-world data for informed decision-making in healthcare. One of the prominent barriers identified is the limited availability and transferability of local real-world data. This limitation poses a challenge in accessing comprehensive and relevant data sources, particularly in specific regions or healthcare contexts. The lack of local data hinders the ability to generate evidence that is tailored to the specific needs and characteristics of the population under assessment. Accessing high-quality data is crucial for reliable and credible evidence generation. However, several studies revealed difficulties in accessing reliable and high-quality real-world data. The challenges can arise from issues such as data privacy and confidentiality concerns, limited data standardization, and variations in data collection and reporting practices. These barriers undermine the reliability and credibility of the evidence derived from real-world data sources. Methodological challenges were also identified as a significant barrier to utilizing real-world data and evidence in HTA. Studies pointed out challenges in study design, analysis, and reporting when relying on non-randomized clinical evidence. Addressing these methodological challenges is crucial to ensure the validity and robustness of findings derived from real-world data sources. Insufficient expertise among stakeholders in utilizing real-world evidence emerged as a common barrier. This lack of expertise can hinder the effective use and interpretation of real-world data. Stakeholders, including policymakers, payers, and clinicians, need to possess the necessary skills and knowledge to critically evaluate and utilize real-world evidence in decision-making processes. Fragmentation and lack of collaboration among stakeholders were found to hinder the utilization of real-world data. The absence of harmonized approaches, data sources, methodologies, and decision-making processes limit the consistent and efficient use of real-world evidence. Enhancing collaboration and promoting standardization among stakeholders are essential for maximizing the potential of real-world data in HTA. Data quality and reliability were highlighted as significant concerns. Studies identified issues related to low data quality, confounding biases, incomplete data, and challenges in data protection and confidentiality. These limitations can undermine the validity and generalizability of findings derived from real-world data sources. Overall, the studies underscore the need to address these barriers and challenges to effectively utilize real-world data and evidence in HTA. Improving data availability, ensuring data quality and standardization, addressing methodological challenges, promoting collaboration, and enhancing expertise among stakeholders are key considerations for advancing the use and acceptance of real-world data in healthcare decision-making processes. The results of this review concerning to opportunities related to the RWD inclusion in HTA are in line with the literature and particularly with published manuscript of Crane G, et al. (2022) [46] whom results were similar and highlighted the importance of recommending approaches and initiatives for improving RWE utilization in healthcare decision-making in East Asia and beyond and Encouraging large-scale collaborations among government agencies, hospitals, research organizations, patient groups, and the pharmaceutical industry to ensure access to robust real-world data and alignment on addressing evidence needs.

The systematic review methodology employed in this study offers several strengths, enhancing the reliability and credibility of our findings. One of the key strengths of a systematic review is its comprehensive and rigorous approach. By adhering to a predefined and transparent methodology, we ensured that all relevant studies on the research question were identified, appraised, and synthesized. This minimizes bias and increases the validity of our results. An additional notable strength inherent to the systematic review methodology is its innate capacity to mitigate selection bias. This was achieved through the meticulous application of explicit inclusion and exclusion criteria, thereby effectively diminishing the prospect of selectively favoring studies that align with a particular perspective. Such an approach significantly bolsters the objectivity and neutrality of our review. Moreover, our systematic review facilitated the amalgamation of a diverse body of evidence. We thoughtfully incorporated studies employing a spectrum of methodologies, spanning various populations and settings. This comprehensive and inclusive approach underscores the robustness of our findings, fostering a more holistic perspective on the research question at hand. Despite these strengths, it is important to acknowledge the limitations of our systematic review. Firstly, although we aimed to conduct a comprehensive search, it is possible that some relevant studies may have been inadvertently missed, mainly due to the strict inclusion criteria of English-written manuscripts. This linguistic restriction may introduce a potential source of bias, as relevant studies or data published in languages other than English were not incorporated into our analysis, which could impact the comprehensiveness and generalizability of our findings. While conscientiously implementing strategies to mitigate bias within this systematic review, it is imperative to acknowledge that the specter of publication bias persists as a potential limitation. Despite our diligence in data collection and analysis, it is challenging to wholly obviate this concern, as it hinges upon the selective dissemination of research findings, rendering an absolute negation of its influence unattainable. In conclusion, it is essential to underscore that the efficacy of systematic reviews is intrinsically tied to the caliber and lucidity of the underlying studies. Regrettably, any deficiencies or methodological shortcomings present within the primary studies inherently permeate our review, thereby potentially undermining the veracity of our collective findings. It is imperative to note that, by and large, the studies incorporated into our review demonstrated commendable quality, although a minority exhibited a moderate level of quality, signaling the importance of interpreting our findings within this context.

## 5. Conclusions

RWD and RWE have gained significance in healthcare decision-making when RCTs are impractical. While systematic reviews and RCTs are preferred for HTA process, HTA organizations acknowledge the value of observational studies, patient registries, and primary care databases. Acceptance and utilization of RWD/RWE vary globally, with a growing trend towards their increased use. However, challenges persist, including concerns about bias, methodological issues, and the need for further discussions. Stakeholders generally recognize the value of RWD/RWE but still prefer data from RCTs. RWD/RWE are frequently employed in HTA to describe disease epidemiology, illustrate treatment landscapes, inform cost-effectiveness models, assess long-term effectiveness and costs, and evaluate health-related quality of life. This review identified key barriers, such as limited availability and transferability of local data, difficulties accessing reliable data, methodological challenges, insufficient expertise, and fragmentation among stakeholders. Data quality and reliability, including issues of low quality, biases, and confidentiality, are also concerns. Overcoming these barriers is crucial for effective use of real-world data in HTA. Improving data availability, ensuring quality and standardization, addressing methodological challenges, promoting collaboration, and enhancing stakeholder expertise are essential considerations. By addressing these challenges, the full potential of real-world data can be harnessed, leading to more informed and evidence-based healthcare decisions.

## Supporting information

S1 Appendix. Search strategy

PRISMA_2020_checklist

S2 Appendix. Quality assessment of included studies

## Data Availability

All relevant data are within the manuscript and its Supporting Information files.

## 6. Acknowledgments

Not applicable/no acknowledgments

