## Supplementary material for "Real-world data: A systematic literature review on the barriers, challenges, and opportunities associated with their inclusion in the health technology assessment process": S2 Appendix. Quality assessment of included studies: CASP-Qualitative-Checklist-2018_Al-Omar HA, et al, (2021).pdf

CASP
Critical Appraisal
Skills Programme

www.casp-uk.net

Summertown Pavilion, Middle
Way Oxford OX2 7LG

### CASP Checklist: 10 questions to help you make sense of a Qualitative research

**How to use this appraisal tool:** Three broad issues need to be considered when appraising a qualitative study:

- Are the results of the study valid? (Section A)
- What are the results? (Section B)
- Will the results help locally? (Section C)

The 10 questions on the following pages are designed to help you think about these issues systematically. The first two questions are screening questions and can be answered quickly. If the answer to both is "yes", it is worth proceeding with the remaining questions. There is some degree of overlap between the questions, you are asked to record a "yes", "no" or "can't tell" to most of the questions. A number of italicised prompts are given after each question. These are designed to remind you why the question is important. Record your reasons for your answers in the spaces provided.

**About:** These checklists were designed to be used as educational pedagogic tools, as part of a workshop setting, therefore we do not suggest a scoring system. The core CASP checklists (randomised controlled trial & systematic review) were based on JAMA 'Users' guides to the medical literature 1994 (adapted from Guyatt GH, Sackett DL, and Cook DJ), and piloted with health care practitioners.

For each new checklist, a group of experts were assembled to develop and pilot the checklist and the workshop format with which it would be used. Over the years overall adjustments have been made to the format, but a recent survey of checklist users reiterated that the basic format continues to be useful and appropriate.

**Referencing:** we recommend using the Harvard style citation, i.e.: *Critical Appraisal Skills Programme (2018). CASP (insert name of checklist i.e. Qualitative) Checklist. [online] Available at: URL. Accessed: Date Accessed.*

©CASP this work is licensed under the Creative Commons Attribution – Non-Commercial-Share A like. To view a copy of this license, visit http://creativecommons.org/licenses/by-nc-sa/3.0/ www.casp-uk.net

CASP
Critical Appraisal
Skills Programme

Paper for appraisal and reference: Value drivers for pharmaceutical products in HTA in Saudi

#### Section A: Are the results valid?

1. Was there a clear statement of the aims of the research?

| | |
|---|---|
| Yes | ✔ |
| Can't Tell | |
| No | |

HINT: Consider
- what was the goal of the research
- why it was thought important
- its relevance

Comments: The primary aim of this research study was to explore and understand the perspectives of a multi-stakeholder group of local Saudi experts regarding the "value drivers" that are important for health technology assessment (HTA) processes and methods, specifically in the context of pharmaceutical products in Saudi Arabia.

2. Is a qualitative methodology appropriate?

| | |
|---|---|
| Yes | ✔ |
| Can't Tell | |
| No | |

HINT: Consider
- If the research seeks to interpret or illuminate the actions and/or subjective experiences of research participants
- Is qualitative research the right methodology for addressing the research goal

Comments: Qualitative methodology seems appropriate for this research as it is well-suited for interpreting and illuminating the subjective experiences and opinions of research participants, which aligns with the research goal of exploring value elements in the context of health technology assessment.

#### Is it worth continuing?

3. Was the research design appropriate to address the aims of the research?

| | |
|---|---|
| Yes | ✔ |
| Can't Tell | |
| No | |

HINT: Consider
- if the researcher has justified the research design (e.g. have they discussed how they decided which method to use)

Comments: The research design, as described in the manuscript text, appears to be appropriate for addressing the research aims, considering the engagement of diverse stakeholders, systematic value driver selection, and a combination of data collection methods. However, a more explicit discussion of the research design's rationale would have provided further insight into its appropriateness.

4. Was the recruitment strategy appropriate to the aims of the research?

| | |
|---|---|
| Yes | |
| Can't Tell | ✔ |
| No | |

HINT: Consider
- If the researcher has explained how the participants were selected
- If they explained why the participants they selected were the most appropriate to provide access to the type of knowledge sought by the study
- If there are any discussions around recruitment (e.g. why some people chose not to take part)

Comments: We can't make a clear assessment of whether the recruitment strategy was appropriate to the aims of the research. While the text mentions that invitations were sent to a diverse set of relevant stakeholders, it lacks specific details regarding the criteria or rationale for selecting these participants.

5. Was the data collected in a way that addressed the research issue?

| | |
|---|---|
| Yes | ✔ |
| Can't Tell | |
| No | |

HINT: Consider
- If the setting for the data collection was justified
- If it is clear how data were collected (e.g. focus group, semi-structured interview etc.)
- If the researcher has justified the methods chosen
- If the researcher has made the methods explicit (e.g. for interview method, is there an indication of how interviews are conducted, or did they use a topic guide)
- If methods were modified during the study. If so, has the researcher explained how and why
- If the form of data is clear (e.g. tape recordings, video material, notes etc.)
- If the researcher has discussed saturation of data

Comments: The data collection methods, setting, and form of data appear to be appropriate for addressing the research aims, as they align with the aim of engaging a multi-stakeholder group to assess value elements in health technology assessment. However, a more explicit discussion of method selection and potential modifications would have strengthened the research methodology.

CASP
Critical Appraisal Skills Programme

6. Has the relationship between researcher and participants been adequately considered?

Yes ☐
Can't Tell ☐
No ☑

HINT: Consider
- If the researcher critically examined their own role, potential bias and influence during (a) formulation of the research questions (b) data collection, including sample recruitment and choice of location
- How the researcher responded to events during the study and whether they considered the implications of any changes in the research design

Comments: The manuscript text does not offer explicit information regarding the researcher's self-reflection on their role, potential biases, and influence during various stages of the research process. A more thorough discussion of these aspects would have provided insight into the researcher's awareness of their own impact on the study and how they navigated potential biases and changes during the research.

#### Section B: What are the results?

7. Have ethical issues been taken into consideration?

Yes ☐
Can't Tell ☐
No ☑

HINT: Consider
- If there are sufficient details of how the research was explained to participants for the reader to assess whether ethical standards were maintained
- If the researcher has discussed issues raised by the study (e.g. issues around informed consent or confidentiality or how they have handled the effects of the study on the participants during and after the study)
- If approval has been sought from the ethics committee

Comments: The study does not provide sufficient details to assess whether ethical standards were maintained in the research. It lacks information about how the research was explained to participants, discussion of ethical issues, and confirmation of ethics committee approval. It is crucial in research involving human participants to address these ethical considerations and provide transparency about the ethical aspects of the study.

8. Was the data analysis sufficiently rigorous?

Yes ✔
Can't Tell ☐
No ☐

HINT: Consider
- If there is an in-depth description of the analysis process
- If thematic analysis is used. If so, is it clear how the categories/themes were derived from the data
- Whether the researcher explains how the data presented were selected from the original sample to demonstrate the analysis process
- If sufficient data are presented to support the findings
- To what extent contradictory data are taken into account
- Whether the researcher critically examined their own role, potential bias and influence during analysis and selection of data for presentation

Comments: While there is room for improvement, the data analysis in the provided texts is reasonably rigorous given the context of the study. The discussion effectively conveys the research findings, explores thematic elements, presents sufficient data, and considers contradictory perspectives. However, explicit details about the analysis process and the researcher's role, potential bias, and influence could enhance the rigor of the analysis.

9. Is there a clear statement of findings?

Yes ✔
Can't Tell ☐
No ☐

HINT: Consider whether
- If the findings are explicit
- If there is adequate discussion of the evidence both for and against the researcher's arguments
- If the researcher has discussed the credibility of their findings (e.g. triangulation, respondent validation, more than one analyst)
- If the findings are discussed in relation to the original research question

Comments: The study provides a reasonably clear statement of findings by explicitly presenting the results of the workshop, discussing evidence for and against arguments, and addressing the research question. However, a more detailed discussion of the credibility of the findings in terms of research methodologies would enhance the overall clarity of the findings.

CASP
Critical Appraisal
Skills Programme

Section C: Will the results help locally?

10. How valuable is the research?

HINT: Consider
- If the researcher discusses the contribution the study makes to existing knowledge or understanding (e.g. do they consider the findings in relation to current practice or policy, or relevant research-based literature
- If they identify new areas where research is necessary
- If the researchers have discussed whether or how the findings can be transferred to other populations or considered other ways the research may be used

Comments: The value of this research can be assessed based on its potential contributions to the field of health technology assessment (HTA) and healthcare decision-making in Saudi Arabia. Overall, the research's value lies in its potential to inform and improve the HTA process in Saudi Arabia by considering local perspectives and challenges. It can be a valuable resource for policymakers, healthcare professionals, and researche
