## Supplementary material for "Real-world data: A systematic literature review on the barriers, challenges, and opportunities associated with their inclusion in the health technology assessment process": S2 Appendix. Quality assessment of included studies: CASP-Qualitative-Checklist-2018_Dai WF, et al, (2021).pdf

Paper for appraisal and reference: Building a National Reassessment Process for Oncology D

### Section A: Are the results valid?

| | | | |
|---|---|---|---|
| 1. Was there a clear statement of the aims of the research? | Yes | ✔ | HINT: Consider<br>• what was the goal of the research<br>• why it was thought important<br>• its relevance |
| | Can't Tell | | |
| | No | | |

Comments:

| | | | |
|---|---|---|---|
| 2. Is a qualitative methodology appropriate? | Yes | ✔ | HINT: Consider<br>• If the research seeks to interpret or illuminate the actions and/or subjective experiences of research participants<br>• Is qualitative research the right methodology for addressing the research goal |
| | Can't Tell | | |
| | No | | |

Comments: Yes, the qualitative research design was appropriate for the study as the authors used a simulated reassessment exercise to collect data.

### Is it worth continuing?

| | | | |
|---|---|---|---|
| 3. Was the research design appropriate to address the aims of the research? | Yes | ✔ | HINT: Consider<br>• if the researcher has justified the research design (e.g. have they discussed how they decided which method to use) |
| | Can't Tell | | |
| | No | | |

Comments: The study used a qualitative research design involving focus group discussions and interviews with stakeholders involved in the reassessment process.

CASP
Critical Appraisal
Skills Programme

4. Was the recruitment strategy appropriate to the aims of the research?

Yes ✔
Can't Tell ☐
No ☐

HINT: Consider
- If the researcher has explained how the participants were selected
- If they explained why the participants they selected were the most appropriate to provide access to the type of knowledge sought by the study
- If there are any discussions around recruitment (e.g. why some people chose not to take part)

Comments: Appropriate as the authors recruited participants who were involved in the reassessment of oncology drugs.

5. Was the data collected in a way that addressed the research issue?

Yes ✔
Can't Tell ☐
No ☐

HINT: Consider
- If the setting for the data collection was justified
- If it is clear how data were collected (e.g. focus group, semi-structured interview etc.)
- If the researcher has justified the methods chosen
- If the researcher has made the methods explicit (e.g. for interview method, is there an indication of how interviews are conducted, or did they use a topic guide)
- If methods were modified during the study. If so, has the researcher explained how and why
- If the form of data is clear (e.g. tape recordings, video material, notes etc.)
- If the researcher has discussed saturation of data

Comments: The authors used a simulated reassessment exercise to collect data on the national reassessment process for oncology drugs.

CASP
Critical Appraisal Skills Programme

6. Has the relationship between researcher and participants been adequately considered?

| | |
|---|---|
| Yes | ✔ |
| Can't Tell | |
| No | |

HINT: Consider
- If the researcher critically examined their own role, potential bias and influence during (a) formulation of the research questions (b) data collection, including sample recruitment and choice of location
- How the researcher responded to events during the study and whether they considered the implications of any changes in the research design

Comments: The authors considered the relationship between researcher and participants by ensuring anonymity and confidentiality of the participants.

### Section B: What are the results?

7. Have ethical issues been taken into consideration?

| | |
|---|---|
| Yes | ✔ |
| Can't Tell | |
| No | |

HINT: Consider
- If there are sufficient details of how the research was explained to participants for the reader to assess whether ethical standards were maintained
- If the researcher has discussed issues raised by the study (e.g. issues around informed consent or confidentiality or how they have handled the effects of the study on the participants during and after the study)
- If approval has been sought from the ethics committee

Comments: By ensuring anonymity and confidentiality of the participants.

Critical Appraisal Skills Programme

8. Was the data analysis sufficiently rigorous?

Yes ✔

Can't Tell ☐

No ☐

HINT: Consider
- If there is an in-depth description of the analysis process
- If thematic analysis is used. If so, is it clear how the categories/themes were derived from the data
- Whether the researcher explains how the data presented were selected from the original sample to demonstrate the analysis process
- If sufficient data are presented to support the findings
- To what extent contradictory data are taken into account
- Whether the researcher critically examined their own role, potential bias and influence during analysis and selection of data for presentation

Comments: The authors used a framework analysis approach

9. Is there a clear statement of findings?

Yes ✔

Can't Tell ☐

No ☐

HINT: Consider whether
- If the findings are explicit
- If there is adequate discussion of the evidence both for and against the researcher's arguments
- If the researcher has discussed the credibility of their findings (e.g. triangulation, respondent validation, more than one analyst)
- If the findings are discussed in relation to the original research question

Comments: The findings have implications for the development and implementation of a national reassessment process for oncology drugs in Canada and potentially in other countries. The study highlights the importance of stakeholder engagement and the need for transparent and evidence-based decision-making in drug reassessmen

CASP
Critical Appraisal
Skills Programme

Section C: Will the results help locally?

10. How valuable is the research?

HINT: Consider
- If the researcher discusses the contribution the study makes to existing knowledge or understanding (e.g. do they consider the findings in relation to current practice or policy, or relevant research-based literature
- If they identify new areas where research is necessary
- If the researchers have discussed whether or how the findings can be transferred to other populations or considered other ways the research may be used

Comments: The research is valuable in that it provides insights into the current state of HTA implementation in the MENA region, as well as identifying areas where improvements can be made to promote greater use and uptake of HTA in this region.
