## Supplementary material for "Real-world data: A systematic literature review on the barriers, challenges, and opportunities associated with their inclusion in the health technology assessment process": S2 Appendix. Quality assessment of included studies: CASP-Qualitative-Checklist-2018_Facey KM, et al, (2020).pdf

Paper for appraisal and reference: Real-world evidence to support Payer/HTA decisions abou

### Section A: Are the results valid?

1. Was there a clear statement of the aims of the research?

Yes ✔
Can't Tell ☐
No ☐

HINT: Consider
- what was the goal of the research
- why it was thought important
- its relevance

Comments: The article clearly states the aim of the research, which is to explore recent policy proposals about the use of real-world evidence (RWE) applied to the specific context of payer/health technology assessment (HTA) decisions about highly innovative technologies and to develop feasible actions that were implementable by each stakeholder group.

2. Is a qualitative methodology appropriate?

Yes ✔
Can't Tell ☐
No ☐

HINT: Consider
- If the research seeks to interpret or illuminate the actions and/or subjective experiences of research participants
- Is qualitative research the right methodology for addressing the research goal

Comments: A qualitative methodology is appropriate for this study, since the aim of the study was to explore recent policy proposals about the use of real-world evidence (RWE) applied to the specific context of payer/health technology assessment (HTA) decisions about highly innovative technologies and to develop feasible actions that were implementable by each stakeholder group. In particular, mixed-methods approach was used in the research, which included workshops, teleconferences, and email consultations involving stakeholders from different sectors.

### Is it worth continuing?

3. Was the research design appropriate to address the aims of the research?

Yes ✔
Can't Tell ☐
No ☐

HINT: Consider
- if the researcher has justified the research design (e.g. have they discussed how they decided which method to use)

Comments: The mixed-methods approach allowed for a comprehensive exploration of the issue and the case study approach provided context-specific insights.

4. Was the recruitment strategy appropriate to the aims of the research?

| | |
|---|---|
| Yes | |
| Can't Tell | ✔ |
| No | |

HINT: Consider
- If the researcher has explained how the participants were selected
- If they explained why the participants they selected were the most appropriate to provide access to the type of knowledge sought by the study
- If there are any discussions around recruitment (e.g. why some people chose not to take part)

Comments: The article does not provide detailed information on the recruitment strategy

5. Was the data collected in a way that addressed the research issue?

| | |
|---|---|
| Yes | ✔ |
| Can't Tell | |
| No | |

HINT: Consider
- If the setting for the data collection was justified
- If it is clear how data were collected (e.g. focus group, semi-structured interview etc.)
- If the researcher has justified the methods chosen
- If the researcher has made the methods explicit (e.g. for interview method, is there an indication of how interviews are conducted, or did they use a topic guide)
- If methods were modified during the study. If so, has the researcher explained how and why
- If the form of data is clear (e.g. tape recordings, video material, notes etc.)
- If the researcher has discussed saturation of data

Comments: The authors used range of methods including presentations, desk-top research, case studies, and stakeholder actions.

6. Has the relationship between researcher and participants been adequately considered?

| | |
|---|---|
| Yes | |
| Can't Tell | ✔ |
| No | |

HINT: Consider

- If the researcher critically examined their own role, potential bias and influence during (a) formulation of the research questions (b) data collection, including sample recruitment and choice of location
- How the researcher responded to events during the study and whether they considered the implications of any changes in the research design

Comments: The authors did not consider the relationship between research and participants; However, the authors did follow the "Bad Gastein" principle of equal representation of all stakeholders and involved representatives from HTA bodies, payers, regulators, government bodies, clinical research organizations, pharmaceutical industry, patient groups, and academia.

### Section B: What are the results?

7. Have ethical issues been taken into consideration?

| | |
|---|---|
| Yes | |
| Can't Tell | |
| No | ✔ |

HINT: Consider

- If there are sufficient details of how the research was explained to participants for the reader to assess whether ethical standards were maintained
- If the researcher has discussed issues raised by the study (e.g. issues around informed consent or confidentiality or how they have handled the effects of the study on the participants during and after the study)
- If approval has been sought from the ethics committee

Comments: The study did not provide detailed information on the ethical considerations.

Critical Appraisal Skills Programme

8. Was the data analysis sufficiently rigorous?

Yes ☐
Can't Tell ☑
No ☐

HINT: Consider
- If there is an in-depth description of the analysis process
- If thematic analysis is used. If so, is it clear how the categories/themes were derived from the data
- Whether the researcher explains how the data presented were selected from the original sample to demonstrate the analysis process
- If sufficient data are presented to support the findings
- To what extent contradictory data are taken into account
- Whether the researcher critically examined their own role, potential bias and influence during analysis and selection of data for presentation

Comments: The authors grouped stakeholder views on RWE in decision making into themes. Also, the article does not provide a detailed description of the data analysis methods used in the research, but it does state that the case studies were discussed and actions were developed and refined by the Taskforce and their networks over several months.

9. Is there a clear statement of findings?

Yes ☑
Can't Tell ☐
No ☐

HINT: Consider whether
- If the findings are explicit
- If there is adequate discussion of the evidence both for and against the researcher's arguments
- If the researcher has discussed the credibility of their findings (e.g. triangulation, respondent validation, more than one analyst)
- If the findings are discussed in relation to the original research question

Comments: The study provided a clear statement of its findings. A set of stakeholder actions for consistent use of RWE in payer/HTA decisions about highly innovative technologies is also included.

Section C: Will the results help locally?

10. How valuable is the research?

HINT: Consider
- If the researcher discusses the contribution the study makes to existing knowledge or understanding (e.g. do they consider the findings in relation to current practice or policy, or relevant research-based literature
- If they identify new areas where research is necessary
- If the researchers have discussed whether or how the findings can be transferred to other populations or considered other ways the research may be used

Comments: The research provides valuable insights into the challenges and opportunities of using RWE in payer/HTA decisions about highly innovative technologies. The stakeholder actions developed through the research have the potential to support the development of more consistent and evidence-based decision-making processes in this area.
