## Supplementary material for "Real-world data: A systematic literature review on the barriers, challenges, and opportunities associated with their inclusion in the health technology assessment process": S2 Appendix. Quality assessment of included studies: CASP-Qualitative-Checklist-2018_Fasseeh H, et al. (2020).pdf

Paper for appraisal and reference: Implementation of Health Technology Assessment in the M

Section A: Are the results valid?

1. Was there a clear statement of the aims of the research?

Yes ✔
Can't Tell ☐
No ☐

HINT: Consider
- what was the goal of the research
- why it was thought important
- its relevance

Comments:

2. Is a qualitative methodology appropriate?

Yes ✔
Can't Tell ☐
No ☐

HINT: Consider
- If the research seeks to interpret or illuminate the actions and/or subjective experiences of research participants
- Is qualitative research the right methodology for addressing the research goal

Comments: Yes, the qualitative research design was appropriate, as the study used a comparative approach to analyze the differences between the current and preferred status of HTA implementation in the MENA region.

Is it worth continuing?

3. Was the research design appropriate to address the aims of the research?

Yes ✔
Can't Tell ☐
No ☐

HINT: Consider
- if the researcher has justified the research design (e.g. have they discussed how they decided which method to use)

Comments: Comparative analysis of differences of current and future approach in HTA

CASP
Critical Appraisal
Skills Programme

4. Was the recruitment strategy appropriate to the aims of the research?

| | |
|---|---|
| Yes | ✔ |
| Can't Tell | |
| No | |

HINT: Consider
- If the researcher has explained how the participants were selected
- If they explained why the participants they selected were the most appropriate to provide access to the type of knowledge sought by the study
- If there are any discussions around recruitment (e.g. why some people chose not to take part)

Comments: The recruitment strategy appears to be appropriate, as the study recruited a sample of experts in HTA and related fields from various countries in the MENA region.

5. Was the data collected in a way that addressed the research issue?

| | |
|---|---|
| Yes | ✔ |
| Can't Tell | |
| No | |

HINT: Consider
- If the setting for the data collection was justified
- If it is clear how data were collected (e.g. focus group, semi-structured interview etc.)
- If the researcher has justified the methods chosen
- If the researcher has made the methods explicit (e.g. for interview method, is there an indication of how interviews are conducted, or did they use a topic guide)
- If methods were modified during the study. If so, has the researcher explained how and why
- If the form of data is clear (e.g. tape recordings, video material, notes etc.)
- If the researcher has discussed saturation of data

Comments: The data were collected through semi-structured interviews that were designed to elicit information about the current and preferred status of HTA implementation in the MENA region.

6. Has the relationship between researcher and participants been adequately considered?
