## Supplementary material for "Real-world data: A systematic literature review on the barriers, challenges, and opportunities associated with their inclusion in the health technology assessment process": S2 Appendix. Quality assessment of included studies: CASP-Qualitative-Checklist-2018_Hampson G, et al, (2018).pdf

Paper for appraisal and reference: Real-world evidence for coverage decisions: opportunities

### Section A: Are the results valid?

1. Was there a clear statement of the aims of the research?

| | |
|---|---|
| Yes | ✔ |
| Can't Tell | |
| No | |

HINT: Consider
- what was the goal of the research
- why it was thought important
- its relevance

Comments: The aim of the research was to gather information and insights on the use of real-world evidence (RWE) for coverage decisions, as well as to identify opportunities and challenges associated with RWE.

2. Is a qualitative methodology appropriate?

| | |
|---|---|
| Yes | ✔ |
| Can't Tell | |
| No | |

HINT: Consider
- If the research seeks to interpret or illuminate the actions and/or subjective experiences of research participants
- Is qualitative research the right methodology for addressing the research goal

Comments: A qualitative methodology was appropriate for this study as it involved semi-structured interviews and a literature review to gather in-depth information and insights from experts in the field.

### Is it worth continuing?

3. Was the research design appropriate to address the aims of the research?

| | |
|---|---|
| Yes | ✔ |
| Can't Tell | |
| No | |

HINT: Consider
- if the researcher has justified the research design (e.g. have they discussed how they decided which method to use)

Comments: The research design was appropriate as it included semi-structured interviews with a diverse range of experts and a literature review to gather information and insights on the use of RWE for coverage decisions, as well as to identify opportunities and challenges associated with RWE.

CASP
Critical Appraisal
Skills Programme

4. Was the recruitment strategy appropriate to the aims of the research?

Yes ✔
Can't Tell ☐
No ☐

HINT: Consider
- If the researcher has explained how the participants were selected
- If they explained why the participants they selected were the most appropriate to provide access to the type of knowledge sought by the study
- If there are any discussions around recruitment (e.g. why some people chose not to take part)

Comments: The recruitment strategy was appropriate as the interviewees were selected based on their expertise and experience in the field of RWE, representing a diverse range of stakeholders including pharmaceutical manufacturers, payers, and academia.

5. Was the data collected in a way that addressed the research issue?

Yes ✔
Can't Tell ☐
No ☐

HINT: Consider
- If the setting for the data collection was justified
- If it is clear how data were collected (e.g. focus group, semi-structured interview etc.)
- If the researcher has justified the methods chosen
- If the researcher has made the methods explicit (e.g. for interview method, is there an indication of how interviews are conducted, or did they use a topic guide)
- If methods were modified during the study. If so, has the researcher explained how and why
- If the form of data is clear (e.g. tape recordings, video material, notes etc.)
- If the researcher has discussed saturation of data

Comments: The data collection methods, including semi-structured interviews and a literature review, were designed to gather information and insights on the use of RWE for coverage decisions and to identify opportunities and challenges associated with RWE.

6. Has the relationship between researcher and participants been adequately considered?

Yes ☐
Can't Tell ☐
No ☑

HINT: Consider
- If the researcher critically examined their own role, potential bias and influence during (a) formulation of the research questions (b) data collection, including sample recruitment and choice of location
- How the researcher responded to events during the study and whether they considered the implications of any changes in the research design

Comments: The paper did not provide detailed information on the relationship between the researchers and participants

Section B: What are the results?

7. Have ethical issues been taken into consideration?

Yes ☐
Can't Tell ☐
No ☑

HINT: Consider
- If there are sufficient details of how the research was explained to participants for the reader to assess whether ethical standards were maintained
- If the researcher has discussed issues raised by the study (e.g. issues around informed consent or confidentiality or how they have handled the effects of the study on the participants during and after the study)
- If approval has been sought from the ethics committee

Comments: The paper did not provide detailed information on the ethical considerations of the study,

CASP
Critical Appraisal
Skills Programme

8. Was the data analysis sufficiently rigorous?

Yes ☑
Can't Tell ☐
No ☐

HINT: Consider
- If there is an in-depth description of the analysis process
- If thematic analysis is used. If so, is it clear how the categories/themes were derived from the data
- Whether the researcher explains how the data presented were selected from the original sample to demonstrate the analysis process
- If sufficient data are presented to support the findings
- To what extent contradictory data are taken into account
- Whether the researcher critically examined their own role, potential bias and influence during analysis and selection of data for presentation

Comments: The authors provide an in-depth description of the analysis process, including the use of a thematic analysis approach and a clear explanation of how the categories/themes were derived from the data.

9. Is there a clear statement of findings?

Yes ☑
Can't Tell ☐
No ☐

HINT: Consider whether
- If the findings are explicit
- If there is adequate discussion of the evidence both for and against the researcher's arguments
- If the researcher has discussed the credibility of their findings (e.g. triangulation, respondent validation, more than one analyst)
- If the findings are discussed in relation to the original research question

Comments: The authors conclude that while RWE has the potential to provide valuable insights for coverage decisions, there are many challenges that need to be addressed in order to make it a reliable and useful tool. These challenges include issues related to data quality, data privacy, and the need for rigorous study designs and methodologies.

### Section C: Will the results help locally?

10. How valuable is the research?

HINT: Consider
- If the researcher discusses the contribution the study makes to existing knowledge or understanding (e.g. do they consider the findings in relation to current practice or policy, or relevant research-based literature
- If they identify new areas where research is necessary
- If the researchers have discussed whether or how the findings can be transferred to other populations or considered other ways the research may be used

Comments: The study provides a comprehensive overview of the opportunities and challenges associated with using RWE to inform coverage decisions. It highlights the need for careful consideration of the strengths and limitations of RWE, as well as the importance of rigorous study designs and methodologies. The study provides useful insights for researchers, policymakers, and other stakeholders who are interested in using RWE to inform coverage decisions.
