## Supplementary material for "Real-world data: A systematic literature review on the barriers, challenges, and opportunities associated with their inclusion in the health technology assessment process": S2 Appendix. Quality assessment of included studies: CASP-Qualitative-Checklist-2018_Hogervorst Milou A, et al, (2022).pdf

Paper for appraisal and reference: Real World Data in Health Technology Assessment of Con

### Section A: Are the results valid?

1. Was there a clear statement of the aims of the research?

Yes ✔
Can't Tell ☐
No ☐

HINT: Consider
- what was the goal of the research
- why it was thought important
- its relevance

Comments: The authors clearly state the aim of the research, which is to gain insight into daily practice in health technology assessment (HTA) organizations by using a questionnaire to collect data on the use of real-world data (RWD) in HTA, as part of the Horizon 2020 HTx project.

2. Is a qualitative methodology appropriate?

Yes ✔
Can't Tell ☐
No ☐

HINT: Consider
- If the research seeks to interpret or illuminate the actions and/or subjective experiences of research participants
- Is qualitative research the right methodology for addressing the research goal

Comments: The study used a mixed-methods approach, combining quantitative data from the questionnaire with qualitative data from open-ended questions. This approach is appropriate for the research aim of gaining insight into daily practice in HTA organizations.

### Is it worth continuing?

3. Was the research design appropriate to address the aims of the research?

Yes ✔
Can't Tell ☐
No ☐

HINT: Consider
- if the researcher has justified the research design (e.g. have they discussed how they decided which method to use)

Comments: A mixed-methods approach using a questionnaire and open-ended questions is appropriate to address the aims of the research, which were to gain insight into the use of RWD in HTA as part of the Horizon 2020 HTx project.

CASP
Critical Appraisal
Skills Programme

4. Was the recruitment strategy appropriate to the aims of the research?

| | |
|---|---|
| Yes | ✔ |
| Can't Tell | |
| No | |

HINT: Consider
- If the researcher has explained how the participants were selected
- If they explained why the participants they selected were the most appropriate to provide access to the type of knowledge sought by the study
- If there are any discussions around recruitment (e.g. why some people chose not to take part)

Comments: The recruitment strategy through dissemination of a questionnaire is appropriate for the research aim of gaining insight into daily practice in HTA organizations.

5. Was the data collected in a way that addressed the research issue?

| | |
|---|---|
| Yes | ✔ |
| Can't Tell | |
| No | |

HINT: Consider
- If the setting for the data collection was justified
- If it is clear how data were collected (e.g. focus group, semi-structured interview etc.)
- If the researcher has justified the methods chosen
- If the researcher has made the methods explicit (e.g. for interview method, is there an indication of how interviews are conducted, or did they use a topic guide)
- If methods were modified during the study. If so, has the researcher explained how and why
- If the form of data is clear (e.g. tape recordings, video material, notes etc.)
- If the researcher has discussed saturation of data

Comments: The data were collected through a questionnaire and open-ended questions, which addressed the research issue of the use of RWD in HTA as part of the Horizon 2020 HTx project.

6. Has the relationship between researcher and participants been adequately considered?

| | |
|---|---|
| Yes | ☐ |
| Can't Tell | ☐ |
| No | ☑ |

HINT: Consider

- If the researcher critically examined their own role, potential bias and influence during (a) formulation of the research questions (b) data collection, including sample recruitment and choice of location
- How the researcher responded to events during the study and whether they considered the implications of any changes in the research design

Comments: The authors did not provide information for this topic.

Section B: What are the results?

7. Have ethical issues been taken into consideration?

| | |
|---|---|
| Yes | ☐ |
| Can't Tell | ☐ |
| No | ☑ |

HINT: Consider

- If there are sufficient details of how the research was explained to participants for the reader to assess whether ethical standards were maintained
- If the researcher has discussed issues raised by the study (e.g. issues around informed consent or confidentiality or how they have handled the effects of the study on the participants during and after the study)
- If approval has been sought from the ethics committee

Comments: The study did not consider ethical issues

CASP Critical Appraisal Skills Programme

8. Was the data analysis sufficiently rigorous?

Yes ✔
Can't Tell ☐
No ☐

HINT: Consider
- If there is an in-depth description of the analysis process
- If thematic analysis is used. If so, is it clear how the categories/themes were derived from the data
- Whether the researcher explains how the data presented were selected from the original sample to demonstrate the analysis process
- If sufficient data are presented to support the findings
- To what extent contradictory data are taken into account
- Whether the researcher critically examined their own role, potential bias and influence during analysis and selection of data for presentation

Comments: The study used a mixed-methods approach and provided a detailed description of the data analysis, which included both quantitative and qualitative analyses. The data analysis was therefore sufficiently rigorous.

9. Is there a clear statement of findings?

Yes ✔
Can't Tell ☐
No ☐

HINT: Consider whether
- If the findings are explicit
- If there is adequate discussion of the evidence both for and against the researcher's arguments
- If the researcher has discussed the credibility of their findings (e.g. triangulation, respondent validation, more than one analyst)
- If the findings are discussed in relation to the original research question

Comments: The study provided a clear statement of findings, which included insights into the use of RWD in HTA as part of the Horizon 2020 HTx project, as well as the benefits and challenges associated with using RWD in HTA.

Critical Appraisal Skills Programme

Section C: Will the results help locally?

10. How valuable is the research?

HINT: Consider
- If the researcher discusses the contribution the study makes to existing knowledge or understanding (e.g. do they consider the findings in relation to current practice or policy, or relevant research-based literature
- If they identify new areas where research is necessary
- If the researchers have discussed whether or how the findings can be transferred to other populations or considered other ways the research may be used

Comments: The research is valuable as it provides insights into the use of RWD in HTA as part of the Horizon 2020 HTx project, and highlights the benefits and challenges associated with using RWD in HTA. These insights can inform future research and practice in the field of HTA.
