## Supplementary material for "Real-world data: A systematic literature review on the barriers, challenges, and opportunities associated with their inclusion in the health technology assessment process": S2 Appendix. Quality assessment of included studies: CASP-Qualitative-Checklist-2018_Husereau, D, et al, (2019).pdf

Paper for appraisal and reference: Use of Real-World Data Sources for Canadian Drug Pricin

### Section A: Are the results valid?

1. Was there a clear statement of the aims of the research?

Yes ☑
Can't Tell ☐
No ☐

HINT: Consider
- what was the goal of the research
- why it was thought important
- its relevance

Comments: There was a clear statement of the aims of the research in the provided text. The primary aim of the research is to "reflect on perceptions of the value of real-world evidence in pricing and reimbursement decisions, barriers to its optimal use in pricing and reimbursement, current initiatives that may lead to its increased use, and what role the pharmaceutical industry may play in this." This research also intends to inform an international discussion on optimizing the use of real-world evidence for pricing and reimbursement decisions.

2. Is a qualitative methodology appropriate?

Yes ☑
Can't Tell ☐
No ☐

HINT: Consider
- If the research seeks to interpret or illuminate the actions and/or subjective experiences of research participants
- Is qualitative research the right methodology for addressing the research goal

Comments: The research methodology outlined in the provided text describes a qualitative approach in which multi-stakeholder meetings were conducted to capture perceptions and discussions about the use of real-world evidence in Canada. Participants from various backgrounds and geographic regions were invited to roundtable discussions, and the data collection involved detailed notes, audio recordings, and transcriptions of discussions. These data were then analyzed qualitatively, with the identification of concepts and categorization into themes.

### Is it worth continuing?

3. Was the research design appropriate to address the aims of the research?

Yes ☑
Can't Tell ☐
No ☐

HINT: Consider
- if the researcher has justified the research design (e.g. have they discussed how they decided which method to use)

Comments: While the text lacks a detailed discussion of the reasoning behind the research design, the chosen methodology, which includes roundtable discussions, note-taking, audio recordings, and qualitative analysis, aligns well with the research's qualitative nature and its aim to explore the challenges, opportunities, and perceptions surrounding real-world evidence in healthcare decision-making in Canada.

4. Was the recruitment strategy appropriate to the aims of the research?

| | |
|---|---|
| Yes | ✔ |
| Can't Tell | |
| No | |

HINT: Consider
- If the researcher has explained how the participants were selected
- If they explained why the participants they selected were the most appropriate to provide access to the type of knowledge sought by the study
- If there are any discussions around recruitment (e.g. why some people chose not to take part)

Comments: A convenience sample of participants were identified according to background roles (i.e., academic/public/private sector) and geography (i.e., across Canada) and invited to participate in roundtable discussions.

5. Was the data collected in a way that addressed the research issue?

| |
|---|
| Yes |
| Can't Tell |
| No |

HINT: Consider
- If the setting for the data collection was justified
- If it is clear how data were collected (e.g. focus group, semi-structured interview etc.)
- If the researcher has justified the methods chosen
- If the researcher has made the methods explicit (e.g. for interview method, is there an indication of how interviews are conducted, or did they use a topic guide)
- If methods were modified during the study. If so, has the researcher explained how and why
- If the form of data is clear (e.g. tape recordings, video material, notes etc.)
- If the researcher has discussed saturation of data

Comments: The data collection methods described in the text appear to be appropriate for addressing the research issue, which is focused on capturing perceptions of opportunities and challenges related to the use of real-world evidence in healthcare decision-making in Canada. The collection allow for the exploration of diverse perspectives and in-depth insights related to the use of real-world evidence in healthcare decision-making in Canada.

6. Has the relationship between researcher and participants been adequately considered?

Yes ☐

Can't Tell ☑

No ☐

HINT: Consider

- If the researcher critically examined their own role, potential bias and influence during (a) formulation of the research questions (b) data collection, including sample recruitment and choice of location
- How the researcher responded to events during the study and whether they considered the implications of any changes in the research design

Comments: No further details and information on this by the Authors.

Section B: What are the results?

7. Have ethical issues been taken into consideration?

Yes ☐

Can't Tell ☐

No ☑

HINT: Consider

- If there are sufficient details of how the research was explained to participants for the reader to assess whether ethical standards were maintained
- If the researcher has discussed issues raised by the study (e.g. issues around informed consent or confidentiality or how they have handled the effects of the study on the participants during and after the study)
- If approval has been sought from the ethics committee

Comments: The study did not provide detailed information on the ethical considerations taken into account during the research process

CASP
Critical Appraisal
Skills Programme

8. Was the data analysis sufficiently rigorous?

| | |
|---|---|
| Yes | ☐ |
| Can't Tell | ☑ |
| No | ☐ |

HINT: Consider
- If there is an in-depth description of the analysis process
- If thematic analysis is used. If so, is it clear how the categories/themes were derived from the data
- Whether the researcher explains how the data presented were selected from the original sample to demonstrate the analysis process
- If sufficient data are presented to support the findings
- To what extent contradictory data are taken into account
- Whether the researcher critically examined their own role, potential bias and influence during analysis and selection of data for presentation

Comments: While the text provides valuable thematic findings related to the barriers and challenges of using real-world data, it lacks some crucial details about the analysis process, including the derivation of themes and the handling of contradictory data. A more in-depth description of the analysis process and the inclusion of data excerpts to support the findings would enhance the rigor of the analysis.

9. Is there a clear statement of findings?

| | |
|---|---|
| Yes | ☑ |
| Can't Tell | ☐ |
| No | ☐ |

HINT: Consider whether
- If the findings are explicit
- If there is adequate discussion of the evidence both for and against the researcher's arguments
- If the researcher has discussed the credibility of their findings (e.g. triangulation, respondent validation, more than one analyst)
- If the findings are discussed in relation to the original research question

Comments: The research provides valuable insights into the use of real-world data in Canada for pricing and eimbursement decisions. It presents explicit findings, discusses evidence both for and against the arguments, and incorporates credibility-enhancing measures like triangulation and respondent validation. The findings are well-connected to the original research question. However, further exploration of counterarguments and perspectives could enhance the depth of the research.

CASP
Critical Appraisal
Skills Programme

Section C: Will the results help locally?

10. How valuable is the research?

HINT: Consider
- If the researcher discusses the contribution the study makes to existing knowledge or understanding (e.g. do they consider the findings in relation to current practice or policy, or relevant research-based literature
- If they identify new areas where research is necessary
- If the researchers have discussed whether or how the findings can be transferred to other populations or considered other ways the research may be used

Comments: The findings of the study are valuable in relation to the original research question as they identify and prioritize key value drivers for pharmaceutical products in HTA in Saudi Arabia, which can inform decision-making processes and improve the efficiency and effectiveness of healthcare systems in the region.
