## Supplementary material for "Real-world data: A systematic literature review on the barriers, challenges, and opportunities associated with their inclusion in the health technology assessment process": S2 Appendix. Quality assessment of included studies: CASP-Qualitative-Checklist-2018_Lou J, et al, (2020).pdf

Paper for appraisal and reference: Real-world data for health technology assessment for reim

### Section A: Are the results valid?

1. Was there a clear statement of the aims of the research?

Yes ✔ | Can't Tell ☐ | No ☐

HINT: Consider
- what was the goal of the research
- why it was thought important
- its relevance

Comments: The aims of the research were clearly stated in the title and throughout the study. The study aimed to assess the current landscape of real-world data (RWD) use in health technology assessment (HTA) for reimbursement decisions in Asia and provide recommendations for its future use.

2. Is a qualitative methodology appropriate?

Yes ✔ | Can't Tell ☐ | No ☐

HINT: Consider
- If the research seeks to interpret or illuminate the actions and/or subjective experiences of research participants
- Is qualitative research the right methodology for addressing the research goal

Comments: A qualitative methodology was appropriate for this study as it involved gathering personal and health system level experiences of using RWD/RWE to inform HTA for reimbursement decisions in eleven health systems in Asia. Qualitative research is often used to explore people's experiences, attitudes, and beliefs.

### Is it worth continuing?

3. Was the research design appropriate to address the aims of the research?

Yes ✔ | Can't Tell ☐ | No ☐

HINT: Consider
- if the researcher has justified the research design (e.g. have they discussed how they decided which method to use)

Comments: The research design was appropriate to address the aims of the research. The study involved three activities: an online survey, a face-to-face meeting, and a teleconference. The study used a mixed-methods approach to gather information from a variety of sources, including leaders and technical staff from HTA agencies, clinicians, and representatives from different health systems.

CASP
Critical Appraisal
Skills Programme

4. Was the recruitment strategy appropriate to the aims of the research?

| | |
|---|---|
| Yes | ✔ |
| Can't Tell | |
| No | |

HINT: Consider
- If the researcher has explained how the participants were selected
- If they explained why the participants they selected were the most appropriate to provide access to the type of knowledge sought by the study
- If there are any discussions around recruitment (e.g. why some people chose not to take part)

Comments: The recruitment strategy was appropriate to the aims of the research. The opportunity to participate was advertised during HTAsiaLink 2019, and responses were voluntary. If a country did not have any responses, contacts from that country in HTAsiaLink were requested to complete the survey.

5. Was the data collected in a way that addressed the research issue?

| | |
|---|---|
| Yes | ✔ |
| Can't Tell | |
| No | |

HINT: Consider
- If the setting for the data collection was justified
- If it is clear how data were collected (e.g. focus group, semi-structured interview etc.)
- If the researcher has justified the methods chosen
- If the researcher has made the methods explicit (e.g. for interview method, is there an indication of how interviews are conducted, or did they use a topic guide)
- If methods were modified during the study. If so, has the researcher explained how and why
- If the form of data is clear (e.g. tape recordings, video material, notes etc.)
- If the researcher has discussed saturation of data

Comments: The data were collected in a way that addressed the research issue. The study used a combination of online surveys, face-to-face meetings, and teleconferences to gather personal and health system level experiences of using RWD/RWE to inform HTA for reimbursement decisions in eleven health systems in Asia.

CASP
Critical Appraisal Skills Programme

6. Has the relationship between researcher and participants been adequately considered?

- [x] Yes
- [ ] Can't Tell
- [ ] No

HINT: Consider
- If the researcher critically examined their own role, potential bias and influence during (a) formulation of the research questions (b) data collection, including sample recruitment and choice of location
- How the researcher responded to events during the study and whether they considered the implications of any changes in the research design

Comments: The relationship between the researcher and participants has been adequately considered. The study followed ethical guidelines and the participants were given informed consent before participating. Chatham House rules were observed during the face-to-face meeting, which allowed for open discussion and ensured anonymity.

### Section B: What are the results?

7. Have ethical issues been taken into consideration?

- [x] Yes
- [ ] Can't Tell
- [ ] No

HINT: Consider
- If there are sufficient details of how the research was explained to participants for the reader to assess whether ethical standards were maintained
- If the researcher has discussed issues raised by the study (e.g. issues around informed consent or confidentiality or how they have handled the effects of the study on the participants during and after the study)
- If approval has been sought from the ethics committee

Comments: Ethical issues have been taken into consideration. The study followed ethical guidelines and the participants were given informed consent before participating. Chatham House rules were observed during the face-to-face meeting, which allowed for open discussion and ensured anonymity.

CASP
Critical Appraisal Skills Programme

8. Was the data analysis sufficiently rigorous?

Yes ✔
Can't Tell
No

HINT: Consider
- If there is an in-depth description of the analysis process
- If thematic analysis is used. If so, is it clear how the categories/themes were derived from the data
- Whether the researcher explains how the data presented were selected from the original sample to demonstrate the analysis process
- If sufficient data are presented to support the findings
- To what extent contradictory data are taken into account
- Whether the researcher critically examined their own role, potential bias and influence during analysis and selection of data for presentation

Comments: The study used a mixed-methods approach to analyze the data, including thematic analysis and descriptive statistics. The researchers explained the analysis process, presented sufficient data to support their findings, and took into account contradictory data.

9. Is there a clear statement of findings?

Yes ✔
Can't Tell
No

HINT: Consider whether
- If the findings are explicit
- If there is adequate discussion of the evidence both for and against the researcher's arguments
- If the researcher has discussed the credibility of their findings (e.g. triangulation, respondent validation, more than one analyst)
- If the findings are discussed in relation to the original research question

Comments: The study identified several challenges to using RWD/RWE for HTA for reimbursement decisions in Asia, including data quality, accessibility, and comparability. The study also provided recommendations for addressing these challenges and promoting the use of RWD/RWE in HTA.

Section C: Will the results help locally?

10. How valuable is the research?

HINT: Consider
- If the researcher discusses the contribution the study makes to existing knowledge or understanding (e.g. do they consider the findings in relation to current practice or policy, or relevant research-based literature
- If they identify new areas where research is necessary
- If the researchers have discussed whether or how the findings can be transferred to other populations or considered other ways the research may be used

Comments: The research is valuable as it provides insights into the current landscape of using RWD/RWE for HTA for reimbursement decisions in Asia, identifies challenges and opportunities for improving the use of RWD/RWE, and provides recommendations for future research and policy development. The study can inform decision-making processes and contribute to the advancement of evidence-based healthcare policies in the region.
