## Supplementary material for "Real-world data: A systematic literature review on the barriers, challenges, and opportunities associated with their inclusion in the health technology assessment process": S2 Appendix. Quality assessment of included studies: CASP-Qualitative-Checklist-2018_Makady A, et al, (2017).pdf

CASP
Critical Appraisal
Skills Programme

www.casp-uk.net

Summertown Pavilion, Middle
Way Oxford OX2 7LG

### CASP Checklist: 10 questions to help you make sense of a Qualitative research

**How to use this appraisal tool:** Three broad issues need to be considered when appraising a qualitative study:

- Are the results of the study valid? (Section A)
- What are the results? (Section B)
- Will the results help locally? (Section C)

The 10 questions on the following pages are designed to help you think about these issues systematically. The first two questions are screening questions and can be answered quickly. If the answer to both is "yes", it is worth proceeding with the remaining questions. There is some degree of overlap between the questions, you are asked to record a "yes", "no" or "can't tell" to most of the questions. A number of italicised prompts are given after each question. These are designed to remind you why the question is important. Record your reasons for your answers in the spaces provided.

**About:** These checklists were designed to be used as educational pedagogic tools, as part of a workshop setting, therefore we do not suggest a scoring system. The core CASP checklists (randomised controlled trial & systematic review) were based on JAMA 'Users' guides to the medical literature 1994 (adapted from Guyatt GH, Sackett DL, and Cook DJ), and piloted with health care practitioners.

For each new checklist, a group of experts were assembled to develop and pilot the checklist and the workshop format with which it would be used. Over the years overall adjustments have been made to the format, but a recent survey of checklist users reiterated that the basic format continues to be useful and appropriate.

**Referencing:** we recommend using the Harvard style citation, i.e.: *Critical Appraisal Skills Programme (2018). CASP (insert name of checklist i.e. Qualitative) Checklist. [online] Available at: URL. Accessed: Date Accessed.*

©CASP this work is licensed under the Creative Commons Attribution – Non-Commercial-Share A like. To view a copy of this license, visit http://creativecommons.org/licenses/by-nc-sa/3.0/ www.casp-uk.net

Paper for appraisal and reference: Policies for Use of Real-World Data in Health Technology

#### Section A: Are the results valid?

1. Was there a clear statement of the aims of the research?

| | |
|---|---|
| Yes | ✔ |
| Can't Tell | |
| No | |

HINT: Consider
- what was the goal of the research
- why it was thought important
- its relevance

Comments: The main objectives of the research are to review and compare the policies of six European Health Technology Assessment (HTA) agencies regarding the use of Real-World Data (RWD) in Relative Effectiveness Assessments (REA) of drugs. The research aims to investigate the context of RWD use in REA, including initial reimbursement discussions, pharmacoeconomic analyses, and conditional reimbursement schemes. The study also intends to identify variations in policies among different HTA agencies and highlight the need for more alignment in policies to facilitate the use of RWD in HTA across Europe. The research is focused on the policies of six HTA agencies and does not aim to provide an exhaustive overview of all European jurisdictions.

2. Is a qualitative methodology appropriate?

| | |
|---|---|
| Yes | ✔ |
| Can't Tell | |
| No | |

HINT: Consider
- If the research seeks to interpret or illuminate the actions and/or subjective experiences of research participants
- Is qualitative research the right methodology for addressing the research goal

Comments: The qualitative methodology employed in this study is appropriate for the research objectives. The study utilizes a mixed-methods approach that combines document reviews, academic literature searches, and semi-structured interviews to gather information on the policies of six European Health Technology Assessment (HTA) agencies regarding the use of Real-World Data (RWD) in Relative Effectiveness Assessments (REA) of drugs. The qualitative methods used, including interviews and document analysis, are well-suited for exploring the policies, practices, and perspectives of these agencies. This approach allows for in-depth data collection and analysis, which aligns with the complexity of the research topic and objectives.

#### Is it worth continuing?

3. Was the research design appropriate to address the aims of the research?

| | |
|---|---|
| Yes | ✔ |
| Can't Tell | |
| No | |

HINT: Consider
- if the researcher has justified the research design (e.g. have they discussed how they decided which method to use)

Comments: The study aimed to review the policies of six European Health Technology Assessment (HTA) agencies regarding the use of Real-World Data (RWD) in Relative Effectiveness Assessments (REA) of drugs. To achieve this, the researchers used a combination of methods, including document reviews, academic literature searches, and semistructured interviews with representatives from the selected agencies.

4. Was the recruitment strategy appropriate to the aims of the research?

| Answer | |
|---|---|
| Yes | ☐ |
| Can't Tell | ✔ |
| No | ☐ |

HINT: Consider
- If the researcher has explained how the participants were selected
- If they explained why the participants they selected were the most appropriate to provide access to the type of knowledge sought by the study
- If there are any discussions around recruitment (e.g. why some people chose not to take part)

Comments: The authors did not indicate clearly the recruitment strategy; although semistructured interviews were conducted with representatives from the selected HTA agencies, no further information regarding how the recruitment strategy implemented was mentioned within the manuscrupt.

5. Was the data collected in a way that addressed the research issue?

| Answer | |
|---|---|
| Yes | ✔ |
| Can't Tell | ☐ |
| No | ☐ |

HINT: Consider
- If the setting for the data collection was justified
- If it is clear how data were collected (e.g. focus group, semi-structured interview etc.)
- If the researcher has justified the methods chosen
- If the researcher has made the methods explicit (e.g. for interview method, is there an indication of how interviews are conducted, or did they use a topic guide)
- If methods were modified during the study. If so, has the researcher explained how and why
- If the form of data is clear (e.g. tape recordings, video material, notes etc.)
- If the researcher has discussed saturation of data

Comments: The researchers employed a combination of strategies to gather comprehensive information on the policies of six European Health Technology Assessment (HTA) agencies regarding the use of Real-World Data (RWD) in Relative Effectiveness Assessments (REA) of drugs. Here are the key points that demonstrate the appropriateness of the data collection methods: 1) The selection of six European HTA agencies, including the "Big Four" and pioneering agencies, was well-justified. These agencies represented a diverse range of European jurisdictions and had significant influence on health policies. This diversity ensured that the data collected would be comprehensive and reflective of different HTA approaches. 2) The methods used for data collection were clearly outlined. The researchers conducted a review of agency guidelines and policy papers, a review of academic publications, and semistructured interviews. The research issue, which was to review policies, was aligned with these data collection methods.

These are only few of several reasons for selecting and managing the data collected in order to answer the research issue.

6. Has the relationship between researcher and participants been adequately considered?

| | |
|---|---|
| Yes | |
| Can’t Tell | |
| No | ✔ |

HINT: Consider

- If the researcher critically examined their own role, potential bias and influence during (a) formulation of the research questions (b) data collection, including sample recruitment and choice of location
- How the researcher responded to events during the study and whether they considered the implications of any changes in the research design

Comments: The authors did not consider the relationship between research and participants

Section B: What are the results?

7. Have ethical issues been taken into consideration?

| | |
|---|---|
| Yes | |
| Can’t Tell | |
| No | ✔ |

HINT: Consider

- If there are sufficient details of how the research was explained to participants for the reader to assess whether ethical standards were maintained
- If the researcher has discussed issues raised by the study (e.g. issues around informed consent or confidentiality or how they have handled the effects of the study on the participants during and after the study)
- If approval has been sought from the ethics committee

Comments: The study did not provide information on the ethical considerations taken into account during the research process

8. Was the data analysis sufficiently rigorous?

| | |
|---|---|
| Yes | ✔ |
| Can't Tell | |
| No | |

HINT: Consider
- If there is an in-depth description of the analysis process
- If thematic analysis is used. If so, is it clear how the categories/themes were derived from the data
- Whether the researcher explains how the data presented were selected from the original sample to demonstrate the analysis process
- If sufficient data are presented to support the findings
- To what extent contradictory data are taken into account
- Whether the researcher critically examined their own role, potential bias and influence during analysis and selection of data for presentation

Comments: The manuscript follows a structured and rigorous approach to data collection and analysis. It provides a clear description of the methods used for both data collection and analysis, ensuring transparency and replicability. The mention of inter-rater reliability and the resolution of discrepancies through consensus demonstrate a rigorous approach to the data analysis process. Therefore, the data analysis appears to be sufficiently rigorous based on the information provided in the manuscript. However, a more detailed description of the coding process and themes derived from the data would enhance the understanding of the analysis.

9. Is there a clear statement of findings?

| | |
|---|---|
| Yes | ✔ |
| Can't Tell | |
| No | |

HINT: Consider whether
- If the findings are explicit
- If there is adequate discussion of the evidence both for and against the researcher's arguments
- If the researcher has discussed the credibility of their findings (e.g. triangulation, respondent validation, more than one analyst)
- If the findings are discussed in relation to the original research question

Comments: The findings are explicit, and the manuscript discusses the variations in policies across agencies, the implications for MAHs, and the need for harmonization. It also points out the limitations of the study and provides recommendations for addressing the issues identified. Overall, the findings are clearly presented and discussed in relation to the original research questions.

Section C: Will the results help locally?

10. How valuable is the research?

HINT: Consider
- If the researcher discusses the contribution the study makes to existing knowledge or understanding (e.g. do they consider the findings in relation to current practice or policy, or relevant research-based literature
- If they identify new areas where research is necessary
- If the researchers have discussed whether or how the findings can be transferred to other populations or considered other ways the research may be used

Comments: The research provides valuable information for stakeholders in the field of pharmaceuticals, HTA, and healthcare policy. It highlights the need for better alignment of RWD policies and emphasizes the practical challenges that these variations pose for both regulatory authorities and MAHs. The recommendations for collaboration and reference to the EUnetHTA further enhance the research's value by pointing toward potential solutions and initiatives for addressing the identified issues.
