## Supplementary material for "Real-world data: A systematic literature review on the barriers, challenges, and opportunities associated with their inclusion in the health technology assessment process": S2 Appendix. Quality assessment of included studies: CASP-Qualitative-Checklist-2018_Sievers H, et al, (2021).pdf

Paper for appraisal and reference: Real-world evidence: perspectives on challenges, value, a

Section A: Are the results valid?

1. Was there a clear statement of the aims of the research?

Yes ✔
Can't Tell ☐
No ☐

HINT: Consider
- what was the goal of the research
- why it was thought important
- its relevance

Comments:

2. Is a qualitative methodology appropriate?

Yes ✔
Can't Tell ☐
No ☐

HINT: Consider
- If the research seeks to interpret or illuminate the actions and/or subjective experiences of research participants
- Is qualitative research the right methodology for addressing the research goal

Comments: Qualitative methodology was appropriate for the research as the study aimed to explore the perspectives and experiences of stakeholders on real-world evidence.

Is it worth continuing?

3. Was the research design appropriate to address the aims of the research?

Yes ✔
Can't Tell ☐
No ☐

HINT: Consider
- if the researcher has justified the research design (e.g. have they discussed how they decided which method to use)

Comments: The authors used a qualitative research design, specifically a thematic analysis of semi-structured interviews, to address the aims of the research.

4. Was the recruitment strategy appropriate to the aims of the research?

| | |
|---|---|
| Yes | ✔ |
| Can't Tell | |
| No | |

HINT: Consider
- If the researcher has explained how the participants were selected
- If they explained why the participants they selected were the most appropriate to provide access to the type of knowledge sought by the study
- If there are any discussions around recruitment (e.g. why some people chose not to take part)

Comments: The authors described the recruitment strategy in detail, and it was appropriate to the aims of the research as they recruited participants with diverse backgrounds and experiences related to real-world evidence.

5. Was the data collected in a way that addressed the research issue?

| | |
|---|---|
| Yes | ✔ |
| Can't Tell | |
| No | |

HINT: Consider
- If the setting for the data collection was justified
- If it is clear how data were collected (e.g. focus group, semi-structured interview etc.)
- If the researcher has justified the methods chosen
- If the researcher has made the methods explicit (e.g. for interview method, is there an indication of how interviews are conducted, or did they use a topic guide)
- If methods were modified during the study. If so, has the researcher explained how and why
- If the form of data is clear (e.g. tape recordings, video material, notes etc.)
- If the researcher has discussed saturation of data

Comments: Data collected through semi-structured interviews, which were designed to address the research issue of exploring stakeholders' perspectives on real-world evidence.

CASP
Critical Appraisal
Skills Programme

6. Has the relationship between researcher and participants been adequately considered?

Yes ✔
Can't Tell ☐
No ☐

HINT: Consider

- If the researcher critically examined their own role, potential bias and influence during (a) formulation of the research questions (b) data collection, including sample recruitment and choice of location
- How the researcher responded to events during the study and whether they considered the implications of any changes in the research design

Comments: The authors discussed the importance of building trust and rapport with the participants, and they described how they maintained ethical considerations throughout the research process.

Section B: What are the results?

7. Have ethical issues been taken into consideration?

Yes ✔
Can't Tell ☐
No ☐

HINT: Consider

- If there are sufficient details of how the research was explained to participants for the reader to assess whether ethical standards were maintained
- If the researcher has discussed issues raised by the study (e.g. issues around informed consent or confidentiality or how they have handled the effects of the study on the participants during and after the study)
- If approval has been sought from the ethics committee

Comments: According to the authors, ethical approval was obtained from the ethics assessors of Maastricht University and the research was classified as a low-risk project.

8. Was the data analysis sufficiently rigorous?

Yes ☑
Can't Tell ☐
No ☐

HINT: Consider
- If there is an in-depth description of the analysis process
- If thematic analysis is used. If so, is it clear how the categories/themes were derived from the data
- Whether the researcher explains how the data presented were selected from the original sample to demonstrate the analysis process
- If sufficient data are presented to support the findings
- To what extent contradictory data are taken into account
- Whether the researcher critically examined their own role, potential bias and influence during analysis and selection of data for presentation

Comments: A rigorous thematic analysis approach, which involved multiple researchers independently coding the data, discussing and refining the codes, and conducting member checking to validate the findings.

9. Is there a clear statement of findings?

Yes ☑
Can't Tell ☐
No ☐

HINT: Consider whether
- If the findings are explicit
- If there is adequate discussion of the evidence both for and against the researcher's arguments
- If the researcher has discussed the credibility of their findings (e.g. triangulation, respondent validation, more than one analyst)
- If the findings are discussed in relation to the original research question

Comments: A rigorous thematic analysis approach, which involved multiple researchers independently coding the data, discussing and refining the codes, and conducting member checking to validate the findings.

CASP
Critical Appraisal
Skills Programme

### Section C: Will the results help locally?

10. How valuable is the research?

HINT: Consider
- If the researcher discusses the contribution the study makes to existing knowledge or understanding (e.g. do they consider the findings in relation to current practice or policy, or relevant research-based literature
- If they identify new areas where research is necessary
- If the researchers have discussed whether or how the findings can be transferred to other populations or considered other ways the research may be used

Comments: A rigorous thematic analysis approach, which involved multiple researchers independently coding the data, discussing and refining the codes, and conducting member checking to validate the findings.
