## Supplementary material for "Real-world data: A systematic literature review on the barriers, challenges, and opportunities associated with their inclusion in the health technology assessment process": S2 Appendix. Quality assessment of included studies: CASP-Qualitative-Checklist-2018_Timbie JW, et al, (2021).pdf

Paper for appraisal and reference: Use of Real-World Evidence for Regulatory Approval and (

### Section A: Are the results valid?

| | | | |
|---|---|---|---|
| 1. Was there a clear statement of the aims of the research? | Yes | ✔ | HINT: Consider<br>• what was the goal of the research<br>• why it was thought important<br>• its relevance |
| | Can't Tell | | |
| | No | | |

Comments: The article clearly states the aim of the research, which is to conduct a landscape assessment of the use of real-world evidence (RWE) for regulatory approval and coverage of medical devices, and to identify challenges and potential solutions to the use of RWE in these contexts.

| | | | |
|---|---|---|---|
| 2. Is a qualitative methodology appropriate? | Yes | ✔ | HINT: Consider<br>• If the research seeks to interpret or illuminate the actions and/or subjective experiences of research participants<br>• Is qualitative research the right methodology for addressing the research goal |
| | Can't Tell | | |
| | No | | |

Comments: It is appropriate for for the aim of the study, as it involves conducting interviews with stakeholders to gain an in-depth understanding of their perspectives and experiences with the use of RWE for regulatory approval and coverage of medical devices.

### Is it worth continuing?

| | | | |
|---|---|---|---|
| 3. Was the research design appropriate to address the aims of the research? | Yes | ✔ | HINT: Consider<br>• if the researcher has justified the research design (e.g. have they discussed how they decided which method to use) |
| | Can't Tell | | |
| | No | | |

Comments: The research design is appropriate as it involves conducting interviews with stakeholders representing different groups to gain insights into the challenges and potential solutions related to the use of RWE for regulatory approval and coverage of medical devices.

4. Was the recruitment strategy appropriate to the aims of the research?

| | |
|---|---|
| Yes | ✔ |
| Can't Tell | |
| No | |

HINT: Consider
- If the researcher has explained how the participants were selected
- If they explained why the participants they selected were the most appropriate to provide access to the type of knowledge sought by the study
- If there are any discussions around recruitment (e.g. why some people chose not to take part)

Comments: Tthe recruitment strategy is appropriate as it involved purposive sampling of stakeholders representing different groups, such as the FDA, medical device manufacturers, and payers or health technology assessment organizations, to gain a comprehensive understanding of the issue at hand.

5. Was the data collected in a way that addressed the research issue?

| | |
|---|---|
| Yes | ✔ |
| Can't Tell | |
| No | |

HINT: Consider
- If the setting for the data collection was justified
- If it is clear how data were collected (e.g. focus group, semi-structured interview etc.)
- If the researcher has justified the methods chosen
- If the researcher has made the methods explicit (e.g. for interview method, is there an indication of how interviews are conducted, or did they use a topic guide)
- If methods were modified during the study. If so, has the researcher explained how and why
- If the form of data is clear (e.g. tape recordings, video material, notes etc.)
- If the researcher has discussed saturation of data

Comments: The data collection involved conducting interviews with stakeholders to gather their perspectives and experiences related to the use of RWE for regulatory approval and coverage of medical devices, which directly addresses the research issue.

CASP
Critical Appraisal
Skills Programme

6. Has the relationship between researcher and participants been adequately considered?

| | |
|---|---|
| Yes | ✔ |
| Can't Tell | |
| No | |

HINT: Consider

- If the researcher critically examined their own role, potential bias and influence during (a) formulation of the research questions (b) data collection, including sample recruitment and choice of location
- How the researcher responded to events during the study and whether they considered the implications of any changes in the research design

Comments: All but 2 interviews were recorded with participants ' consent and were professionally transcribed.

Section B: What are the results?

7. Have ethical issues been taken into consideration?

| | |
|---|---|
| Yes | ✔ |
| Can't Tell | |
| No | |

HINT: Consider

- If there are sufficient details of how the research was explained to participants for the reader to assess whether ethical standards were maintained
- If the researcher has discussed issues raised by the study (e.g. issues around informed consent or confidentiality or how they have handled the effects of the study on the participants during and after the study)
- If approval has been sought from the ethics committee

Comments: The article states that all interviews were conducted with participants ' consent and were professionally transcribed. Additionally, the article mentions that ethical considerations were taken into account, such as maintaining confidentiality and anonymity of participants.

Critical Appraisal Skills Programme

8. Was the data analysis sufficiently rigorous?

| | |
|---|---|
| Yes | ✔ |
| Can't Tell | |
| No | |

HINT: Consider
- If there is an in-depth description of the analysis process
- If thematic analysis is used. If so, is it clear how the categories/themes were derived from the data
- Whether the researcher explains how the data presented were selected from the original sample to demonstrate the analysis process
- If sufficient data are presented to support the findings
- To what extent contradictory data are taken into account
- Whether the researcher critically examined their own role, potential bias and influence during analysis and selection of data for presentation

Comments: A rigorous process of thematic coding and iterative refinement of themes is described based on the interview data to ensure that all relevant themes were included in the findings document.

9. Is there a clear statement of findings?

| | |
|---|---|
| Yes | ✔ |
| Can't Tell | |
| No | |

HINT: Consider whether
- If the findings are explicit
- If there is adequate discussion of the evidence both for and against the researcher's arguments
- If the researcher has discussed the credibility of their findings (e.g. triangulation, respondent validation, more than one analyst)
- If the findings are discussed in relation to the original research question

Comments: Clear findings based on the thematic analysis of the interview data were presented, highlighting the challenges and potential solutions related to the use of RWE for regulatory approval and coverage of medical devices.

### Section C: Will the results help locally?

10. How valuable is the research?

HINT: Consider
- If the researcher discusses the contribution the study makes to existing knowledge or understanding (e.g. do they consider the findings in relation to current practice or policy, or relevant research-based literature
- If they identify new areas where research is necessary
- If the researchers have discussed whether or how the findings can be transferred to other populations or considered other ways the research may be used

Comments: The research provides valuable insights from stakeholders representing different groups, including the FDA, medical device manufacturers, and payers or health technology assessment organizations. By conducting interviews with these stakeholders, the researchers were able to gather information on the challenges and opportunities of using RWE for regulatory approval and coverage of medical devices. The findings of this research can be particularly valuable for policymakers and decision-makers in the healthcare industry, as they can inform the development of policies and guidelines related to the use of RWE for medical devices. By understanding the perspectives of different stakeholders, policymakers can make more informed decisions and ensure that the use of RWE is aligned with the needs and interests of all stakeholders involved. Overall, the research provides a valuable contribution to the field of health technology assessment and the use of RWE, and can serve as a starting point for further research and discussions in this area.
