## Supplementary material for "Real-world data: A systematic literature review on the barriers, challenges, and opportunities associated with their inclusion in the health technology assessment process": S2 Appendix. Quality assessment of included studies: Checklist_for_Systematic_Reviews_and_Research_Syntheses_Bowrin K, et al, (2019).pdf

Critical Appraisal tools for use in JBI Systematic Reviews



CRICOS Provider Number 00123M

### INTRODUCTION

JBI is an international research organisation based in the Faculty of Health and Medical Sciences at the University of Adelaide, South Australia. JBI develops and delivers unique evidence-based information, software, education and training designed to improve healthcare practice and health outcomes. With over 70 Collaborating Entities, servicing over 90 countries, JBI is a recognised global leader in evidence-based healthcare.

#### JBI Systematic Reviews

The core of evidence synthesis is the systematic review of literature of a particular intervention, condition or issue. The systematic review is essentially an analysis of the available literature (that is, evidence) and a judgment of the effectiveness or otherwise of a practice, involving a series of complex steps. JBI takes a particular view on what counts as evidence and the methods utilised to synthesise those different types of evidence. In line with this broader view of evidence, JBI has developed theories, methodologies and rigorous processes for the critical appraisal and synthesis of these diverse forms of evidence in order to aid in clinical decision-making in healthcare. There now exists JBI guidance for conducting reviews of effectiveness research, qualitative research, prevalence/incidence, etiology/risk, economic evaluations, text/opinion, diagnostic test accuracy, mixed-methods, umbrella reviews and scoping reviews. Further information regarding JBI systematic reviews can be found in the JBI Evidence Synthesis Manual.

#### JBI Critical Appraisal Tools

All systematic reviews incorporate a process of critique or appraisal of the research evidence. The purpose of this appraisal is to assess the methodological quality of a study and to determine the extent to which a study has addressed the possibility of bias in its design, conduct and analysis. All papers selected for inclusion in the systematic review (that is – those that meet the inclusion criteria described in the protocol) need to be subjected to rigorous appraisal by two critical appraisers. The results of this appraisal can then be used to inform synthesis and interpretation of the results of the study. JBI Critical appraisal tools have been developed by the JBI and collaborators and approved by the JBI Scientific Committee following extensive peer review. Although designed for use in systematic reviews, JBI critical appraisal tools can also be used when creating Critically Appraised Topics (CAT), in journal clubs and as an educational tool.

© JBI, 2020. All rights reserved. JBI grants use of these tools for research purposes only. All other enquiries should be sent to.

### JBI CRITICAL APPRAISAL CHECKLIST FOR SYSTEMATIC REVIEWS AND RESEARCH SYNTHESES

Reviewer: Konstantinos Zisis Date: 21 Apr 2023

Author: Bowrin K, et al Year: 2019 Record Number: N/A

| | | Yes | No | Unclear | Not applicable |
|---|---|---|---|---|---|
| 1. | Is the review question clearly and explicitly stated? | ✓ | ☐ | ☐ | ☐ |
| 2. | Were the inclusion criteria appropriate for the review question? | ✓ | ☐ | ☐ | ☐ |
| 3. | Was the search strategy appropriate? | ✓ | ☐ | ☐ | ☐ |
| 4. | Were the sources and resources used to search for studies adequate? | ✓ | ☐ | ☐ | ☐ |
| 5. | Were the criteria for appraising studies appropriate? | ☐ | ☐ | ✓ | ☐ |
| 6. | Was critical appraisal conducted by two or more reviewers independently? | ☐ | ✓ | ☐ | ☐ |
| 7. | Were there methods to minimize errors in data extraction? | ✓ | ☐ | ☐ | ☐ |
| 8. | Were the methods used to combine studies appropriate? | ✓ | ☐ | ☐ | ☐ |
| 9. | Was the likelihood of publication bias assessed? | ☐ | ✓ | ☐ | ☐ |
| 10. | Were recommendations for policy and/or practice supported by the reported data? | ✓ | ☐ | ☐ | ☐ |
| 11. | Were the specific directives for new research appropriate? | ☐ | ☐ | ✓ | ☐ |

Overall appraisal: Include ✓ Exclude ☐ Seek further info ☐

Comments (Including reason for exclusion)

THE ARTICLE APPEARS TO BE A WELL-WRITTEN AND INFORMATIVE REVIEW OF THE USE OF REAL-WORLD EVIDENCE IN HEALTH TECHNOLOGY ASSESSMENTS.

© JBI, 2020. All rights reserved. JBI grants use of these tools for research purposes only. All other enquiries should be sent to.

### JBI CRITICAL APPRAISAL CHECKLIST FOR SYSTEMATIC REVIEWS AND RESEARCH SYNTHESIS

How to cite: *Aromataris E, Fernandez R, Godfrey C, Holly C, Kahlil H, Tungpunkom P. Summarizing systematic reviews: methodological development, conduct and reporting of an Umbrella review approach. Int J Evid Based Healthc. 2015;13(3):132-40.*

When conducting an umbrella review using the JBI method, the critical appraisal instrument for Systematic Reviews should be used.

The primary and secondary reviewer should discuss each item in the appraisal instrument for each study included in their review. In particular, discussions should focus on what is considered acceptable to the aims of the review in terms of the specific study characteristics. When appraising systematic reviews this discussion may include issues such as what represents an adequate search strategy or appropriate methods of synthesis. The reviewers should be clear on what constitutes acceptable levels of information to allocate a positive appraisal compared with a negative, or response of "unclear". This discussion should ideally take place before the reviewers independently conduct the appraisal.

Within umbrella reviews, quantitative or qualitative systematic reviews may be incorporated, as well as meta-analyses of existing research. There are 11 questions to guide the appraisal of systematic reviews or meta-analyses. Each question should be answered as "yes", "no", or "unclear". Not applicable "NA" is also provided as an option and may be appropriate in rare instances.

##### 1. Is the review question clearly and explicitly stated?

The review question is an essential step in the systematic review process. A well-articulated question defines the scope of the review and aids in the development of the search strategy to locate the relevant evidence. An explicitly stated question, formulated around its PICO (Population, Intervention, Comparator, Outcome) elements aids both the review team in the conduct of the review and the reader in determining if the review has achieved its objectives. Ideally the review question should be articulated in a published protocol; however this will not always be the case with many reviews that are located.

##### 2. Were the inclusion criteria appropriate for the review question?

The inclusion criteria should be identifiable from, and match the review question. The necessary elements of the PICO should be explicit and clearly defined. The inclusion criteria should be detailed and the included reviews should clearly be eligible when matched against the stated inclusion criteria. Appraisers of meta-analyses will find that inclusion criteria may include criteria around the ability to conduct statistical analyses which would not be the norm for a systematic review. The types of included studies should be relevant to the review question, for example, an umbrella review aiming to summarize a range of effective non-pharmacological interventions for aggressive behaviors amongst elderly patients with dementia will limit itself to including systematic reviews and meta-analyses that synthesize quantitative studies assessing the various interventions; qualitative or economic reviews would not be included.

##### 3. Was the search strategy appropriate?

A systematic review should provide evidence of the search strategy that has been used to locate the evidence. This may be found in the methods section of the review report in some cases, or as an appendix that may be provided as supplementary information to the review publication. A systematic review should present a clear search strategy that addresses each of the identifiable PICO components of the review question. Some reviews may also provide a description of the approach to searching and how the terms that were ultimately used were derived, though due to limits on word counts in journals this may be more the norm in online only publications. There

© JBI, 2020. All rights reserved. JBI grants use of these tools for research purposes only. All other enquiries should be sent to.

should be evidence of logical and relevant keywords and terms and also evidence that Subject

Headings and Indexing terms have been used in the conduct of the search. Limits on the search should also be considered and their potential impact; for example, if a date limit was used, was this appropriate and/or justified? If only English language studies were included, will such a language bias have an impact on the review? The response to these considerations will depend, in part, on the review question.

##### 4. Were the sources and resources used to search for studies adequate?

A systematic review should attempt to identify "all" the available evidence and as such there should be evidence of a comprehensive search strategy. Multiple electronic databases should be searched including major bibliographic citation databases such as MEDLINE and CINAHL. Ideally, other databases that are relevant to the review question should also be searched, for example, a systematic review with a question about a physical therapy intervention should also look to search the PEDro database, whilst a review focusing on an educational intervention should also search the ERIC. Reviews of effectiveness should aim to search trial registries. A comprehensive search is the ideal way to minimize publication bias, as a result, a well conducted systematic review should also attempt to search for grey literature, or "unpublished" studies; this may involve searching websites relevant to the review question, or thesis repositories.

##### 5. Were the criteria for appraising studies appropriate?

The systematic review should present a clear statement that critical appraisal was conducted and provide the details of the items that were used to assess the included studies. This may be presented in the methods of the review, as an appendix of supplementary information, or as a reference to a source that can be located. The tools or instruments used should be appropriate for the review question asked and the type of research conducted. For example, a systematic review of effectiveness should present a tool or instrument that addresses aspects of validity for experimental studies and randomized controlled trials such as randomization and blinding – if the review includes observational research to answer the same question a different tool would be more appropriate. Similarly, a review assessing diagnostic test accuracy may refer to the recognized QUADAS[1] tool.

##### 6. Was critical appraisal conducted by two or more reviewers independently?

Critical appraisal or some similar assessment of the quality of the literature included in a systematic review is essential. A key characteristic to minimize bias or systematic error in the conduct of a systematic review is to have the critical appraisal of the included studies completed independently and in duplicate by members of the review team. The systematic review should present a clear statement that critical appraisal was conducted by at least two reviewers working independently from each other and conferring where necessary to reach decision regarding study quality and eligibility on the basis of quality.

##### 7. Were there methods to minimize errors in data extraction?

Efforts made by review authors during data extraction can also minimize bias or systematic errors in the conduct of a systematic review. Strategies to minimize bias may include conducting all data extraction in duplicate and independently, using specific tools or instruments to guide data extraction and some evidence of piloting or training around their use.

##### 8. Were the methods used to combine studies appropriate?

A synthesis of the evidence is a key feature of a systematic review. The synthesis that is presented should be appropriate for the review question and the stated type of systematic review and evidence it refers to. If a meta-analysis has been conducted this needs to be reviewed carefully.

© JBI, 2020. All rights reserved. JBI grants use of these tools for research purposes only. All other enquiries should be sent to.

Was it appropriate to combine the studies? Have the reviewers assessed heterogeneity statistically and provided some explanation for heterogeneity that may be present? Often, where heterogeneous studies are included in the systematic review, narrative synthesis will be an appropriate method for presenting the results of multiple studies. If a qualitative review, are the methods that have been used to synthesize findings congruent with the stated methodology of the review? Is there adequate descriptive and explanatory information to support the final synthesized findings that have been constructed from the findings sourced from the original research?

##### 9. Was the likelihood of publication bias assessed?

As mentioned, a comprehensive search strategy is the best means by which a review author may alleviate the impact of publication bias on the results of the review. Reviews may also present statistical tests such as Egger's test or funnel plots to also assess the potential presence of publication bias and its potential impact on the results of the review. This question will not be applicable to systematic reviews of qualitative evidence.

##### 10. Were recommendations for policy and/or practice supported by the reported data?

Whilst the first nine (9) questions specifically look to identify potential bias in the conduct of a systematic review, the final questions are more indictors of review quality rather than validity. Ideally a review should present recommendations for policy and practice. Where these recommendations are made there should be a clear link to the results of the review. Is there evidence that the strength of the findings and the quality of the research been considered in the formulation of review recommendations?

##### 11. Were the specific directives for new research appropriate?

The systematic review process is recognized for its ability to identify where gaps in the research, or knowledge base, around a particular topic exist. Most systematic review authors will provide some indication, often in the discussion section of the report, of where future research direction should lie. Where evidence is scarce or sample sizes that support overall estimates of effect are small and effect estimates are imprecise, repeating similar research to those identified by the review may be necessary and appropriate. In other instances, the case for new research questions to investigate the topic may be warranted.



© JBI, 2020. All rights reserved. JBI grants use of these tools for research purposes only. All other enquiries should be sent to.
