## Supplementary material for "Real-world data: A systematic literature review on the barriers, challenges, and opportunities associated with their inclusion in the health technology assessment process": S2 Appendix. Quality assessment of included studies: Checklist_for_Systematic_Reviews_and_Research_Syntheses_Brogaard N, et al, (2021).pdf

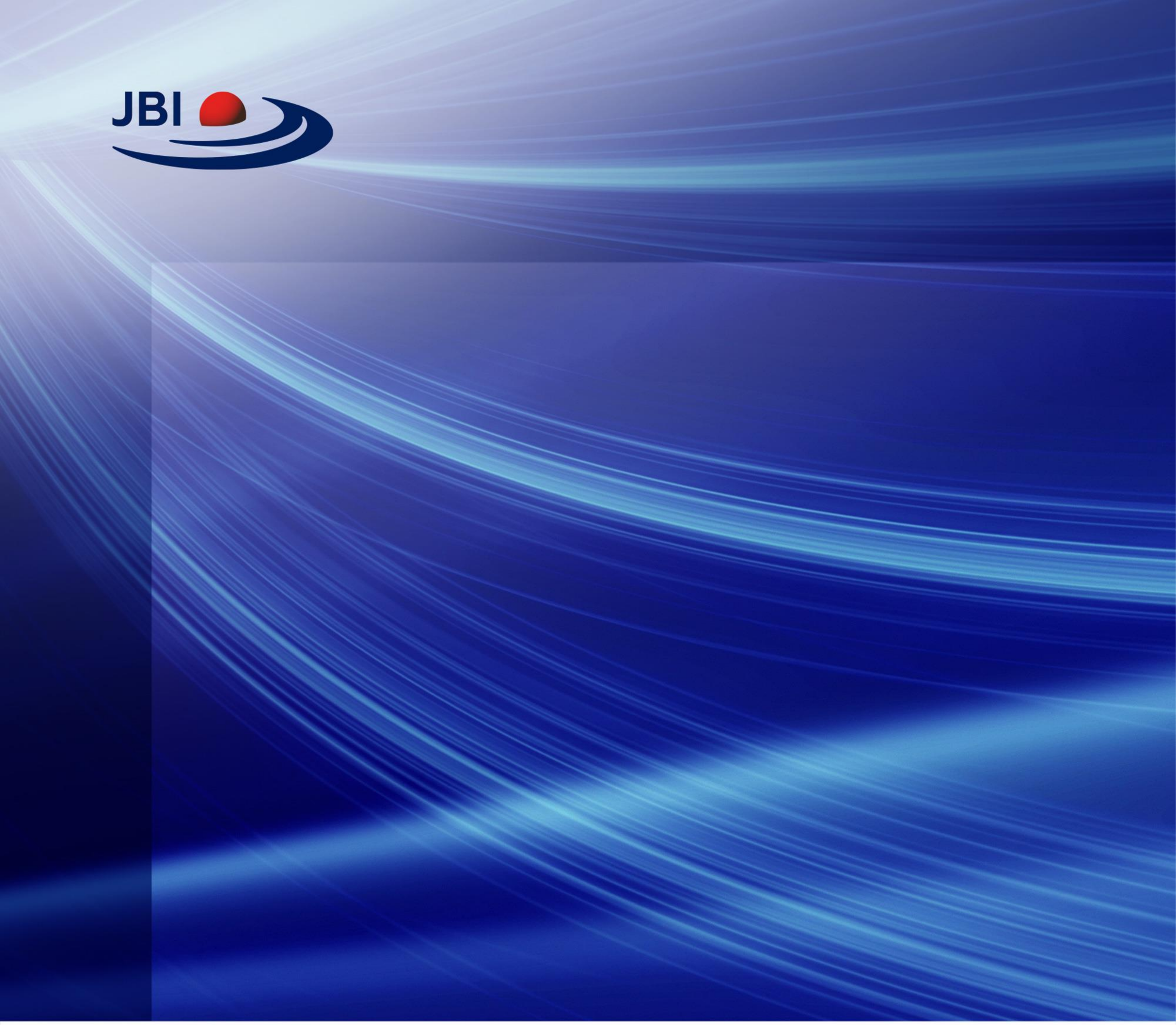

### CHECKLIST FOR SYSTEMATIC REVIEWS AND RESEARCH SYNTHESES

Critical Appraisal tools for use in JBI Systematic Reviews

jbi.global

CRICOS Provider Number 00123M

### INTRODUCTION

JBI is an international research organisation based in the Faculty of Health and Medical Sciences at the University of Adelaide, South Australia. JBI develops and delivers unique evidence-based information, software, education and training designed to improve healthcare practice and health outcomes. With over 70 Collaborating Entities, servicing over 90 countries, JBI is a recognised global leader in evidence-based healthcare.

Overall appraisal: Include ☐ Exclude ☐ Seek further info ✓

Comments (Including reason for exclusion)

As this report is not a systematic review, but an analysis of cases based on literature review some of the questions in the jbi checklist for systematic reviews may not be applicable. overall, the quality of the report appears to be good, but there is some missing information that would be helpful in fully assessing the quality of the study since it seems to be a simple literature review and the choice of assessment tools for these cases is difficult to assess these kind of studies.
