## Supplementary material for "Real-world data: A systematic literature review on the barriers, challenges, and opportunities associated with their inclusion in the health technology assessment process": S2 Appendix. Quality assessment of included studies: Checklist_for_Systematic_Reviews_and_Research_Syntheses_Fuchs S, et al (2016).pdf

Overall appraisal: Include ✓ Exclude ☐ Seek further info ☐

Comments (Including reason for exclusion)

This study has some strengths and weaknesses. the review question was clearly and explicitly stated, and the search strategy was appropriate with adequate sources and resources used to search for studies. however, the criteria for appraising studies were not appropriate, and critical appraisal was not conducted by two or more reviewers independently, which could affect the validity of the study. there were methods to minimize errors in data extraction, and the methods used to combine studies were appropriate. the likelihood of publication bias was not assessed. the study supported recommendations for policy and/or practice, and the specific directives for new research were appropriate. overall, the study has some strengths but also some weaknesses, and the evidence rate may be moderate or low depending on the weight given to the limitations of the study.  overall, the study has some limitations in terms of the appraisal of studies and the critical appraisal process, but it provides good quality evidence to support the use of real-world data in the assessment of health technologies.
