## Supplementary material for "Real-world data: A systematic literature review on the barriers, challenges, and opportunities associated with their inclusion in the health technology assessment process": S2 Appendix. Quality assessment of included studies: Checklist_for_Systematic_Reviews_and_Research_Syntheses_Kamusheva M, et al, (2022).pdf

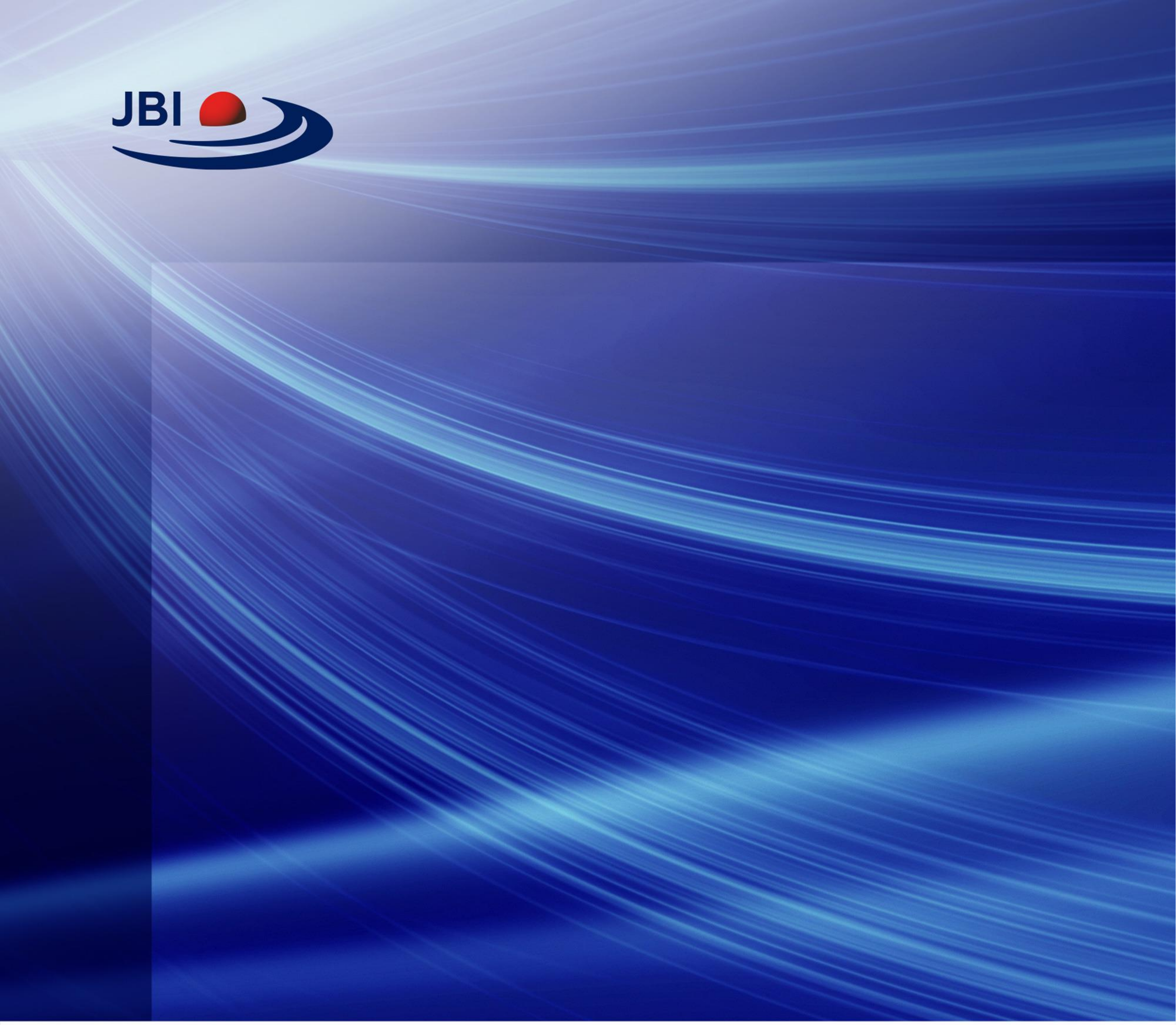

### CHECKLIST FOR SYSTEMATIC REVIEWS AND RESEARCH SYNTHESES

Critical Appraisal tools for use in JBI Systematic Reviews

jbi.global

CRICOS Provider Number 00123M

### INTRODUCTION

JBI is an international research organisation based in the Faculty of Health and Medical Sciences at the University of Adelaide, South Australia. JBI develops and delivers unique evidence-based information, software, education and training designed to improve healthcare practice and health outcomes. With over 70 Collaborating Entities, servicing over 90 countries, JBI is a recognised global leader in evidence-based healthcare.

Overall appraisal: Include ✓ Exclude ☐ Seek further info ☐

Comments (Including reason for exclusion)

The study seems to be a well-conducted systematic review with a clear research question, appropriate inclusion criteria, and a comprehensive search strategy. the study also followed appropriate methods for critical appraisal, data extraction, and synthesis of findings. however, the study did not report an assessment of publication bias. overall, if the study is relevant to your systematic review on the role of real-world evidence in health technology assessment, it may be a valuable source of information.
