## Supplementary material for "Real-world data: A systematic literature review on the barriers, challenges, and opportunities associated with their inclusion in the health technology assessment process": S2 Appendix. Quality assessment of included studies: Checklist_for_Systematic_Reviews_and_Research_Syntheses_Kent S et al, (2021).pdf

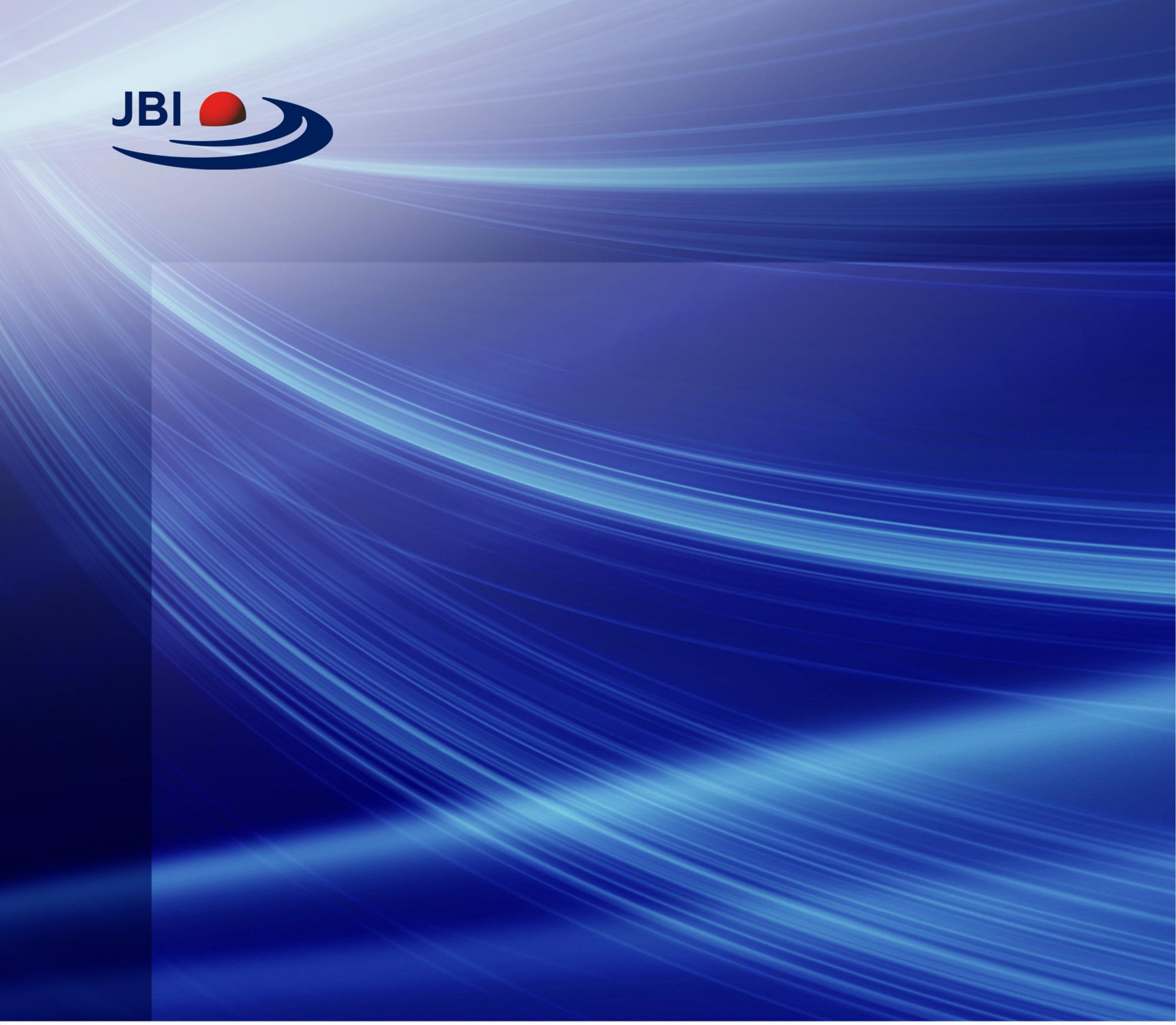

### CHECKLIST FOR SYSTEMATIC REVIEWS AND RESEARCH SYNTHESES

Critical Appraisal tools for use in JBI Systematic Reviews

jbi.global

CRICOS Provider Number 00123M

### INTRODUCTION

JBI is an international research organisation based in the Faculty of Health and Medical Sciences at the University of Adelaide, South Australia. JBI develops and delivers unique evidence-based information, software, education and training designed to improve healthcare practice and health outcomes. With over 70 Collaborating Entities, servicing over 90 countries, JBI is a recognised global leader in evidence-based healthcare.

Overall appraisal: Include ✓ Exclude ☐ Seek further info ☐

Comments (Including reason for exclusion)

This study includes pragmatic and ad-hoc reviews and seems to be of moderate quality based on the above answers. it is not a clear systematic review and probably this is the main reason of non-inclusion of several points indicated above in the questionnaire.
