## Supplementary material for "Real-world data: A systematic literature review on the barriers, challenges, and opportunities associated with their inclusion in the health technology assessment process": S2 Appendix. Quality assessment of included studies: Checklist_for_Systematic_Reviews_and_Research_Syntheses_Leahy T P, et al, (2020) .pdf

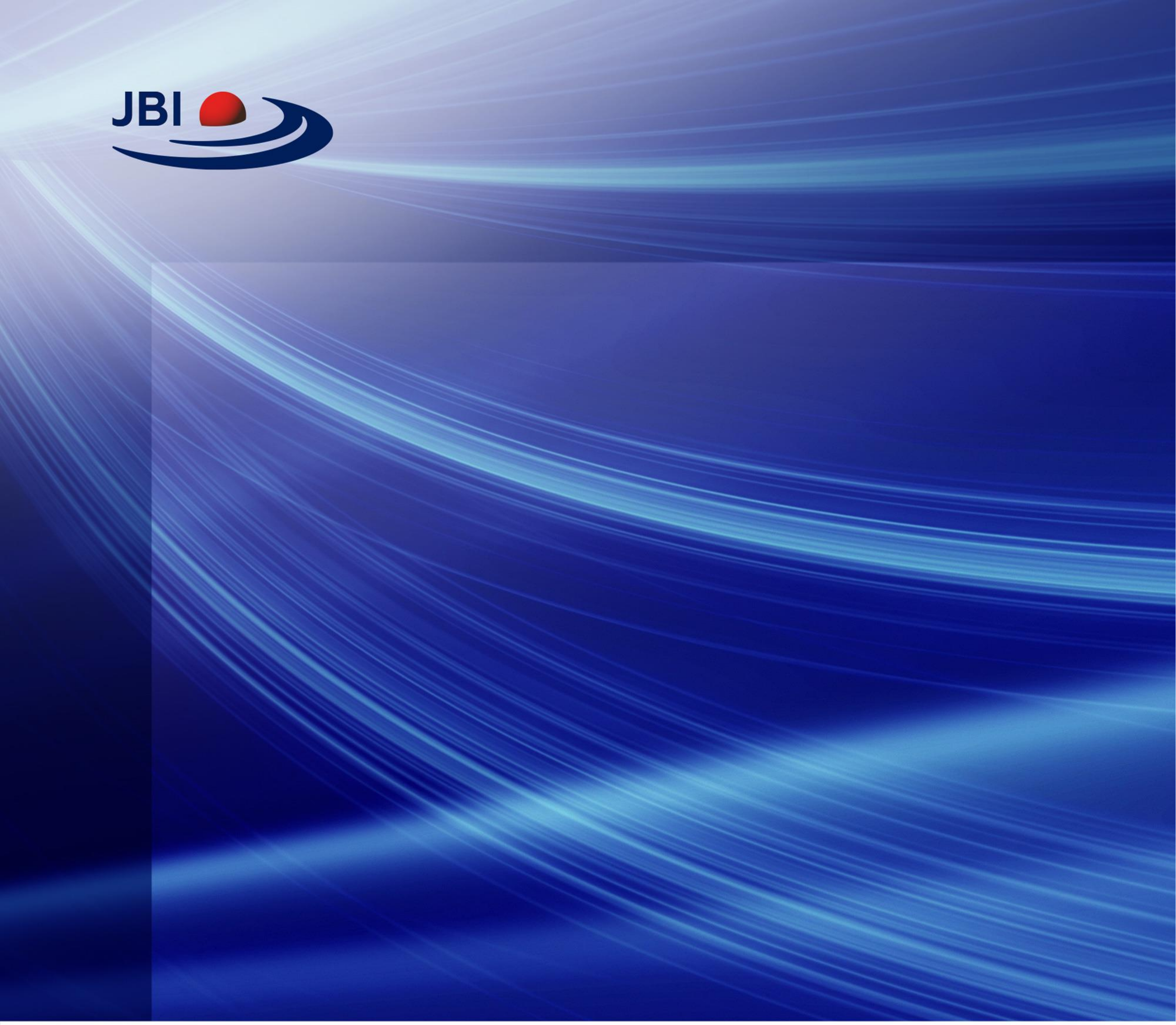

### CHECKLIST FOR SYSTEMATIC REVIEWS AND RESEARCH SYNTHESES

#### Critical Appraisal tools for use in JBI Systematic Reviews

jbi.global

CRICOS Provider Number 00123M

### INTRODUCTION

JBI is an international research organisation based in the Faculty of Health and Medical Sciences at the University of Adelaide, South Australia. JBI develops and delivers unique evidence-based information, software, education and training designed to improve healthcare practice and health outcomes. With over 70 Collaborating Entities, servicing over 90 countries, JBI is a recognised global leader in evidence-based healthcare.

Overall appraisal: Include ✓ Exclude ☐ Seek further info ☐

Comments (Including reason for exclusion)

The findings of this study suggest that when planning Health Economics and Outcomes Research (HEOR) strategies to support market access in England, greater consideration should be given to the inclusion and purposeful utilization of UK primary care databases in NICE submissions. This can help enhance the quality and relevance of real-world evidence in the technology assessment process. This study is of moderate quality.

© JBI, 2020. All rights reserved. JBI grants use of these tools for research purposes only. All other enquiries should be sent to.

### JBI CRITICAL APPRAISAL CHECKLIST FOR SYSTEMATIC REVIEWS AND RESEARCH SYNTHESIS



© JBI, 2020. All rights reserved. JBI grants use of these tools for research purposes only. All other enquiries should be sent to.
