## Supplementary material for "Real-world data: A systematic literature review on the barriers, challenges, and opportunities associated with their inclusion in the health technology assessment process": S2 Appendix. Quality assessment of included studies: Checklist_for_Text_and_Opinion_George E, (2016).pdf

Reviewer: Konstantinos Zisis  Date: 18 Apr 2023

Author: Elisabeth George  Year: 2016  Record Number: N/A

| | Yes | No | Unclear | Not applicable |
|---|---|---|---|---|
| 1. Is the source of the opinion clearly identified? | ✓ | ☐ | ☐ | ☐ |
| 2. Does the source of opinion have standing in the field of expertise? | ☐ | ☐ | ✓ | ☐ |
| 3. Are the interests of the relevant population the central focus of the opinion? | ✓ | ☐ | ☐ | ☐ |
| 4. Is the stated position the result of an analytical process, and is there logic in the opinion expressed? | ✓ | ☐ | ☐ | ☐ |
| 5. Is there reference to the extant literature? | ✓ | ☐ | ☐ | ☐ |
| 6. Is any incongruence with the literature/sources logically defended? | ☐ | ☐ | ✓ | ☐ |

Overall appraisal: Include ✓ Exclude ☐ Seek further info ☐

Comments (Including reason for exclusion)

The article provides a thoughtful and informative perspective on the use of real-world data in health technology assessment. The author presents a well-reasoned argument for the potential benefits of using real-world data to complement traditional randomized controlled trials in HTA, and offers recommendations for how to ensure the validity and reliability of real-world data.

© JBI, 2020. All rights reserved. JBI grants use of these tools for research purposes only. All other enquiries should be sent to.

### EXPLANATION OF TEXT AND EXPERT OPINION CRITICAL APPRAISAL TOOL

How to cite: *McArthur A, Klugarova J, Yan H, Florescu S. Innovations in the systematic review of text and opinion. Int J Evid Based Healthc. 2015;13(3):188–195.*

Answers: Yes, No, Unclear or Not/Applicable

1. **Is the source of the opinion clearly identified?**

   Is there a named author? Unnamed editorial pieces in journals or newspapers, or magazines give broader licence for comment, however authorship should be identifiable.

2. **Does the source of opinion have standing in the field of expertise?**

   The qualifications, current appointment and current affiliations with specific groups need to be stated in the publication and the reviewer needs to be satisfied that the author(s) has some standing within the field.

3. **Are the interests of the relevant population the central focus of the opinion?**

   The aim of this question is to establish the author's purpose in writing the paper by considering the intended audience. If the review topic is related to a clinical intervention, or aspect of health care delivery, a focus on health outcomes will be pertinent to the review. However, if for example the review is focused on addressing an issue of inter-professional behaviour or power relations, a focus on the relevant groups is desired and applicable. Therefore this question should be answered in context with the purpose of the review.

4. **Is the stated position the result of an analytical process, and is there logic in the opinion expressed?**

   In order to establish the clarity or otherwise of the rationale or basis for the opinion, give consideration to the direction of the main lines of argument. Questions to pose of each textual paper include: What are the main points in the conclusions or recommendations? What arguments does the author use to support the main points? Is the argument logical? Have important terms been clearly defined? Do the arguments support the main points?

5. **Is there reference to the extant literature?**

   If there is reference to the extant literature, is it a non-biased, inclusive representation, or is it a non-critical description of content specifically supportive of the line of argument being put forward? These considerations will highlight the robustness of how cited literature was managed.

6. **Is any incongruence with the literature/sources logically defended?**

   Is there any reference provided in the text to ascertain if the opinion expressed has wider support? Consider also if the author demonstrated awareness of alternate or dominant opinions in the literature and provided an informed defence of their position as it relates to other or similar discourses.

© JBI, 2020. All rights reserved. JBI grants use of these tools for research purposes only. All other enquiries should be sent to.
